# Convalescent Plasma in Critically ill Patients with Covid-19

**DOI:** 10.1101/2021.06.11.21258760

**Authors:** The REMAP-CAP Investigators, Lise J. Estcourt

## Abstract

**BACKGROUND:** The evidence for benefit of convalescent plasma for critically ill patients with Covid-19 is inconsistent. We hypothesized that convalescent plasma would improve outcomes for critically ill adult patients with Covid-19.

**METHODS:** In an ongoing adaptive platform trial, critically ill patients with confirmed Covid-19, defined as receiving intensive care-level organ support, were randomized to open-label convalescent plasma or not (i.e., control group). The primary end point was organ support-free days (i.e., days alive and free of ICU-based organ support) up to day 21. The primary analysis was a Bayesian cumulative logistic model with predefined criteria for superiority or futility. An odds ratio greater than 1 represented improved survival, more organ support–free days, or both.

**RESULTS:** The convalescent plasma intervention was stopped after pre-specified criteria for futility were met. At that time, 1084 participants had been randomized to convalescent plasma and 916 to no convalescent plasma (control). The median organ support-free days were 0 (interquartile range, -1 to 16) for the convalescent plasma group and 3 (interquartile range, -1 to 16) days for the control group. The median adjusted odds ratio (OR) was 0.97 (95% credible interval 0.83 to 1.15) and posterior probability of futility (OR < 1.2) was 99.4% for convalescent plasma compared to control. In-hospital mortality was 37.3% (401/1075) in convalescent plasma group, and 38.4% (347/904) in controls. The observed treatment effects were consistent across primary and secondary outcomes.

**CONCLUSIONS:** In critically ill adults with confirmed Covid-19, treatment with convalescent plasma, did not improve clinical outcomes.

Clinicaltrials.gov: NCT02735707

## Introduction

Coronavirus disease 2019 (Covid-19) is an acute illness caused by the severe acute respiratory syndrome coronavirus 2 (SARS-CoV-2).^1^ Amongst the many treatments studied,^2^ only two immunomodulatory drugs have been shown to save lives in hospitalized adults with Covid-19 - corticosteroids and interleukin-6 receptor antagonists,^3,4^ with critically ill patients showing greater benefit with corticosteroids.^5^ Convalescent plasma (blood product containing SARS-CoV-2 specific antibodies), is a biologically plausible antiviral treatment with immunomodulatory properties^6-9^ that may offer greater benefit to critically ill patients with Covid-19, in whom multiorgan dysfunction could be driven by the higher prevalence of viral RNAemia, and by progressive host response.^10,11^ However, convalescent plasma use in Covid-19 patients has either been outside of clinical trials, with 500,000 estimated to have received it in the United States,^12^ or in randomized clinical trials (RCTs) that have not focused on critically ill patients with Covid-19, including the recently published RECOVERY Trial.^13-19^

Therefore, we conducted an international, multicenter randomized clinical trial (RCT) to address this uncertainty in the evidence and to determine whether convalescent plasma compared to no convalescent plasma (control) improves outcomes in critically ill patients with Covid-19 within the Randomized, Embedded, Multifactorial, Adaptive Platform Trial for Community-Acquired Pneumonia (REMAP-CAP).

## Methods

### Trial Design

REMAP-CAP is an international, multicenter, open-label adaptive platform designed to determine the best treatment strategies for patients with severe pneumonia in both pandemic and non-pandemic settings. REMAP-CAP’s design and results regarding corticosteroids, anticoagulants, antivirals, and IL-6 receptor antagonists (IL-6ra) in Covid-19 have been reported previously.^3,4,20-22^

Patients are assessed for eligibility and randomized to different interventions across several domains. The trial is overseen by an International Trial Steering Committee (ITSC) blinded to the treatment allocation and an independent Data and Safety Monitoring Board (DSMB). REMAP-CAP is funded through multiple sources, approved by relevant regional research ethics committees, and conducted in accordance with Good Clinical Practice guidelines and the principles of the Declaration of Helsinki.

The Immunoglobulin Domain evaluating convalescent plasma enrolled participants at trial sites in Australia, Canada, the United Kingdom, and the United States. Informed consent, in accordance with local regulations, was obtained from all patients or their surrogates. Details of the trial design have been reported previously and in the **Protocol**.^20^

### Participants

Patients aged 18 years or older with confirmed SARS-CoV-2 infection admitted to hospital and classified as moderately or severely ill were eligible for enrollment in the Covid-19 Immunoglobulin Domain, equivalent to severely or critically ill respectively, as per the World Health Organization (WHO) case definitions.^23^ Outcomes for the critically ill participants, defined as patients admitted to an intensive care unit (ICU) and receiving respiratory or cardiovascular organ support, are reported in this manuscript.^3,4,20-22^

REMAP-CAP exclusion criteria included presumption that death was imminent with lack of commitment to full support, or participation in REMAP-CAP in the prior 90 days. Immunoglobulin Domain-specific exclusion criteria included: known hypersensitivity to convalescent plasma; objection to receiving plasma products; previous history of transfusion-related acute lung injury (TRALI; and more than 48h elapsed since ICU admission or 14 days since hospital admission. Further details regarding eligibility are listed in the **Protocol** and **Supplementary Appendix**.

### Treatment Allocation

The Covid-19 Immunoglobulin Domain contained three interventions, convalescent plasma at randomization, delayed convalescent plasma (given if clinical deterioration - only available at one site in the United States), and control. Participants were randomized via a centralized computer program to each intervention (available locally) starting with balanced assignment. The funders of the trial had no role in trial design, data collection, data analysis, data interpretation, or writing of the manuscript. Members of the writing committee had full access to all data and had final responsibility for the decision to submit for publication.

### Procedures

The trial used an open-label design, where convalescent plasma was supplied by each site’s transfusion laboratory, and the clinical team were provided instructions for convalescent plasma administration. Details of donor selection, plasma manufacture, testing of convalescent plasma in each country, and convalescent plasma administration are in the **Supplementary Appendix**.

Other aspects of care were provided as per each site’s standard of care. In addition to assignments in the Immunoglobulin Domain, participants could be randomized to additional interventions within other domains, depending on domains active at the site, patient eligibility, and consent (see www.remapcap.org). Randomization to the Corticosteroid Domain for Covid-19 closed on June 17, 2020.^20^ Thereafter, corticosteroids were allowed as per recommended standard of care.

Although clinical staff were aware of their individual patient’s intervention assignment, neither they nor the ITSC were provided any information about aggregate patient outcomes.

### Interventions

Participants were randomized to receive high-titer ABO compatible convalescent plasma (total volume approximately 550 +/- 150 ml) within 48 hours of randomization, or control. All convalescent plasma used in the trial were tested for SARS-CoV-2 antibodies and met a minimum titer criterion prior to administration. The details of the method of testing and minimum titer criterion for convalescent plasma in each country are in the **Supplementary Appendix**.

### Outcomes

The primary outcome was respiratory and cardiovascular organ support-free days up to day 21. The definitions of respiratory and cardiovascular organ support were the same as in the inclusion criteria. In this composite ordinal outcome, all deaths within the hospital, up to day 90, are assigned the worst outcome (–1 day). Among survivors, respiratory and cardiovascular organ support-free days were calculated up to day 21, such that a higher number represents faster recovery. Prespecified secondary outcomes included: 28-day survival; 90-day survival; progression to invasive mechanical ventilation, extra corporeal mechanical oxygenation (ECMO) or death; intensive care and hospital length-of-stay; and World Health Organization ordinal scale at day 14. (**Supplementary Appendix)**.

### Trial Power and Sample Size

REMAP-CAP uses a Bayesian design with no maximum sample size. Regular adaptive analyses are conducted, and randomization continues, potentially with response-adaptive randomization with preferential assignment to those interventions that appear most favorable, until a pre-defined statistical trigger of superiority or futility is met. Response-adaptive randomization was used in the Immunoglobulin Domain from November 23,2020.

### Statistical Analysis

The statistical analysis plan for the Covid-19 Immunoglobulin Domain was written blinded to treatment allocation and posted online (www.remapcap.org) before data lock and analyses (**Protocol and SAP**). The primary analysis was generated from a Bayesian cumulative logistic model, which calculated posterior probability distributions of organ support-free days at day 21 (primary outcome) based on evidence accumulated in the trial and assumed prior knowledge in the form of a prior distribution. Prior distributions for individual treatment effects were neutral. The primary model adjusted for location (site, nested within country), age (categorized into six groups), sex, and time-period (two-week epochs). The model contained treatment effects for each intervention within each domain.

The primary analysis was conducted on all randomized patients as of, January 18, 2021, and not just those randomized within the Immunoglobulin Domain. This approach allowed maximal incorporation of all information, providing the most robust estimation of the coefficients of all included covariates. Importantly, not all patients were eligible for all domains nor for all interventions within each domain (depending on site participation, baseline entry criteria, and patient or surrogate preference).

Because the primary model included information about assignment to interventions within domains whose evaluation is on-going, it was run by the Statistical Analysis Committee unblinded to the treatment allocation (**Protocol and SAP**), who conduct all protocol-specified trial adaptive analyses and report results to the DSMB.

The cumulative log odds for the primary endpoint were modeled such that a parameter >0 reflects an increase in the cumulative odds for the organ support free-days outcome, which implies benefit. The model assumed proportional effects across the ordinal organ support-free days scale. There was no imputation of missing primary (or secondary) endpoint values.

The posterior distribution of the odds ratios is summarized by, allowing calculation of odds ratios with 95% credible intervals (CrI) and the probability that convalescent plasma was superior or futile compared with control. An odds ratio >1 represents improved survival and/or more organ support-free days. The pre-defined statistical triggers for trial conclusions and disclosure of results were: superiority if >99% posterior probability the odds ratio was >1 compared with control; or futility if >95% posterior probability the odds ratio was <1.2 compared with control.

Analysis of the primary outcome was then repeated using only data from those critically ill patients with Covid-19 enrolled in domains that had stopped and were unblinded at the time of analysis with no adjustment for assignment in other ongoing domains. The secondary outcomes were also analyzed in this secondary analysis population. The prespecified six subgroups were binary categories for: presence of SARS-CoV-2 antibodies at baseline; detectable virus at baseline in an upper respiratory sample; need for mechanical ventilation; immunosuppressed state; time from hospitalization to enrolment in recipients (categorized as <3, 3-7 and >7 days); and antibody titer of the convalescent plasma transfused. Further details of all analyses are provided in the **Supplementary Appendix**. Pre-specified analyses are listed in the statistical analysis plan (**Protocol and SAP**).

## Results

At a scheduled adaptive analysis, the statistical trigger for futility in critically ill participants with Covid-19 was met (posterior probability of futility 96.4%, (OR 0.95, 95% Credible Interval (CrI) 0.73 to 1.23). Assignment to this domain closed on January 11, 2021 for critically ill participants (randomization continued for participants who were not critically ill). After announcement of the preliminary RECOVERY trial results on January 15,2021, the ITSC halted recruitment to all patients within the domain.^24^

### Participants

Between March 9, 2020 and January 18, 2021, of 10,282 screened patients, 4,763 who were hospitalized with Covid-19, were enrolled in REMAP-CAP, and randomized within at least one therapeutic domain (Fig. 1). Patients were recruited to the Immunoglobulin Domain from May 5, 2020 at 129 sites. Among the 2,097 participants enrolled in the domain, 2,011 were critically ill, and were randomly assigned to convalescent plasma at randomization (n=1084), convalescent plasma if clinical deterioration (n=11), and control (n=916). Further information about hospitalized participants with Covid-19 who were not critically ill and participants in the convalescent plasma if clinical deterioration group are in the **Supplementary Appendix**.

**Figure 1.**
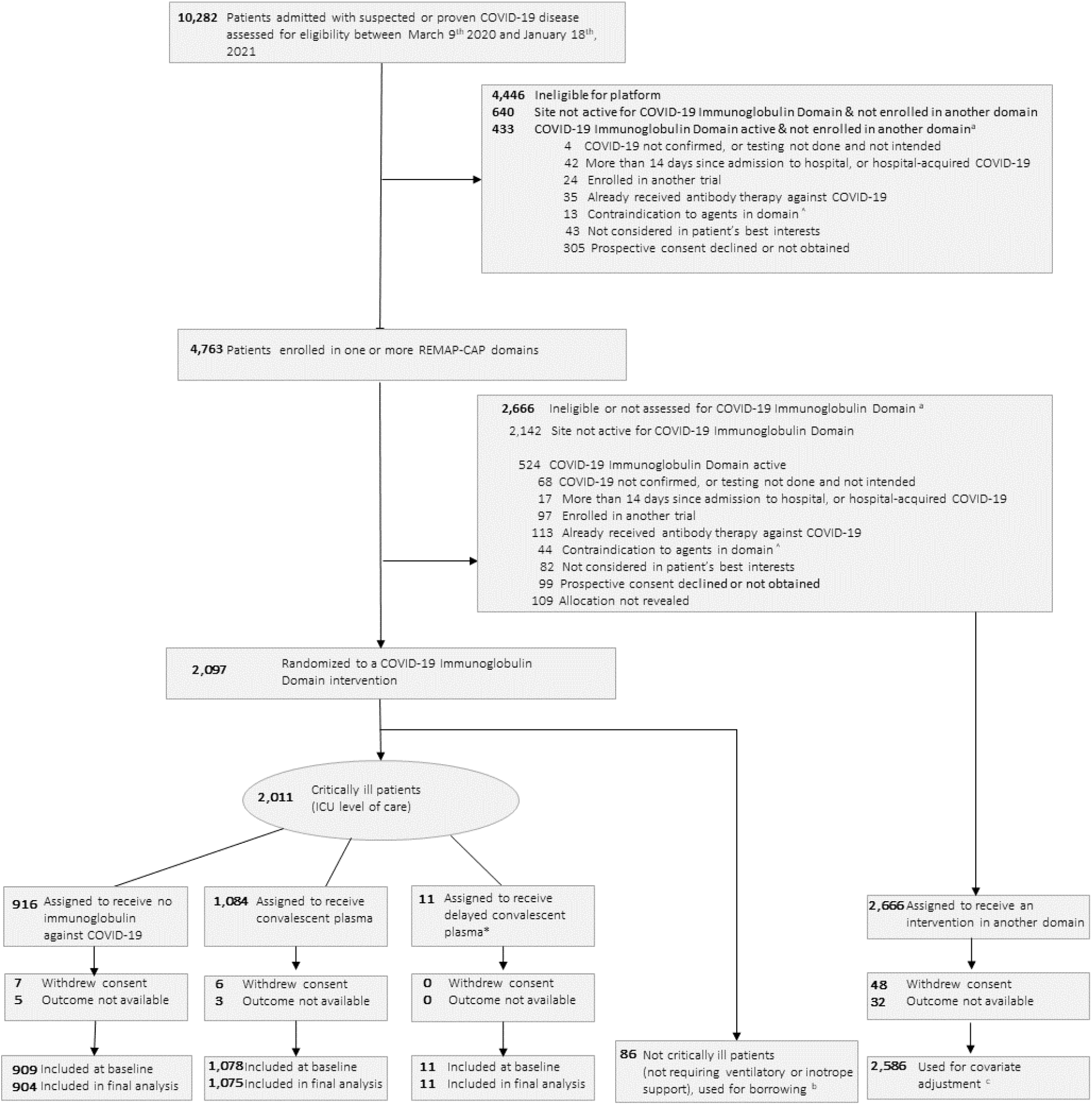
Screening, Randomization, and Follow-up of Participants in the REMAP-CAP COVID-19 Immunoglobulin Domain Randomized Clinical Trial. ^a^ Patients could meet more than 1 ineligibility criterion. ^b^ Results for patients who were not critically ill were used for borrowing within the primary model ^c^ The primary analysis of alternative interventions within the Immunoglobulin Domain is estimated from a model that adjusts for patient factors and for assignment to interventions in other domains. To obtain the most reliable estimation of the effect of these patient factors and of other interventions on the primary outcome, all patients enrolled in the COVID-19 cohort (for whom there is consent and follow-up) are included. Importantly, however, the model also factors eligibility for the Immunoglobulin Domain and its interventions, such that the final estimate of an Immunoglobulin Domain intervention’s effectiveness relative to any other within that domain is generated from those patients that might have been randomized to either.

**Figure 2.**
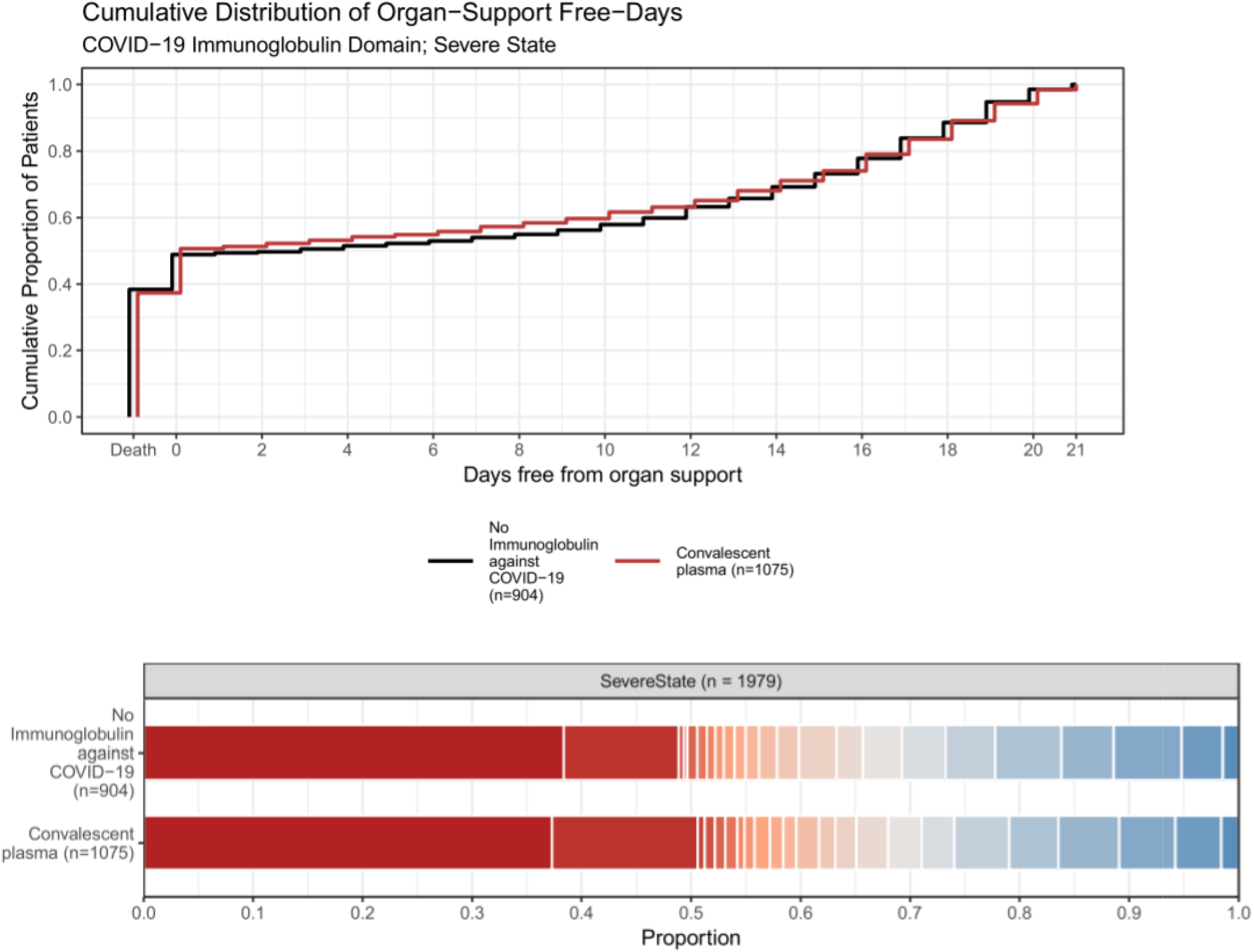
Organ Support-free Days. The upper panel displays the distributions of organ support-free days (see Methods for definition) by trial group as the cumulative proportion (y axis) for each trial group by day (x axis), with death listed first. Curves that rise more slowly are more favorable. The lower panel displays organ support-free days as horizontally stacked proportions by trial group. Red represents worse values and blue represents better values. The median adjusted odds ratios from the primary analysis, using a Bayesian cumulative logistic model, was 0.97, 95% credible interval 0.83 to 1.15 for convalescent plasma group compared with the no convalescent plasma(control) group, yielding 37.8% probability of superiority and 99.4% probability of futility over the control group.

Thirteen participants subsequently withdrew consent, and eight participants had missing primary outcome data. The baseline characteristics of the participants randomized to convalescent plasma were similar across intervention groups and typical of patients requiring ICU care for Covid-19 (Table 1 and Table S1). All but three participants were receiving respiratory support at the time of randomization, including high flow nasal oxygen (22%), noninvasive (45%) and invasive (33%) mechanical ventilation.

**Table 1.** Participant Characteristics at Baseline.

| No. (%) of participants * |  |  |
| --- | --- | --- |
| Characteristic | Convalescent plasma<br>(N=1078) | Control<br>(N=909) |
| Age – mean (SD), years | 60.2 (12.7) | 60.2 (13.1) |
| Sex – Male | 727 (67.4) | 618 (68.0) |
| Race/Ethnicity<sup>†</sup> - White – n/N (%) | 731 / 976 (74.9) | 619 / 832 (74.4) |
| Asian – n/N (%) | 144 / 976 (14.8) | 133 / 832 (16.0) |
| Black – n/N (%) | 51 / 976 (5.2) | 38 / 832 (4.6) |
| Mixed – n/N (%) | 16 / 976 (1.6) | 8 / 832 (1.0) |
| Other<sup>†</sup> – n/N (%) | 34 / 976 (3.5) | 34 / 832 (4.1) |
| APACHE II score<sup>‡</sup> - median (IQR) | 13.0 (8.0 - 19.0)<br>(n = 1044) | 12.0 (8.0 - 19.0)<br>(n = 888) |
| Pre-existing conditions – n/N (%) |  |  |
| Diabetes mellitus | 339 / 1078 (31.4) | 268 / 907 (29.5) |
| Respiratory disease | 245 / 1078 (22.7) | 216 / 907 (23.8) |
| Kidney disease | 107 / 1000 (10.7) | 83 / 837 (9.9) |
| Severe cardiovascular disease | 96 / 1053 (9.1) | 67 / 890 (7.5) |
| Immunosuppressive disease or therapy | 67 / 1066 (6.3) | 60 / 907 (6.6) |
| SARS-CoV-2 RNA detected (respiratory sample) at randomization – n/N (%)<sup>§</sup> | 676 / 845 (80.0) | 489 / 599 (81.6) |
| SARS-CoV-2 Antibody negative at baseline – n/N (%) | 271 / 874 (31.0) | 149 / 558 (26.7) |
| Time to enrollment - median (IQR) |  |  |
| From hospital admission - days | 1.8 (1.0 - 3.3) | 1.7 (0.9 - 3.5) |
| From ICU admission - hours | 17.7 (10.2 - 23.5) | 17.2 (10.6 - 23.2) |
| Acute respiratory support |  |  |
| None/supplemental oxygen only | 2 (0.2) | 1 (0.1) |
| High flow nasal cannula | 225 (20.9) | 211 (23.2) |
| Non-invasive ventilation only | 493 (45.7) | 407 (44.8) |
| Invasive mechanical ventilation | 356 (33.0) | 289 (31.8) |
| ECMO – n/N (%) | 2 (0.2) | 1 (0.1) |
| **Vasopressor support** | 207 (19.2) | 175 (19.3) |

Abbreviations: SD, standard deviation; APACHE, Acute Physiology and Chronic Health Evaluation; IQR, interquartile range; ECMO, extracorporeal membrane oxygenation.
\* Unless otherwise indicated. Percentages may not sum to 100 because of rounding.
† Data collection not approved in Canada and continental Europe. 'Other' includes 'declined' and 'multiple'.
‡ Range: 0 - 71, with higher scores indicating greater severity of illness.
§ Classified as positive when the SARS-CoV-2 PCR test was positive between 24 hours before and after randomization.

### Interventions and co-interventions

In the convalescent plasma group, 85.6% (920/1075) received convalescent plasma as per protocol (definition **Supplementary Appendix**), and 94.5% (1016/1075) received some convalescent plasma. In the control group, 0.6% (5/905) received convalescent plasma. Most patients were enrolled after the announcement of the dexamethasone result from the RECOVERY trial,^25^ and therefore 94% (1859/1987) of participants were treated with corticosteroids (Table S1). Remdesivir use, as part of routine care, was recorded in 45% of participants. Most participants were enrolled before the announcement of the IL6-receptor antagonist result from the REMAP-CAP trial (Table S1 and S2).^4^

### Primary Outcome

The median number of organ support-free days was 0 (interquartile range, -1 to 16) in the convalescent plasma group and 3 (interquartile range, -1 to 16) in the control group (Fig.2 and Fig.3). Relative to control, the median adjusted odds ratio from the primary model was 0.97 (95% CrI 0.83 to 1.15), yielding a 99.4% posterior probability of futility. Hospital mortality was 37% (401/1075) for convalescent plasma and 38% (347/904) for control. Compared with control, the median adjusted odds ratio for in-hospital survival was 1.04 (95% CrI 0.85 to 1.27) for convalescent plasma, yielding a 91.8% posterior probability of futility. The sensitivity analyses were consistent with the primary analysis (Tables S4 and S5). Potential interactions between convalescent plasma and other interventions were evaluated and reported (Table S2 and S6). There were no significant interactions.

The pre-specified secondary analyses of the primary outcome using only data from participants in the Immunoglobulin Domain were consistent with the primary analysis (Table S3).

In the pre-specified subgroup analyses, based on participant characteristics at baseline, there were no differences seen in the effect of convalescent plasma based on SARS-CoV-2 PCR status, detectable anti-SARS-CoV-2 antibody at baseline, mechanical ventilation at baseline, or antibody titer in plasma product (Fig. 4). In the small number of participants (n=126) with immunodeficiency at baseline, convalescent plasma demonstrated potential benefit (89.8% posterior probability of superiority). In the subgroup of participants randomized more than 7 days into hospitalization (n=126) convalescent plasma may be harmful (90.3% posterior probability of harm, OR < 1.0).

### Secondary Outcomes

The secondary outcomes are presented in (Figure 3 and Table S3). Full model results of all outcome analyses are provided in eAppendices A and B in the **Supplementary Appendix**. Compared with control, convalescent plasma had treatment effects consistent with the primary outcome, for the pre-specified secondary outcomes.

**Figure 3a.**
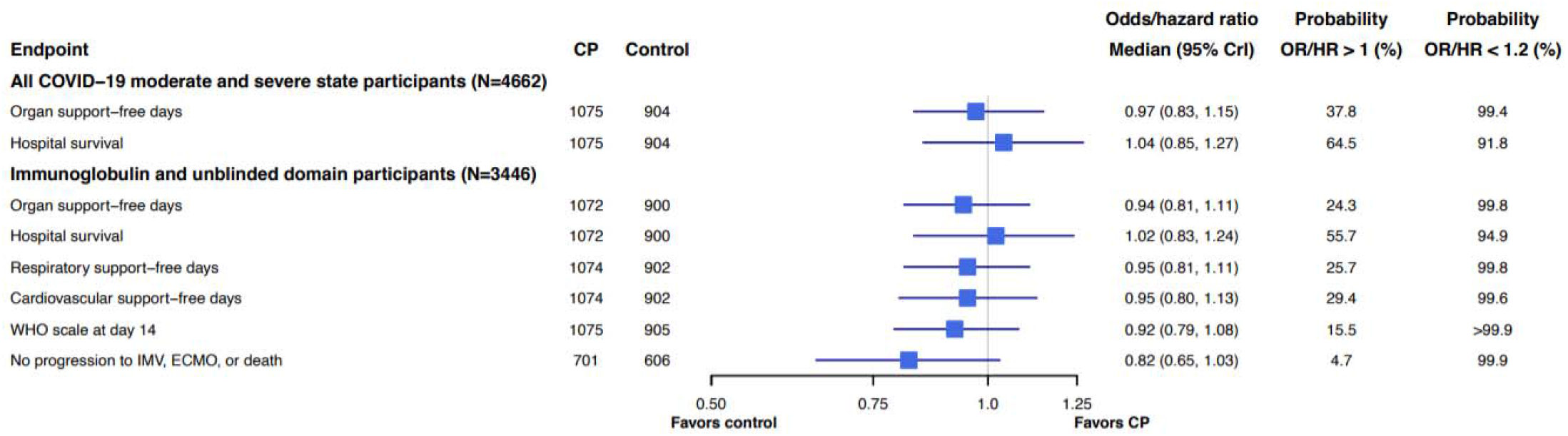
Dichotomous primary and secondary outcomes.

**Figure 3b.**
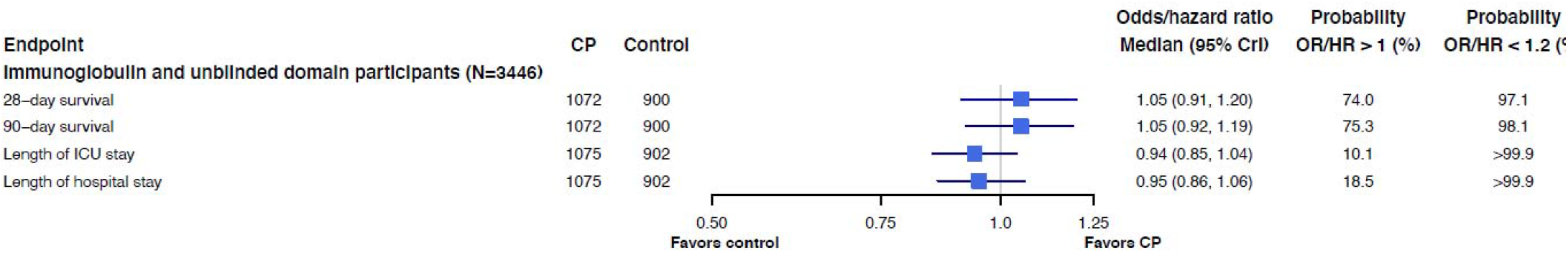
Time to event secondary outcomes. Data for secondary analyses excluded participants who had been randomized within another domain within the moderate stratum and then randomized to the immunoglobulin domain in the severe stratum (excluded 7 participants), maximum of 1980 participant included within secondary analyses (See Table S3). An odds ratio < 1.2 equates to the threshold for futility for the primary outcome

**Figure 4.**
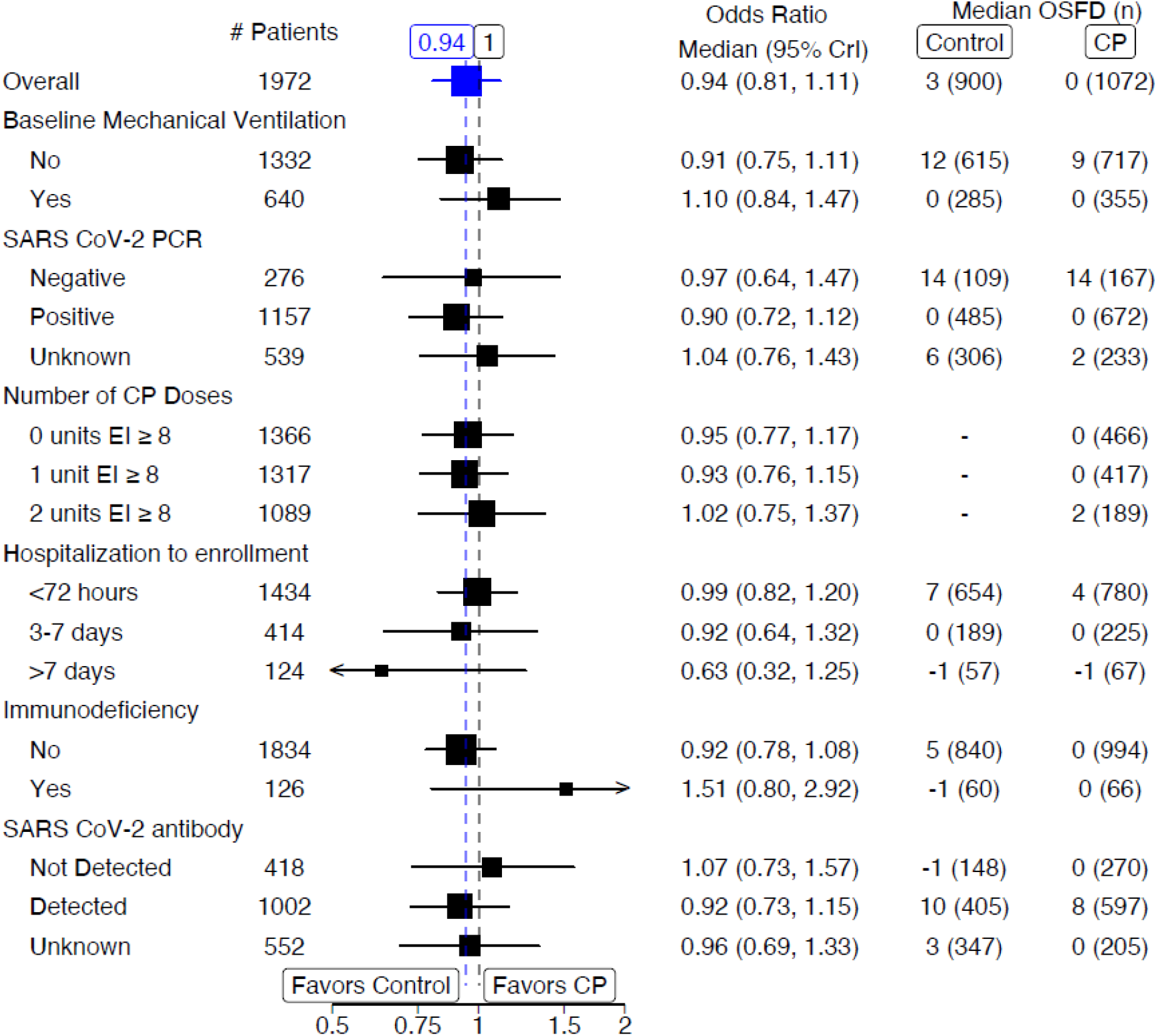
Pre-specified sub-group analyses*. Abbreviations: CP - convalescent plasma; EI – Euroimmun; PCR – polymerization chain reaction. *Data for sub-group analyses excluded participants who had been randomized within another domain within the moderate stratum and then randomized to the immunoglobulin domain in the severe stratum (excluded 7 participants), maximum of 1980 participant included within the sub-group analyses. An odds ratio < 1.2 equates to the threshold for futility for the primary outcome. For the number of convalescent plasma doses administered with a Euroimmun titer ≥ 8, the number of participants analyzed equal total number in the control group (900) plus the number in the intervention group who received those number of convalescent plasma doses. Immunodeficiency was defined as immunosuppressive treatment or disease (See supplementary appendix for full definition)

There were 44/1980 (2.2%) participants who had at least one serious adverse event, 32/1075 (3.0%), in the convalescent plasma group and 12/905 (1.3%) in the control group. (Table S7).

## Discussion

In critically ill patients with Covid-19, treatment with convalescent plasma did not improve organ-support free days compared with no convalescent plasma (control) with a 99.4% probability of futility. The observed treatment effects were consistent across secondary outcomes, and in all sensitivity analyses. Among the pre-defined subgroups, there was no evidence for heterogeneity of treatment effect with convalescent plasma compared with controls.

This trial’s results are consistent with the lack of benefit of convalescent plasma for patients hospitalized with moderate or severe Covid-19 reported in the RECOVERY trial,^13^ and with the meta-analyses within the Cochrane systematic review.^15,26^ In this trial, 75% of participants received advanced respiratory support which often occurs between 7-10 days after symptom onset, and by which stage in the disease many immunocompetent patients will have developed endogenous antibody responses responses.^14^ This could explain our overall results, including the effect observed in the pre-specified immunodeficiency subgroup (Fig. 4). These findings warrant further investigation of the hypotheses that benefit with convalescent plasma may be different early on in the illness,^14,27^ and perhaps in patients with an impaired immune system who are unable to mount effective immune responses, including antibody responses.^27,28^

The pre-specified anti-SARS-CoV-2 antibody and SARS-CoV-2 PCR status subgroups evaluated the hypotheses that the antibody negative population and respiratory PCR positive population may derive benefit from convalescent plasma therapy. Overall, anti-SARS-CoV-2 antibody negative participants had a higher mortality compared with antibody positive participants and the viral load in respiratory samples were high,^29^ consistent with greater prevalence of viral RNAemia in critically ill Covid-19 patients.^10^ However, no differences in treatment effect were observed within these two subgroups, even when RECOVERY data were taken into consideration (Figure S2).

Strengths of this trial include a design that assessed the impact of administration of high-titer convalescent plasma in critically ill adults on patient-centered outcomes, with the majority of participants (72.7%) randomized within three days of hospitalization. In this trial, the dose of convalescent plasma administered, was at least one unit Euroimmun (EI) titer ≥8 in 56.6% of participants and EI≥6 in all participants for which data were available, which represents higher titers than the emergency use authorization recommendation by the FDA (EI≥3.5).^30^

The trial has limitations. The trial used an open-label design, although clinician and patient awareness of trial assignment likely had minimal effect on the primary outcome. Only 85.9% of participants received convalescent plasma in the intervention group as per protocol and 0.6% of patients in the control group received convalescent plasma, however, this is unlikely to have biased the results towards null, as the per protocol analyses were similar to the primary analysis. Although most participants received very high titer convalescent plasma, viral neutralization properties were not measured prior to administration. Finally, the trial has only been able to test the potential effectiveness of convalescent plasma in critically ill patients and it remains possible that convalescent plasma or other high-titer antibody-based therapy (alone or in combination with anti-viral chemotherapy) may have a therapeutic effect earlier in the disease process or in some patient groups.

## Conclusions

In critically ill patients with Covid-19, treatment with convalescent plasma did not improve clinical outcomes, including organ-support free days or hospital survival.

## Data Availability

Data will be available to researchers on request subject to sponsor restrictions from 1-12-2022. Please contact

## Article Information

### Authors/Members of the Writing Committee

Lise J. Estcourt, MB BChir, DPhil^1,2*^;Alexis F. Turgeon, MD, MSc^3,4*^; Zoe K. McQuilten, MBBS, PhD^5,6*^; Bryan J. McVerry, MD^7*^; Farah Al-Beidh, PhD^8^; Djillali Annane, MD, PhD^9,10,11^; Yaseen M. Arabi, MD^12^; Donald M. Arnold MD MSc^13^; Abigail Beane, MSc^14^; Philippe Bégin MD PhD^15^; Wilma van Bentum-Puijk, MS^16^; Lindsay R. Berry, PhD^17^; Zahra Bhimani, MPH, PMP^18^; Janet E. Birchall, MBChB^19^; Marc J.M. Bonten, MD^16,20^; Charlotte A. Bradbury, MD, PhD,^21,22^; Frank M. Brunkhorst, MD^23^; Meredith Buxton, PhD^24^; Jeannie L. Callum, MD^25,26,27^; Michael Chassé MD, PhD^15^; Allen C. Cheng, MD^28, 29^; Matthew E. Cove, MBChB^30^; James Daly, MBBS^31^; Lennie Derde, MD^16,32^; Michelle A. Detry, PhD^17^; Menno De Jong, MD^33^; Amy Evans, MMedSci^34^; Dean A. Fergusson, PhD^35^; Matthew Fish, MSc^36^; Mark Fitzgerald, PhD^17^; Claire Foley, BSc^34^; Herman Goossens, MD^37^; Anthony C. Gordon, MD^8^;; Iain B. Gosbell, MBBS, MD^31,38^; Cameron Green, MSc^29^; Rashan Haniffa, MD^39,40^; Heli Harvala MD, PhD^41^; Alisa M. Higgins, PhD^29^; Thomas E. Hills, MBChB, DPhil^42^; Veronica C. Hoad, MBBS, MPH^31^; Christopher Horvat MD MHA^43^; David T. Huang MD MPH^44^; Cara L. Hudson MSc^45^; Nao Ichihara MD, MPH, PhD^46^; Emma Laing BSc^34^; Abigail A. Lamikanra, PhD^1^; Francois Lamontagne, MD, MSc^47^; Patrick R. Lawler, MD, MPH^48^; Kelsey Linstrum, MS^49^; Edward Litton, MD^50^; Elizabeth Lorenzi, PhD^17^; Sheila McLennan, MB BS^51^; John Marshall, MD^18,52^; Daniel F. McAuley, MD^53^; John F. McDyer, MD^7^; Anna McGlothlin, PhD^17^; Shay McGuinness, MD^29,54,55^; Gail Miflin^56^; Stephanie Montgomery, MSc^49,57^; Paul R. Mouncey, MSc^58^; Srinivas Murthy, MD^59^; Alistair Nichol, MD^29,60,61,62^; Rachael Parke, RN, PhD^54,55,63^; Jane C. Parker, RN^29^; Nicole Pridee, MB BCh^64^; Damian F.J. Purcell, PhD^65^; Luis F. Reyes, MD, PhD^66,67^; Peter Richardson, BPharm^19^; Nancy Robitaille, MD^68,69,70^; Kathryn M. Rowan, PhD,^58^; Jennifer Rynne, MSc^36^; Hiroki Saito, MD, MPH^71^; Marlene Santos, MSc^18^; Christina T. Saunders, PhD^17^; Ary Serpa Neto, MD MSc PhD^29,72^; Christopher W. Seymour, MD, MSc^49,57^; Jon A. Silversides, PhD^53^; Alan A. Tinmouth MD MSc^35^; Darrell J. Triulzi, MD^73^; Anne M. Turner, RN, MPH^55^; Frank van de Veerdonk, MD^74^; Tim Walsh, MD^75^; Erica M. Wood^5,6^; Scott Berry, PhD^17^; Roger J. Lewis, MD, PhD^17,76,77^; David K. Menon MD, PhD^78**^; Colin McArthur, MD^58,79**^; Ryan Zarychanski, MD, MSc^80**^; Derek C. Angus, MD, MPH^49,57**^; Steven A. Webb, MD, PhD^4,20,32**^;David J. Roberts, MBChB, DPhil1,^2**^; Manu Shankar-Hari MD, PhD^36,81**^.

^*^ joint first author

^**^ joint last author

### Affiliations of Authors/Members of the Writing Committee

^1^ NHS Blood and Transplant, Oxford, UK

^2^ Radcliffe Department of Medicine and BRC Haematology Theme, University of Oxford, Oxford, UK

^3^ Department of Anesthesiology and Critical Care Medicine, Division of Critical Care Medicine, Université Laval, Québec City, Québec, Canada

^4^ CHU de Québec-Université Laval Research Center, Population Health and Optimal Health Practices Unit, Trauma – Emergency – Critical Care Medicine, CHU de Québec- Université Laval, Québec City, Québec, Canada

^5^ Transfusion Research Unit, School of Public Health & Preventive Medicine, Monash University, Melbourne, Australia

^6^ Department of Clinical Haematology, Monash Health, Melbourne, Australia

^7^ Department of Medicine, University of Pittsburgh School of Medicine, Pittsburgh, Pennsylvania, USA

^8^ Division of Anaesthetics, Pain Medicine and Intensive Care Medicine, Department of Surgery and Cancer, Imperial College London and Imperial College Healthcare NHS Trust, London, UK

^9^ Intensive Care Unit, Raymond Poincaré Hospital (AP-HP), Paris, France

^10^ Simone Veil School of Medicine, University of Versailles, Versailles, France

^11^ University Paris Saclay, Garches, France

^12^ Intensive Care Department, College of Medicine, King Saud Bin Abdulaziz University for Health Sciences, King Abdullah International Medical Research Center, Ministry of National Guard Health Affairs, Riyadh, Saudi Arabia

^13^ McMaster University, Hamilton, Ontario, Canada

^14^ Nuffield Department of Clinical Medicine, University of Oxford, Oxford, UK

^15^ Université de Montréal, Montréal, Québec, Canada

^16^ Julius Center for Health Sciences and Primary Care, University Medical Center Utrecht, Utrecht University, Utrecht, the Netherlands

^17^ Berry Consultants, LLC, Austin, Texas, USA

^18^ Li Ka Shing Knowledge Institute, Unity Health Toronto, St. Michael’s Hospital, Toronto, Ontario, Canada

^19^ Welsh Blood Service, UK

^20^ Department of Medical Microbiology, University Medical Center Utrecht, Utrecht University, Utrecht, the Netherlands

^21^ University Hospitals Bristol and Weston NHS Foundation Trust, Bristol, UK

^22^ Faculty of Health Sciences, University of Bristol, Bristol, UK

^23^ Center for Clinical Studies and Center for Sepsis Control and Care (CSCC), Department of Anesthesiology and Intensive Care Medicine, Jena University Hospital, Jena, Germany

^24^ Global Coalition for Adaptive Research, San Francisco, California, USA

^25^ Canadian Blood Services, Ottawa, Ontario, Canada

^26^ Department of Pathology and Molecular Medicine, Kingston Health Sciences Centre and Queens University, Kingston, Ontario, Canada

^27^ Department of Laboratory Medicine and Molecular Diagnostics, Sunnybrook Health Sciences Centre, Toronto, Ontario, Canada

^28^ Infection Prevention and Healthcare Epidemiology Unit, Alfred Health, Melbourne, Victoria, Australia

^29^ Australian and New Zealand Intensive Care Research Centre, School of Public Health and Preventive Medicine, Monash University, Melbourne, Victoria, Australia

^30^ Department of Medicine, Yong Loo Lin School of Medicine, National University of Singapore,1 E Kent Ridge Road, NUHS Tower Block, Level10, Singapore119228

^31^ Australian Red Cross Lifeblood, Australia

^32^ Intensive Care Center, University Medical Center Utrecht, Utrecht University, Utrecht, the Netherlands

^33^ Department of Medical Microbiology, Amsterdam UMC, University of Amsterdam, the Netherlands^1^ NHS Blood and Transplant, Bristol, UK

^34^ NHSBT Clinical Trials Unit, NHS Blood and Transplant, Cambridge, UK

^35^ Ottawa Hospital Research Institute, Clinical Epidemiology Unit, Canada

^36^ School of Immunology & Microbial Sciences, Kings College London, SE19 RT

^37^ Department of Microbiology, Antwerp University Hospital, Antwerp, Belgium

^38^ Western Sydney University, Australia

^39^ Network for Improving Critical Care Systems and Training, Colombo, Sri Lanka

^40^ Mahidol Oxford Tropical Medicine Research Unit, Bangkok, Thailand

^41^ NHS Blood and Transplant, London, UK

^42^ Medical Research Institute of New Zealand, Wellington, New Zealand

^43^ UPMC Children’s Hospital of Pittsburgh, Pittsburgh, Pennsylvania, USA

^44^ Department of Critical Care Medicine, University of Pittsburgh School of Medicine, Pittsburgh, Pennsylvania, USA

^45^ NHSBT Clinical Trials Unit, Bristol, UK

^46^ Department of Healthcare Quality Assessment, Graduate School of Medicine, The University of Tokyo, Japan

^47^ Université de Sherbrooke, Sherbrooke, Québec, Canada

^48^ Cardiac Intensive Care Unit, Peter Munk Cardiac Centre, University Health Network, Interdepartmental Division of Critical Care Medicine, University of Toronto, Toronto, Canada

^49^ The Clinical Research Investigation and Systems Modeling of Acute Illness (CRISMA) Center, Department of Critical Care Medicine, University of Pittsburgh School of Medicine, Pittsburgh, Pennsylvania, USA

^50^ School of Medicine and Pharmacology, University of Western Australia, Crawley, Western Australia, Australia

^51^ NHS Blood and Transplant, Barnsley, UK

^52^ Interdepartmental Division of Critical Care, University of Toronto, Toronto, Ontario, Canada

^53^ Centre for Experimental Medicine, School of Medicine, Dentistry and Biomedical Sciences, Queen’s University Belfast, Belfast, UK

^54^ Cardiothoracic and Vascular Intensive Care Unit, Auckland City Hospital, Auckland, New Zealand

^55^ The Medical Research Institute of New Zealand, Wellington, New Zealand

^56^ NHS Blood and Transplant,500 North Bristol Park, Filton, Bristol, UK

^57^ The UPMC Health System Office of Healthcare Innovation, Pittsburgh, Pennsylvania, USA

^58^ Clinical Trials Unit, Intensive Care National Audit & Research Centre (ICNARC), London, UK

^59^ University of British Columbia School of Medicine, Vancouver, Canada

^60^ Department of Anesthesia and Intensive Care, St Vincent’s University Hospital, Dublin, Ireland

^61^ School of Medicine and Medical Sciences, University College Dublin, Dublin, Ireland

^62^ Department of Intensive Care, Alfred Health, Melbourne, Victoria, Australia

^63^ School of Nursing, University of Auckland, Auckland, New Zealand

^64^ Scottish National Blood Transfusion Service, Edinburgh, Scotland, UK

^65^ The Peter Doherty Institute for Infection and Immunity, The University of Melbourne, Melbourne, Australia

^66^ Universidad de La Sabana, Chia, Colombia

^67^ Clinica Universidad de La Sabana, Chia, Colombia

^68^ Héma-Québec, Montréal, Québec, Canada

^69^ Division of Hematology and Oncology, Department of Pediatrics, CHU Sainte-Justine, Montréal, Québec, Canada

^70^ Department of Pediatrics, Université de Montréal, Montréal, Québec, Canada

^71^ Department of Emergency and Critical Care Medicine, St. Marianna University School of Medicine Yokohama City Seibu Hospital, Yokohama, Japan

^72^ Department of Critical Care Medicine, Hospital Israelita Albert Einstein, São Paulo, Brazil

^73^ Department of Pathology, University of Pittsburgh School of Medicine, Pittsburgh, Pennsylvania, USA

^74^ Radboud Institute for Molecular Life Sciences, Radboud University Medical Center, Nijmegen, Netherlands

^75^ University of Edinburgh, Scotland, UK

^76^ Department of Emergency Medicine, Harbor-UCLA Medical Center, Torrance, California, USA

^77^ Department of Emergency Medicine, David Geffen School of Medicine at UCLA, Los Angeles, California, USA

^78^ University Division of Anaesthesia, Addenbrooke’s Hospital Cambridge, Cambridge, UK

^79^ Department of Critical Care Medicine, Auckland City Hospital, Auckland, New Zealand

^80^ Department of Medicine, Critical Care and Hematology/Medical Oncology, University of Manitoba, Winnipeg, Manitoba, Canada

^81^ Guy’s and St Thomas’ NHS Foundation Trust, ICU support Offices,1 st Floor, East Wing, St Thomas’ Hospital, SE17 EH, UK

## Author contributions

To be generated from author contribution forms.

## The REMAP-CAP Investigators

See attachment ‘REMAP-CAP Investigators’.

## Data Sharing Statement

See Data Sharing Statement.

## Acknowledgement

The views expressed in this publication are those of the author(s) and not necessarily those of the NHS, the UK National Institute for Health Research or the Department of Health and Social Care.

We are grateful to the NIHR Clinical Research Network (UK), UPMC Health System Health Services Division (US), and the Direction de la Recherche Clinique et de l’Innovation de l’AP-HP (France) for their support of participant recruitment. ACG is funded by an NIHR Research Professorship (RP-2015-06-18). AFT is the chairholder of a Canada Research Chair. ZM is supported by an NHMRC Emerging Leader Fellowship (APP194811). MSH is funded by an NIHR Clinician Scientist Fellowship (CS-2016-16-011).

## Summary of Supplements

1. **Trial Protocol and SAP Documents**
2. **Supplementary Appendix**

## References

1. Gupta A, Madhavan MV, Sehgal K, et al. Extrapulmonary manifestations of COVID-19. Nature Medicine 2020;26:1017–32.

2. Rochwerg B, Siemieniuk RA, Agoritsas T, et al. A living WHO guideline on drugs for covid-19. BMJ 2020:m3379.

3. Angus DC, Derde L, Al-Beidh F, et al. Effect of Hydrocortisone on Mortality and Organ Support in Patients With Severe COVID-19. JAMA 2020;324:1317.

4. Interleukin-6 Receptor Antagonists in Critically Ill Patients with Covid-19. New England Journal of Medicine 2021;384:1491–502.

5. Sterne JAC, Murthy S, Diaz JV, et al. Association Between Administration of Systemic Corticosteroids and Mortality Among Critically Ill Patients With COVID-19. JAMA 2020;324:1330.

6. Bloch EM, Shoham S, Casadevall A, et al. Deployment of convalescent plasma for the prevention and treatment of COVID-19. J Clin Invest 2020.

7. Luke TC, Kilbane EM, Jackson JL, Hoffman SL. Meta-Analysis: Convalescent Blood Products for Spanish Influenza Pneumonia: A Future H5N1 Treatment? Annals of Internal Medicine 2006;145:599.

8. Mair-Jenkins J, Saavedra-Campos M, Baillie JK, et al. The Effectiveness of Convalescent Plasma and Hyperimmune Immunoglobulin for the Treatment of Severe Acute Respiratory Infections of Viral Etiology: A Systematic Review and Exploratory Meta-analysis. Journal of Infectious Diseases 2015;211:80–90.

9. Casadevall A, Pirofski L-A. The convalescent sera option for containing COVID-19. Journal of Clinical Investigation 2020.

10. Gutmann C, Takov K, Burnap SA, et al. SARS-CoV-2 RNAemia and proteomic trajectories inform prognostication in COVID-19 patients admitted to intensive care. Nature Communications 2021;12.

11. Bermejo-Martin JF, González-Rivera M, Almansa R, et al. Viral RNA load in plasma is associated with critical illness and a dysregulated host response in COVID-19. Critical Care 2020;24.

12. Casadevall A, Dragotakes Q, Johnson PW, et al. Convalescent plasma use in the United States was inversely correlated with COVID-19 mortality. eLife 2021;10.

13. Abani O, Abbas A, Abbas F, et al. Convalescent plasma in patients admitted to hospital with COVID-19 (RECOVERY): a randomised controlled, open-label, platform trial. The Lancet 2021.

14. Libster R, Pérez Marc G, Wappner D, et al. Early High-Titer Plasma Therapy to Prevent Severe Covid-19 in Older Adults. New England Journal of Medicine 2021.

15. Piechotta V, Iannizzi C, Chai KL, et al. Convalescent plasma or hyperimmune immunoglobulin for people with COVID-19: a living systematic review. Cochrane Database of Systematic Reviews 2021.

16. Agarwal A, Mukherjee A, Kumar G, Chatterjee P, Bhatnagar T, Malhotra P. Convalescent plasma in the management of moderate covid-19 in adults in India: open label phase II multicentre randomised controlled trial (PLACID Trial). BMJ 2020:m3939.

17. Li L, Zhang W, Hu Y, et al. Effect of Convalescent Plasma Therapy on Time to Clinical Improvement in Patients With Severe and Life-threatening COVID-19. JAMA 2020.

18. Simonovich VA, Burgos Pratx LD, Scibona P, et al. A Randomized Trial of Convalescent Plasma in Covid-19 Severe Pneumonia. New England Journal of Medicine 2020.

19. Xia X, Li K, Wu L, et al. Improved clinical symptoms and mortality among patients with severe or critical COVID-19 after convalescent plasma transfusion. Blood 2020;136:755–9.

20. Angus DC, Berry S, Lewis RJ, et al. The REMAP-CAP (Randomized Embedded Multifactorial Adaptive Platform for Community-acquired Pneumonia) Study. Rationale and Design. Ann Am Thorac Soc 2020;17:879–91.

21. Zarychanski R. Therapeutic Anticoagulation in Critically Ill Patients with Covid-19 – Preliminary Report. Cold Spring Harbor Laboratory; 2021.

22. Lawler PR, Goligher EC, Berger JS, et al. Therapeutic Anticoagulation in Non-Critically Ill Patients with Covid-19. Cold Spring Harbor Laboratory; 2021.

23. (WHO) WHO. COVID-19 Clinical management. 2021.

24. RECOVERY trial closes recruitment to convalescent plasma treatment for patients hospitalised with COVID-19. 2021. at https://www.recoverytrial.net/news/statement-from-the-recovery-trial-chief-investigators-15-january-2021-recovery-trial-closes-recruitment-to-convalescent-plasma-treatment-for-patients-hospitalised-with-covid-19.)

25. Dexamethasone in Hospitalized Patients with Covid-19 — Preliminary Report. New England Journal of Medicine 2020.

26. Horby PW, Estcourt L, Peto L, et al. Convalescent plasma in patients admitted to hospital with COVID-19 (RECOVERY): a randomised, controlled, open-label, platform trial. Cold Spring Harbor Laboratory; 2021.

27. Laing AG, Lorenc A, Del Molino Del Barrio I, et al. A dynamic COVID-19 immune signature includes associations with poor prognosis. Nature Medicine 2020;26:1623–35.

28. Mathew D, Giles JR, Baxter AE, et al. Deep immune profiling of COVID-19 patients reveals distinct immunotypes with therapeutic implications. Science 2020;369:eabc8511.

29. Ratcliff J, Nguyen D, Fish M, et al. Virological and serological characterization of critically ill patients with COVID-19 in the UK: Interactions of viral load, antibody status and B.1.1.7 variant infection. The Journal of Infectious Diseases 2021.

30. FDA. Convalescent Plasma EUA Letter of Authorization March 9, 2021. 2021.

