## Supplementary Appendix for "Convalescent Plasma in Critically ill Patients with Covid-19"

#### The REMAP-CAP Investigators

#### Supplementary Appendix

### 1. REMAP-CAP investigators and collaborators

#### REMAP-CAP Trial Investigators & Collaborators

**International Trial Steering Committee:**

Farah Al-Beidh, Derek Angus, Djillali Annane, Yaseen Arabi, Abigail Beane, Wilma van Bentum-Puijk, Scott Berry, Zahra Bhimani, Marc Bonten, Charlotte Bradbury, Frank Brunkhorst, Meredith Buxton, Allen Cheng, Matthew Cove, Menno de Jong, Lennie Derde, Lise Estcourt, Herman Goossens, Anthony Gordon, Cameron Green, Rashan Haniffa, Nao Ichihara, Francois Lamontagne, Patrick Lawler, Edward Litton, John Marshall, Colin McArthur, Daniel McAuley, Shay McGuinness, Bryan McVerry, Stephanie Montgomery, Paul Mouncey, Srinivas Murthy, Alistair Nichol, Rachael Parke, Jane Parker, Luis Felipe Reyes, Kathryn Rowan, Hiroki Saito, Marlene Santos, Christopher Seymour, Manu Shankar-Hari, Alexis Turgeon, Anne Turner, Frank van de Veerdonk, Steve Webb (Chair), Ryan Zarychanski

**Regional Management Committees**

***Australia and New Zealand***

Yaseen Arabi, Lewis Campbell, Allen Cheng, Lennie Derde, Andrew Forbes, David Gattas, Cameron Green, Stephane Heritier, Peter Kruger, Edward Litton, Colin McArthur (Deputy Executive Director), Shay McGuinness (Chair), Alistair Nichol, Rachael Parke, Jane Parker, Sandra Peake, Jeffrey Presneill, Ian Seppelt, Tony Trapani, Anne Turner, Steve Webb (Executive Director), Paul Young

***Canadian Regional Management Committee***

Zahra Bhimani, Brian Cuthbertson, Rob Fowler, Francois Lamontagne, John Marshall (Executive Director), Venika Manoharan, Srinivas Murthy (Deputy Executive Director), Marlene Santos, Alexis Turgeon, Ryan Zarychanski

***CRIT Care Asia (CCA) Regional Management Committee***

Diptesh Aryal, Abi Beane (Chair), Arjen M Dondrop, Cameron Green, Rashan Haniffa (Executive Director), Madiha Hashmi, Issrah Jawad, Deva Jayakumar, John Marshall, Colin McArthur, Srinivas Murthy, Timo Tolppa, Vanessa Singh, Steve Webb

***European Regional Management Committee***

Farah Al-Beidh, Derek Angus, Djillali Annane, Wilma van Bentum-Puijk, Scott Berry, Marc Bonten (Executive Director and Chair), Nicole Brillinger, Frank Brunkhorst, Maurizio Cecconi, Lennie Derde, Stephan Ermann, Bruno Francois, Herman Goossens, Anthony Gordon, Cameron Green, Sebastiaan Hullegie, Rene Markgraff, Colin McArthur, Paul Mouncey, Alistair Nichol, Mathias Pletz, Pedro Povoa, Gernot Rohde, Kathryn Rowan, Steve Webb

***United States Regional Management Committee***

Brian Alexander, Derek Angus (Executive Director), Kim Basile, Meredith Buxton (Chair), Timothy Girard, Christopher Horvat, David Huang, Kelsey Linstrum, Florian Mayr, Bryan McVerry, Stephanie Montgomery, Christopher Seymour

**Regional Coordinating Centers**

***Australia, CCA region, and Saudi Arabia:*** The Australia and New Zealand Intensive Care Research Centre (ANZIC-RC), Monash University

***Canada:*** St. Michael's Hospital, Unity Health Toronto (central regional coordination) and CHU de Québec-Université Laval Hospital, Université Laval (Immunoglobulin Domain regional coordination)

***Europe:*** University Medical Center Utrecht (UMCU)

***Ireland:*** Irish Critical Care Clinical Trials Network, University College Dublin Research Centre, St. Vincent's Hospital

***New Zealand:*** The Medical Research Institute of New Zealand (MRINZ)

***United Kingdom:*** Intensive Care National Audit and Research Centre (ICNARC), and Imperial College London

***United States:*** Global Coalition for Adaptive Research (GCAR), and University of Pittsburgh Medical Center

**CRIT Care Asia (CCA):** NICS MORU.

**Domain-Specific Working Groups**

***Antibiotic and Macrolide Duration Domain-Specific Working Group***

Richard Beasley, Marc Bonten, Allen Cheng (Chair), Nick Daneman, Lennie Derde, Robert Fowler, David Gattas, Anthony Gordon, Cameron Green, Peter Kruger, Colin McArthur, Steve McGloughlin, Susan Morpeth, Srinivas Murthy, Alistair Nichol, Mathias Pletz, David Paterson, Gernot Rohde, Steve Webb

***Corticosteroid Domain-Specific Working Group***

Derek Angus (Chair), Wilma van Bentum-Puijk, Lennie Derde, Anthony Gordon, Sebastiaan Hullegie, Peter Kruger, Edward Litton, John Marshall, Colin McArthur, Srinivas Murthy, Alistair Nichol, Bala Venkatesh, Steve Webb

***Influenza Antiviral Domain-Specific Working Group***

Derek Angus, Scott Berry, Marc Bonten, Allen Cheng, Lennie Derde, Herman Goossens, Sebastiaan Hullegie, Menno de Jong, John Marshall, Colin McArthur, Srinivas Murthy (Chair), Tim Uyeki, Steve Webb

***COVID-19 Antiviral Domain-Specific Working Group***

Derek Angus, Yaseen Arabi (Chair), Kenneth Baillie, Richard Beasley, Scott Berry, Marc Bonten, Allen Cheng, Menno de Jong, Lennie Derde, Eamon Duffy, Rob Fowler, Herman Goossens, Anthony Gordon, Cameron Green, Thomas Hills, Colin McArthur, Susan Morpeth, Srinivas Murthy, Alistair Nichol, Katrina Orr, Rachael Parke, Jane Parker, Asad Patanwala, Kathryn Rowan, Steve Tong, Tim Uyeki, Frank van de Veerdonk, Steve Webb

**COVID-19 Immune Modulation Domain-Specific Working Group**

Derek Angus, Yaseen Arabi, Kenneth Baillie, Richard Beasley, Scott Berry, Marc Bonten, Frank Brunkhorst, Allen Cheng, Nichola Cooper, Olaf Cremer, Menno de Jong, Lennie Derde (Chair), Eamon Duffy, James Galea, Herman Goossens, Anthony Gordon, Cameron Green, Thomas Hills, Andrew King, Helen Leavis, John Marshall, Florian Mayr, Colin McArthur, Bryan McVerry, Susan Morpeth, Srinivas Murthy, Mihai Netea, Alistair Nichol, Kayode Ogungbenro, Katrina Orr, Jane Parker, Asad Patawala, Ville Pettilä (Deputy Chair), Emma Rademaker, Kathryn Rowan, Manoj Saxena, Christopher Seymour, Wendy Sligl, Steven Tong, Tim Uyeki, Frank van de Veerdonk, Steve Webb, Taryn Youngstein

**COVID-19 Immune Modulation -2 Domain-Specific Working Group**

Derek Angus, Scott Berry, Lennie Derde, Cameron Green, David Huang, Florian Mayr, Bryan McVerry, Stephanie Montgomery, Christopher W. Seymour (Chair), Steve Webb

***Therapeutic Anticoagulation Domain-Specific Working Group***

Derek Angus, Diptesh Aryal, Scott Berry, Shilesh Bihari, Charlotte Bradbury, Marc Carrier, Dean Fergusson, Robert Fowler, Ewan Goligher (Deputy Chair), Anthony Gordon, Christopher Horvat, David Huang, Beverley Hunt, Devachandran Jayakumar, Anand Kumar, Mike Laffan, Patrick Lawler, Sylvain Lother, Colin McArthur,

Bryan McVerry, John Marshall, Saskia Middeldopr, Zoe McQuilten, Matthew Neal, Alistair Nichol, Christopher Seymour, Roger Schutgens, Simon Stanworth, Alexis Turgeon, Steve Webb, Ryan Zarychanski (Chair)

***Vitamin C Domain-Specific Working Group***

Neill Adhikari (Chair), Derek Angus, Djillali Annane, Matthew Anstey, Yaseen Arabi, Scott Berry, Emily Brant, Angelique de Man, Lennie Derde, Anthony Gordon, Cameron Green, David Huang, Francois Lamonagne (Chair), Edward Litton, John Marshall, Marie-Helene Masse, Colin McArthur, Shay McGuinness, Paul Mouncey, Srinivas Murthy, Rachael Parke, Alistair Nichol, Tony Trapani, Andrew Udy, Steve Webb

***COVID-19 Immunoglobulin Domain-Specific Working Group***

Derek Angus, Donald Arnold, Phillipe Begin, Scott Berry, Richard Charlewood, Michael Chassé, Mark Coyne, Jamie Cooper, James Daly, Lise Estcourt (Chair, UK lead), Dean Fergusson, Anthony Gordon, Iain Gosbell, Heli Harvala-Simmonds, Tom Hills (New Zealand lead), Christopher Horvat, David Huang, Sheila MacLennan, John Marshall, Colin McArthur (New Zealand lead), Bryan McVerry (USA lead), David Menon, Susan Morpeth, Paul Mouncey, Srinivas Murthy, John McDyer, Zoe McQuilten (Australia lead), Alistair Nichol (Ireland lead), Nicole Pridee, David Roberts, Kathryn Rowan, Christopher Seymour, Manu Shankar-Hari (Co-chair, UK lead), Helen Thomas, Alan Tinmouth, Darrell Triulzi, Alexis Turgeon (Canada lead), Tim Walsh, Steve Webb, Erica Wood, Ryan Zarychanski (Canada lead)

***Simvastatin Domain-Specific Working Group***

Derek Angus, Yaseen Arabi, Abi Beane, Carolyn Calfee, Anthony Gordon, Cameron Green, Rashan Haniffa, Deva Jayakumar, Peter Kruger, Patrick Lawler, Edward Litton, Colin McArthur, Daniel McAuley (Chair), Bryan McVerry, Matthew Neal, Alistair Nichol, Cecilia O'Kane, Murali Shyamsundar, Pratik Sinha, Taylor Thompson, Steve Webb, Ian Young

***Antiplatelet Domain-Specific Working Group***

Derek Angus, Scott Berry, Shailesh Bihari, Charlotte Bradbury (Chair), Marc Carrier, Timothy Girard, Ewan Goligher, Anthony Gordon, Ghady Haidar, Christopher Horvat, David Huang, Beverley Hunt, Anand Kumar, Patrick Lawler, Colin McArthur, Bryan McVerry, John Marshall, Zoe McQuilten, Matthew Neal, Alistair Nichol, Christopher Seymour, Simon Stanworth, Steve Webb, Alexandra Weissman, Ryan Zarychanski

***Mechanical Ventilation Domain***

Derek Angus, Wilma van Bentum-Puijk, Lewis Campbell, Lennie Derde, Niall Ferguson, Timothy Girard, Ewan Goligher, Anthony Gordon, Cameron Green, Carol Hodgson, Peter Kruger, John Laffey, Edward Litton, John Marshall, Colin McArthur, Daniel McAuley, Shay McGuinness, Alistair Nichol (Chair) Neil Orford, Kathryn Rowan, Ary Neto, Steve Webb

***ACE-2 RAS Domain***

Rebecca Baron, Lennie Derde, Slava Epelman, Claudia Frankfurter, David Gattas, Frank Gommans, Anthony Gordon, Rashan Haniffa, David Huang, Edy Kim, Francois Lamontagne, Patrick Lawler (Chair), David Leaf, John Marshall, Colin McArthur, Bryan McVerry, Daniel McAuley, Muthiah Vaduganathan, Roland van Kimmenade, Frank van de Veerdonk, Steve Webb

**Statistical Analysis Committee**

Michelle Detry, Mark Fitzgerald, Roger J Lewis (Chair), Anna McGlothlin, Ashish Sanil, Christina Saunders

**Statistical Design Team**

Lindsay Berry, Scott Berry, Elizabeth Lorenzi

**Project Management**

***Australia and Saudi Arabia:*** Jane Parker, Vanessa Singh, Claire Zammit

***Canada:*** Central regional management: Zahra Bhimani, Marlene Santos; Immunoglobulin Domain regional management: David Bellemare, Olivier Costerousse, Rana Daher

***CCA:*** Abi Beane, Rashan Haniffa, Timo Tolppa

***Europe:*** Wilma van Bentum Puijk, Wietske Bouwman, Radhika Ganpat, Erika Groenveld, Denise van Hout, Yara Mangindaan, Clementina Okundaye, Lorraine Parker, Svenja Peters, Ilse Rietveld, Linda Rikkert, Kik Raymakers, Irma Scheepstra-Beukers, Albertine Smit,

***Germany:*** Nicole Brillinger, Rene Markgraf

***Global:*** Cameron Green

***Ireland:*** Kate Ainscough, Kathy Brickell, Peter Doran

***New Zealand:*** Anne Turner

***United Kingdom:*** Farah Al-Beidh, Aisha Anjum, Janis-Best Lane, Elizabeth Fagbodun, Lorna Miller, Paul Mouncey, Karen Parry-Billings, Sam Peters, Alvin Richards-Belle, Michelle Saull, Stefan Sprinckmoller, Daisy Wiley

***United States of America:*** Kim Basile, Meredith Buxton, Kelsey Linstrum, Stephanie Montgomery, Tracey Roberts, Renee Wunderely

**<u>Data and Safety Monitoring Board</u>**

Julian Bion, Jason Connor, (Deputy Chair), Simon Gates, Victoria Manax (Chair), Tom van der Poll, John Reynolds

**<u>Database Providers</u>**

***Research Online:***

Marloes van Beurden, Evelien Effelaar, Joost Schotsman,

***Spinnaker Software:***

Craig Boyd, Cain Harland, Audrey Shearer, Jess Wren

***University of Pittsburgh Medical Center***

Giles Clermont, William Garrard, Christopher Horvat, Kyle Kalchthaler, Andrew King, Daniel Ricketts, Salim Malakouti, Oscar Marroquin, Edvin Music, Kevin Quinn

**NICS MORU: on behalf of CCA;**

Udara Attanayaka, Abi Beane, Sri Darshana, Rashan Haniffa, Pramodya Ishani, Issrah Jawad, Upulee Pabasara, Timo Tolppa, Ishara Udayanga.

**<u>Site Investigators and Research Coordinators</u>**

**Australia:**

*The Alfred Hospital*: Andrew Udy, Phoebe McCracken, Meredith Young, Jasmin Board, Emma Martin;

*Ballarat Health Services*: Khaled El-Khawas, Angus Richardson, Dianne Hill, Robert J Commons, Hussam Abdelkharim;

*Bendigo Hospital*: Cameron Knott, Julie Smith, Catherine Boschert;

*Caboolture Hospital*: Julia Affleck, Yogesh Apte, Umesh Subbanna, Roland Bartholdy, Thuy Frakking;

*Campbelltown Hospital*: Karuna Keat, Deepak Bhonagiri, Ritesh Sanghavi, Jodie Nema, Megan Ford;

*Canberra Hospital*: Harshel G. Parikh, Bronwyn Avard, Mary Nourse;

*Concord Repatriation General Hospital*: Winston Cheung, Mark Kol, Helen Wong, Asim Shah, Atul Wagh;

*Eastern Health (Box Hill, Maroondah & Angliss Hospitals)*: Joanna Simpson, Graeme Duke, Peter Chan, Brittney Carter, Stephanie Hunter;

*Flinders Medical Centre*: Shailesh Bihari, Russell D Laver, Tapaswi Shrestha, Xia Jin;

*Fiona Stanley Hospital*: Edward Litton, Adrian Regli, Susan Pellicano, Annamaria Palermo, Ege Eroglu;

*Footscray Hospital*: Craig French, Samantha Bates, Miriam Towns, Yang Yang, Forbes McGain;

*Gold Coast University Hospital*: James McCullough, Mandy Tallott;

*John Hunter Hospital*: Nikhil Kumar, Rakshit Panwar, Gail Brinkerhoff, Cassandra Koppen, Federica Cazzola;

*Launceston General Hospital*: Matthew Brain; Sarah Mineall;

*Lyell McEwin Hospital*: Roy Fischer, Vishwanath Biradar, Natalie Soar;

*Logan Hospital*: Hayden White, Kristen Estensen, Lynette Morrison, Joanne Sutton, Melanie Cooper;

*Monash Health (Monash Medical Centre, Dandenong Hospital & Casey Hospital):* Yahya Shehabi, Wisam Al-Bassam, Amanda Hulley; Umesh Kadam, Kushaharan Sathianathan;

*Nepean Hospital*: Ian Seppelt, Christina Whitehead, Julie Lowrey, Rebecca Gresham, Kristy Masters;

*Princess Alexandra Hospital*: Peter Kruger, James Walsham, Mr Jason Meyer, Meg Harward, Ellen Venz;

*The Prince Charles Hospital*: Kara Brady, Cassandra Vale, Kiran Shekar, Jayshree Lavana, Dinesh Parmar;

*The Queen Elizabeth Hospital*: Sandra Peake, Patricia Williams, Catherine Kurenda;

*Rockhampton Hospital*: Helen Miles, Antony Attokaran;

*Royal Adelaide Hospital*: Samuel Gluck, Stephanie O'Connor, Marianne Chapman, Kathleen Glasby;

*Royal Darwin Hospital*: Lewis Campbell, Kirsty Smyth, Margaret Phillips;

*Royal Melbourne Hospital*: Jeffrey Presneill, Deborah Barge, Kathleen Byrne, Alana Driscoll, Louise Fortune;

*Royal North Shore Hospital*: Pierre Janin, Elizabeth Yarad, Frances Bass, Naomi Hammond, Anne O'Connor;

*Royal Perth Hospital*: Sharon Waterson, Steve Webb, Robert McNamara;

*Royal Prince Alfred Hospital*: David Gattas, Heidi Buhr, Jennifer Coles;

*Sir Charles Gardiner Hospital*: Sacha Schweikert, Bradley Wibrow, Matthew Anstey, Rashmi Rauniyar;

*St George Hospital*: Kush Deshpande, Pam Konecny, Jennene Miller, Adeline Kintono, Raymond Tung

*St. John of God Midland Public and Private Hospitals*: Ed Fysh, Ashlish Dawda, Bhaumik Mevavala;

*St. John of God Hospital, Murdoch*: Annamaria Palermo, Adrian Regli, Bart De Keulenaer;

*St. John of God Hospital, Subiaco*: Ed Litton, Janet Ferrier;

*St. Vincent's Hospital (NSW)*: Priya Nair, Hergen Buscher, Claire Reynolds, Sally Newman;

*St. Vincent's Hospital (VIC)*: John Santamaria, Leanne Barbazza, Jennifer Homes, Roger Smith;

*Sunshine Coast University Hospital*: Peter Garrett, Lauren Murray, Jane Brailsford, Loretta Forbes, Teena Maguire;

*Sunshine Hospital*: Craig French, Gerard Fennessy, John Mulder, Rebecca Morgan, Rebecca McEldrew;

*The Sutherland Hospital*: Anas Naeem, Laura Fagan, Emily Ryan;

*Toowoomba Hospital*: Vasanth Mariappa, Judith Smith;

*University Hospital Geelong*: Scott Simpson, Matthew Maiden, Allison Bone, Michelle Horton, Tania Salerno;

*Wollongong Hospital*: Martin Sterba, Wenli Geng;

***Belgium:***

*Ghent University Hospital*: Pieter Depuydt, Jan De Waele, Liesbet De Bus, Jan Fierens, Stephanie Bracke, Joris Vermassen, Daisy Vermeiren;

***Canada*:**

*Brantford General Hospital:* Brenda Reeve, William Dechert;

Institut universitaire de cardiologie et de pneumologie de Québec: Francois Lellouche, Patricia Lizotte

*Centre Hospitalier de l'Universite de Montreal:* Michaël Chassé, François Martin Carrier, Dounia Boumahni, Fatna Benettaib, Ali Ghamraoui;

*CHU de Québec – Université Laval*: Alexis Turgeon, David Bellemare, Marie-Claude Boulanger, Ève Cloutier, Olivier Costerousse, Rana Daher, François Lauzier, Charles Francoeur;

*Centre Hospitalier Universitaire de Sherbrooke:* François Lamontagne, Frédérick D'Aragon, Elaine Carbonneau, Julie Leblond;

Grace Hospital: Gloria Vazquez-Grande, Nicole Marten

Grand River Hospital (Kitchener): Theresa Liu, Atif Siddiqui

*Health Sciences Centre, Winnipeg:* Ryan Zarychanski, Gloria Vazquez-Grande, Nicole Marten, RN, Maggie Wilson;

*Hôpital du Sacré Coeur de Montréal:* Martin Albert, Karim Serri, Alexandros Cavayas, Mathilde Duplaix, Virginie Williams;

*Juravinski Hospital:* Bram Rochwerg, Tim Karachi, Simon Oczkowski, John Centofanti, Tina Millen

McGill University Health Centre: Josie Campisi, Kosar Khwaja,

*Niagara Health (St. Catherine's Hospital):* Erick Duan, Jennifer Tsang, Lisa Patterson;

*Regina General Hospital*: Eric Sy, Chiraag Gupta, Sandy Kassir, Jonathan Mailman, Stephen Lee

*Royal Alexandra Hospital*: Demetrios Kutsogiannis, Patricia Thompson

*Sunnybrook Health Sciences Centre*: Rob Fowler, Neill Adhikari, Maneesha Kamra, Nicole Marinoff

*St. Boniface General Hospital*: Ryan Zarychanski, Nicole Marten

*St. Joseph's Healthcare Hamilton*: Deborah Cook, Frances Clarke

*St. Mary's General Hospital (Kitchener)*: Rebecca Kruisselbrink, Atif Siddiqui

*St. Michael's Hospital*: John Marshall, Laurent Brochard, MD, Karen Burns, MD, Gyan Sandhu, Imrana Khalid;

*The Ottawa Hospital:* Shane English, Irene Watpool, Rebecca Porteous, Sydney Miezitis, Lauralyn McIntyre;

*University Health Network:* Elizabeth Wilcox, Lorenzo del Sorbo, Hesham Abdelhady, Tina Romagnuolo

*University of Alberta:* Wendy Sligl, Nadia Baig, Oleksa Rewa, Sean Bagshaw

*William Osler Health System:* Alexandra Binnie, Elizabeth Powell, Alexandra McMillan, Tracy Luk, Noah Aref

**CCA:**

India Apollo Speciality Hospital - OMR, Chennai: Devachandran Jayakumar, Suresh Babu;

Apollo Main Hospital, Chennai: C Vignesh, Augustian James

Apollo Speciality Vanagaram, Vanagaram, Chennai: R Ebenezer, S Krishnamurthy, Lakshmi Ranganathan, Manisha

Nepal Grande International Hospital : Sushil Khanal, Sameena Amatya;

HAMS Hospital: Hem Raj Paneru, Sabin Koirala, Pratibha Paudel;

Nepal Mediciti Hospital: Diptesh Aryal, Kanchan Koirala, Namrata Rai, Subekshya Luitel;

Tribhuvan University Teaching Hospital : Hem Raj Paneru, Binita Bhattarai;

Pakistan (CCA):

Ziauddin Hospital Clifton Campus: Madiha Hashmi; Ashok Panjwani; Zulfiqar Ali Umrani, Shoaib Siddiq; Mohiuddin Shaikh; National Institute of Cardiovascular Diseases Pakistan: Nawal Salahuddin, Sobia Masood;

***Croatia:***

*General Hospital Pozega*: Zdravko Andric, Sabina Cviljevic, Renata Đimoti, Marija Zapalac, Gordan *Mirković;*

*University Hospital of Infectious Diseases "Dr Fran Milhajevid"*: Bruno Baršić, Marko Kutleša, Viktor Kotarski;

*University Hospital of Zagreb*: Ana Vujaklija Brajković, Jakša Babel, Helena Sever, Lidija Dragija, Ira Kušan;

***Finland:***

*Helsinki University Hospital*: Suvi Vaara, Leena Pettilä, Jonna Heinonen, Ville Pettilä;

Tampere University Hospital: Anne Kuitunen, Sari Karlsson, Annukka Vahtera, Heikki Kiiski, Sanna Ristimäki;

***France:***

*Ambroise Pare Hospital*: Amine Azaiz, Cyril Charron, Mathieu Godement, Guillaume Geri, Antoine Vieillard-Baron;

*Centre Hospitalier de Melun*: Franck Pourcine, Mehran Monchi;

*Centre Hospitalier Simone Veil, Beauvais*: David Luis, Romain Mercier, Anne Sagnier, Nathalie Verrier, Cecile Caplin, Jack Richecoeu, Daniele Combaux;

*Centre Hospitalier Sud Essonne*: Shidasp Siami, Christelle Aparicio, Sarah Vautier, Asma Jeblaoui, Delphine Lemaire-Brunel;

*Centre Hospitalier Tenon*: Muriel Fartoukh, Laura Courtin, Vincent Labbe, Guillaume Voiriot, Sara Nesrine Salhi;

*Centre Hospitalier Victor Dupouy*: Gaetan Plantefeve, Cécile Leparco, Damien Contou;

CHR d'Orleans: Grégoire Muller, Mai-Anh Nay, Toufik Kamel, Dalila Benzekri, MD, Sophie Jacquier, Isabelle Runge, Armelle Mathonnet, François Barbier, MD, Anne Bretagnol;

*CHRU Tours Hopital Bretonneau*: Emmanuelle Mercier, Delphine Chartier, Charlotte Salmon, Pierre-François Dequin, Denis Garot;

*CHU Dupuytren, Limoges;*

*Hôpital Civil, Hôpitaux Universitaires de Strasbourg;*

*Hôpital de Hautepierre, Hôpitaux Universitaires de Strasbourg*: Francis Schneider, Vincent Castelain, Guillaume Morel, Sylvie L'Hotellier;

Hospital Nord Franche-Comté: Julio Badie, Fernando Daniel Berdaguer, Sylvain Malfroy, Chaouki Mezher, Charlotte Bourgoin, Guy Moneger, Elodie Bouvier;

Lariboisière Hospital: Bruno Megarbane, Sebastian Voicu, Nicolas Deye, Isabelle Malissin, Laetitia Sutterlin, Aymen Mrad, Adrien Pépin Lehalleur, Giulia Naim, Philippe Nguyen, Jean-Michel Ekhérian, Yvonnick Boué, Georgios Sidéris, Dominique Vodovar, Emmanuelle Guérin, Caroline Grant;

Le Mans Hospital: Christophe Guitton, Cédric Darreau, Mickaël Landais, Nicolas Chudeau, Alain Robert, Patrice Tirot, Jean Christophe Callahan, Marjorie Saint Martin, Charlène Le Moal, Rémy Marnai, Marie Hélène Leroyer;

Raymond Poincaré Hospital: Djillali Annane, Pierre Moine, Nicholas Heming, Virginie Maxime, Isabelle Bossard, Tiphaine Barbarin Nicholier, Bernard Clair, David Orlikowski, Rania Bounab, Lilia Abdeladim;

*Vendee Hospital*: Gwenhael Colin, Vanessa Zinzoni, Natacha Maquigneau, Matthieu Henri-Lagarrigue, Caroline Pouplet;

***Germany:***

*Carl-Thiem-Klinikum Cottbus gGmbH*: Jens Soukup, Richard Wetzold, Madlen Löbel, Dr. Ing,  Lisa Starke, Patrick Grimm;

*Charité - Universitätsmedizin Berlin*: André Finn, Gabriele Kreß, Uwe Hoff, Carl Friedrich Hinrichs, Jens Nee;

*Jena University Hospital*: Mathias W. Pletz, Stefan Hagel, Juliane Ankert, Steffi Kolanos, Frank Bloos;

*Klinikum Dortmund gGmbH*: Daniela Nickoleit-Bitzenberger, Bernhard Schaaf, Werner Meermeier, Katharina Prebeg, Harun Said Azzaui, Martin Hower, Klaus-Gerd Brieger, Corinna Elender, Timo Sabelhaus, Ansgar Riepe, Ceren Akamp, Julius Kremling, Daniela Klein, Elke Landsiedel-Mechenbier;

*University Hospital of Leipzig*: Sirak Petros, Kevin Kunz, Bianka Schütze;

*Universitätsklinikum Hamburg-Eppendorf*: Stefan Kluge, Axel Nierhaus, Dominik Jarczak, Kevin Roedl, MD;

*University Hospital of Frankfurt*: Gernot Gerhard Ulrich Rohde, MD, Achim Grünewaldt, MD, Jörg Bojunga, MD;

*University Hospital of Wuerzburg*: Dirk Weismann, Anna Frey; Maria Drayss, M.E. Goebeler, Thomas Flor, Gertrud Fragner, Nadine Wahl, Juliane Totzke, Cyrus Sayehli;

*Vivantes Klinikum Neukölln*: Lorenz Reill, Michael Distler, Astrid Maselli;

***Hungary:***

*Almási Balogh Pál Hospital, Ózd*: János Bélteczki, István Magyar, Ágnes Fazekas, Sándor Kovács, Viktória Szőke;

*Jósa András County Hospital, Nyíregyháza*: Gábor Szigligeti, János Leszkoven;

***Ireland:***

*Beacon Hospital Dublin*: Daniel Collins, Kathy Brickell, Liadain Reid, Michelle Smyth, Patrick Breen, Sandra Spain;

*Beaumont Hospital*: Gerard Curley, Natalie McEvoy, Pierce Geoghegan, Jennifer Clarke;

*Galway University Hospitals*: John Laffey, Bairbre McNicholas, Michael Scully, Siobhan Casey, Maeve Kernan, Aoife Brennan, Ritika Rangan, Riona Tully, Sarah Corbett, Aine McCarthy, Oscar Duffy, David Burke;

*St Vincent's University Hospital, Dublin*: Alistair Nichol, Kathy Brickell, Michelle Smyth, Leanne Hayes, Liadain Reid, Lorna Murphy, Andy Neill, Bryan Reidy, Michael O'Dwyer, Donal Ryan, Kate Ainscough;

***Netherlands:***

*Canisius Wilhelmina Ziekenhuis*: Oscar Hoiting, Marco Peters, Els Rengers, Mirjam Evers, Anton Prinssen;

*Deventer Hospital*: Huub L.A. van den Oever, Arriette Kruisdijk-Gerritsen;

*Jeroen Bosch Ziekenhuis*: Koen Simons, Tamara van Zuylen, Angela Bouman;

*Meander Medisch Centrum*: Laura van Gulik;

*Radboud University Medical Center Nijmegen*: Jeroen Schouten, Peter Pickkers, Noortje Roovers, Margreet Klop-Riehl, Hetty van der Eng;

*UMC Leiden*: Evert de Jonge, Jeanette Wigbers, Michael del Prado;

*UMC Utrecht*: Marc Bonten, Olaf Cremer, Lennie Derde, Diederik van Dijk, Emma Rademaker, Jelle Haitsma Mulier, Anna Linda Peters, Birgit Romberg;

Ziekenhuis Gelderse Vallei: Sjoerd van Bree, Marianne Bouw-Ruiter, Barbara Festen, Fiona van Gelder, Mark van Iperen, Margreet Osinga, Roel Schellaars, Dave Tjan, Ruben van der Wekken, Max Melchers, Arthur van Zanten;

***New Zealand*:**

*Auckland City Hospital, Cardiothoracic and Vascular ICU:* Shay McGuinness, Rachael Parke, Eileen Guilder, Magdalena Butler, Keri-Anne Cowdrey, Melissa Woollett;

*Auckland City Hospital, DCCM:* Colin McArthur, Thomas Hills, Lynette Newby, Yan Chen, Catherine Simmonds, Rachael McConnochie, Caroline O'Connor;

*Christchurch Hospital:* Jay Ritzema Carter, Seton Henderson, Kymbalee Van Der Heyden, Jan Mehrtens, Anna Morris, Stacey Morgan;

*Middlemore Hospital:* Tony Williams, Alex Kazemi, Susan Morpeth, Rima Song, Vivian Lai, Dinuraj Girijadevi;

*North Shore Hospital:* Robert Everitt, Robert Russell, Danielle Hacking;

*Rotorua Hospital:* Ulrike Buehner, Erin Williams;

*Tauranga Hospital:* Troy Browne, Kate Grimwade, Jennifer Goodson, Owen Keet, Owen Callender;

*Waikato Hospital:* Robert Martynoga, Kara Trask, Amelia Butler, PGCert,

*Wellington Hospital:* Paul Young, PhD, Chelsea Young, PGDip, Eden Lesona, Shaanti Olatunji, Leanlove Navarra, Raulle Sol Cruz

*Whangarei Hospital:* Katherine Perry, Ralph Fuchs, Bridget Lambert;

*Taranaki Base Hospital:* Jonathan Albrett, Carolyn Jackson, Simon Kirkham;

***Portugal***:

*Hospital de Abrantes:* Nuno José Teodoro Amaro dos Santos Catorze, Tiago Nuno Alfaro Lima Pereira, Ricardo Manuel Castro Ferreira, Joana Margarida Pereira Sousa Bastos, Teresa Margarida Oliveira Batista;

***Romania***:

*"Dr. Victor Babes" Clinical Hospital of Infectious and Tropical Diseases Bucharest:* Simin Aysel Florescu, Delia Stanciu, Mihaela Florentina Zaharia, Alma Gabriela Kosa, Daniel Codreanu;

***Saudi Arabia***:

*King Abdulaziz Medical City- Riyadh:* Yaseen M Arabi, Eman Al Qasim, Mohamed M Hegazy, Hatim Arishi, Ali Al Amri, Samah Y AlQahtani, Brintha Naidu, Haytham Tlayjeh, Sajid Hussain, Farhan Al Enezi, Sheryl Ann Abdukahil, Lolowa Alswaidan

***Spain***:

*Hospital del Mar:* Rosana Muñoz-Bermúdez, Judith Marin-Corral, Anna Salazar Degracia, Francisco Parrilla Gómez, Maria Isabel Mateo López;

*Reina Sofia University Hospital:* Rafael León López, Jorge Rodriguez, Sheila Cárcel, Rosario Carmona, Carmen de la Fuente, Marina Rodriguez;

***United Kingdom***:

*Aberdeen Royal Infirmary:* Callum Kaye, Amanda Coutts, Lynn MacKay; Transfusion: Julia Lussier, Marion Mathie, Lorraine Jappy and all in the Blood Bank Laboratory.

*Addenbrooke's Hospital:* Charlotte Summers, Petra Polgarova, Neda Farahi, Eleonore Fox; Transfusion: Katherine Philpott, Claire Newsam, Michaela Lewin, Tracy Moore, Harriet Madiyiko, Vikkie Rose,

Stephen Grist, Ruth Smith, Helen Dakers-Black, Monzeer Ibrahim, Dora Foukaneli and the laboratory team.

*Alder Hey Children's NHS Foundation Trust:* Stephen J McWilliam, Daniel B Hawcutt, Laura Rad, Laura O'Malley, Jennifer Whitbread, Dawn Jones, Rachael Dore, Paula Saunderson; Transfusion: Tracey Shackleton, SusanOwens, Janet Fu.

*Alexandra Hospital Redditch:* Olivia Kelsall, Nicholas Cowley, Laura Wild, Jessica Thrush, Hannah Wood, Karen Austin; Transfusion: Camran Khan, Gillian Godding, Emily Murphy, Emma Loxley and the Blood Transfusion laboratory team.

*Altnagelvin Hospital:* Adrian Donnelly, Martin Kelly, Naoise Smyth, Sinéad O'Kane, Declan McClintock, Majella Warnock, Ryan Campbell, Edmund McCallion; Transfusion: Adrian Crawford, Bronagh O'Neill, Mary P McNicholl, Josephine Monaghan, Naomi Smyth, Conor Kelly, Clement Kerlin and Haematology/Blood Transfusion/Central Specimen Reception laboratory teams.

*Antrim Area Hospital:* Paul Johnson, Shirley McKenna, Joanne Hanley, Andrew Currie, Barbara Allen, Clare Mc Goldrick, Moyra Mc Master; Transfusion: C A Henry, B Graham, K Potter and the Blood Transfusion laboratory team.

*Barnet Hospital:* Rajeev Jha, Michael Kalogirou, Christine Ellis, Vinodh Krishnamurthy, Aibhilin O'Connor, Saranya Thurairatnam; Transfusion: Rita Atugonza, Anna Li, Jenny Li, Seble Tekle, Ann-Marie Ellis, Anushka Natarajan, Stephanie Mellin and the Blood Transfusion laboratory team.

*Basildon University Hospital:* Dipak Mukherjee, Agilan Kaliappan, Mark Vertue, Anne Nicholson, Joanne Riches, Gracie Maloney, Lauren Kittridge, Amanda Solesbury, Angelo Ramos; Transfusion: Teresa Green, Maria O'Connell and the Blood Transfusion laboratory team.

*Belfast Health and Social Care Trust (Belfast City Hospital, Mater Infirmorium, Royal Victoria Hospital):* Jon Silversides, Peter McGuigan, Kathryn Ward, Aisling O'Neill, Stephanie Finn, Chris Wright, Jackie Green, Érin Collins; Transfusion: Carol Anne Henry, Phillip Windrum, Bronagh O'Neill, Shonagh Reilly and Blood Transfusion laboratory team.

*Brighton and Sussex University Hospitals Trust:* Barbara Phillips, Laura Oritz-Ruiz de Gordoa; Transfusion: Julie Cole, Laura Humber and the Blood Transfusion laboratory team.

*Calderdale and Huddersfield Foundation Trust:* Jez Pinnell, Matt Robinson, Lisa Gledhill, Tracy Wood; Transfusion: Samantha Kershaw, Purav Desai, Michelle Lake, Helen Senior and the Blood Transfusion laboratory team.

*Cardiff and Vale University Health Board:* Matt Morgan, Jade Cole, Helen Hill, Michelle Davies, Angharad Williams, Emma Thomas, Rhys Davies, Matt Wise; Transfusion: Rebecca Carnegie, Sam McWilliam,

Ann Patterson, Rachel Borrell, Claire-Michelle Neville, Lisa Parkinson, Gregory Andrikopoulos, Samuel Burns and the Blood Transfusion laboratory team.

*Charing Cross Hospital:* David Antcliffe, Maie Templeton, Roceld Rojo, Phoebe Coghlan, Joanna Smee, Gareth Barker; Transfusion: A Rahman, David Johnson, Linda Chapple, Eugenia Nweje, K Sabljak, S Kassa, U Munu, H Dawson, Fiona Regan, Fateha Chowdhury, Vashira Chiroma and Blood Transfusion laboratory teams.

*Chesterfield Royal Hospital:* Euan Mackay, Jon Cort, Amanda Whileman, Thomas Spencer, Nick Spittle, Sarah Beavis, Anand Padmakumar, Katie Dale, Joanne Hawes, Emma Moakes, Rachel Gascoyne, Kelly Pritchard, Lesley Stevenson, Justin Cooke, Karolina Nemeth-Roszpopa

*The Christie NHS Foundation Trust:* Vidya Kasipandian, Amit Patel, Suzanne Allibone, Roman Mary-Genetu; Transfusion: Deborah Seals, Sharon Jackson

*Colchester Hospital:* Mohamed Ramali, Ooi Hc, Alison Ghosh, Rawlings Osagie, Malka Jayasinghe Arachchige, Melissa Hartley; Transfusion: Eleanor Byworth, Mahalakshmi Mohan, Susan Turner, Ana Pereira and the Blood Transfusion laboratory team.

*Countess of Chester Hospital:* Peter Bamford, Andrew Reid, Kathryn Cawley, Maria Faulkner, Charlotte Pickering; Transfusion: Nicola Swarbrick, Louise Hodgkinson and the Blood Transfusion laboratory team

*Croydon University Hospital:* Ashok Sundar Raj, Georgios Tsinaslanidis, Reena Nair Khade, Gloria Nwajei Agha, Rose Nalumansi Sekiwala; Transfusion: Matthew Free, Mercy Mbwembwe, Samantha Conran, Betty Cheung and the Blood Transfusion laboratory team.

*Cumberland Infirmary:* Tim Smith, Chris Brewer, Jane Gregory; Transfusion: John Sutton, Janet Nicholson and the Blood Transfusion laboratory team.

*Darlington Memorial Hospital:* James Limb, Amanda Cowton, Julie O'Brien, Kelly Postlethwaite.

*Derriford Hospital:* Nikitas Nikitas, Colin Wells, Liana Lankester, Helen McMillan; Transfusion: Martin Binney, Caroline Lowe and the Blood Transfusion laboratory team.

*Dorset County Hospital:* Mark Pulletz, Patricia Williams, Jenny Birch, Sophie Wiseman, Sarah Horton;

*East Kent Hospitals (Queen Elizabeth the Queen Mother Hospital):* Ana Alegria, Salah Turki, Tarek Elsefi, Nikki Crisp, Louise Allen; Catherine Lorenzen, Rose Spicer, Benjamin Rayner, Lisa March, Keith Kolsteren, Leigh Boorman, Rebecca Jennings, Hannah Moore, Sarah Lymn, Frances Turner and the Blood Transfusion laboratory team.

*East Lancashire Hospitals NHS Trust (Royal Blackburn Hospital):* Nicholas Truman, Matthew Smith, Sri Chukkambotla, Wendy Goddard, Stephen Duberley, Meherunnisa Khan, Aayesha Kazi; Transfusion:

Lynne Mannion, Mary Sokolowski, Stephen Rigby, T Johnson and Blood Transfusion laboratory team.

*Freeman Hospital and Royal Victoria Infirmary, Newcastle upon Tyne*: Iain J McCullagh, Tom Cairns, Helen Hanson, Bijal Patel, Ian Clement; Transfusion: Alison Muir, Aimi Baird, Scott Rowan-Ferry, Gemma Smithson, Matthew Wilson, Caroline Kennedy, Michelle Evans, Haley Ranton and the Blood Transfusion laboratory team

*Frimley Health NHS Foundation Trust:* Omar Touma, Susan Holland, Christopher Hodge, Holly Taylor, Meera Alderman, Nicky Barnes, Joana Da Rocha, Catherine Smith, Nicole Brooks, Thanuja Weerasinghe, Julie-Ann Sinclair, Yousuf Abusamra, Ronan Doherty, Joanna Cudlipp, Rajeev Singh, Haili Yu, Admad Daebis, Christopher Ng, Sara Kendrick, Anita Saran, Ahmed Makky, Danni Greener, Louise Rowe-Leete, Alexandra Edwards, Yvonne Bland, Rozzie Dolman, Tracy Foster; Transfusion: Nelsonseelan Johnson, Josephine Newanji, Joanne Finden, Carol Cole, Kim East, Arul Kandaswamy and the Blood Transfusion laboratory teams.

*Gateshead Health NHS Trust:* Vanessa Linnett, Amanda Sanderson, Jenny Ritzema, Helen Wild, Rachael Lucas, Yvonne Marriott; Transfusion: Lee Sudlow

*George Eliot Hospital:* Divya Khare, Meredith Pinder, Amitha Gopinath, Thogulava Kannan, Steven Dean, Piyush Vanmali;

*Glan Clwyd Hospital:* Richard Pugh, Richard Lean, Xinyi Qiu, Jeremy Scanlan, Andrew Evans, Gwyneth Davies, Joanne Lewis; Transfusion: Luke Hughes, Emma Hall and the Blood Transfusion laboratory team.

*Glangwili General Hospital:* Yvonna Plesnikova, Ahmed Ben Khoud, Samantha Coetzee;

*Glasgow Royal Infirmary:* Kathryn Puxty, Susanne Cathcart, Dominic Rimmer, Catherine Bagot, Kathryn Scott, Laila Martin; Transfusion: Moira Caldwell, Arlene David and the Transfusion Laboratory team.

*Glenfield Hospital Leicester:* Hakeem Yusuff, Graziella Isgro, Chris Brightling, Michelle Bourne, Michelle Craner, Rebecca Boyles; Transfusion: Amardeep Ghattaoraya, Hafiz Qureshi, Yasin Fozdar, Tri Dinh, Marie Browett, all UHL Blood Transfusion Laboratory Staff.

*Grange University Hospital:* Tamas Szakmany, Shiney Cherian, Gemma Williams, Christie James, Abby Waters; Transfusion: Cheryl Davies, Lorraine Lewis-Prosser, Jennifer Summers and the Transfusion Laboratory team.

*Great Western Hospitals NHS Foundation Trust:* Rachel Prout, Roger Stedman, Louisa Davies, Suzannah Pegler, Lynsey Kyeremeh, Louise Moorhouse; Transfusion: Jassy Uppal, Sally Charlton and the Blood Transfusion team.

*Guy's & St Thomas' NHS Foundation Trust:* Manu Shankar-Hari, Gill Arbane, Marina Marotti, Aneta Bociek, Sara Campos; Transfusion: Tim Maggs, Luke Woodford, Julia Wood, Jamie Jones, Beverley Crane, Saidat Turawa, Cristina Lobato, Charlene Furtado, Rosie O'Dea, Ursula Wood, Dharshana Jeyapalan and the Blood Transfusion laboratory teams.

*Hammersmith Hospital:* Stephen Brett, Sonia Sousa Arias, Rebecca Elin Hall; Transfusion: A Rahman, David Johnson, Linda Chapple, Eugenia Nweje, K Sabljak, S Kassa, U Munu, H Dawson, Fiona Regan, Fateha Chowdhury, Vashira Chiroma and Blood Transfusion laboratory teams.

*Homerton University Hospital NHS Foundation Trust:* Susan Jain, Abhinav Gupta, Catherine Holbrook, Pierre Antoine; Transfusion: Mohammed Elmi, Evodian Fonyonga, Alexander Martinez, and the Blood Transfusion laboratory team.

*James Cook University Hospital:* Jeremy Henning, Stephen Bonner, Keith Hugill, Emanuel Cirstea, Dean Wilkinson, Jessica Jones, Mohammed Nagy Tawfik Altomy; Transfusion: Chris Elliott, Carolyn Carveth-Marshall, and the Blood Transfusion laboratory team.

*James Paget University Hospitals:* Michal Karlikowski, Helen Sutherland, Elva Wilhelmsen, Jane Woods, Julie North; Transfusion: Sarah Parsons, Julie Jackson and the Blood Transfusion laboratory team.

*Kettering General Hospital:* Dhinesh Sundaran, Laszlo Hollos, Anna Williams, Margaret Turns, Joanne Walsh; Transfusion: Jason Frankcam, M Silverstone, A Houston, Emily Rich, Charlotte Little and the Blood Transfusion laboratory team.

*King's College Hospital (Denmark Hill site):* Phil Hopkins, John Smith, Harriet Noble, Kevin O'Reilly, Reena Mehta, Onyee Wong, Esther Makanju, Deepak Rao, Nyma Sikondari, Sian Saha, Ele Corcoran, Evita Pappa, Maeve Cockrell, Clare Donegan, Morteza Balaie; Transfusion: Kenneth Amenyah, Kelly Nwankiti, James Davies, David Veniard, Sue Cole, Philip Gold, Catherine Hawkins and the Blood Transfusion laboratory team.

*Lancashire Teaching Hospitals NHS Foundation Trust:* Shondipon Laha, Mark Verlander, Alexandra Williams, Avinash Kumar Jha; Transfusion: Alan Noyon, Derek Wallbank, Sanchia Baines, Emily Fisher, Karis Treuberg, Brittany Houghton, Elisa Ly and the Blood Transfusion laboratory team.

*Leeds Teaching Hospitals Trust:* Elankumaran Paramasivam, Elizabeth Wilby, Bethan Ogg, Clare Howcroft, Angelique Aspinwall, Sam Charlton, Richard Gould, Deena Mistry, Sidra Awan, Caroline Bedford, Joanne Carr-Wilkinson; Transfusion: Amy Ballinger, Diane Howarth, Alexandra Liversidge, Stephanie Ferguson, Jennifer Rock, Marina Karakantza and the Blood Transfusion laboratory team.

*Leicester General Hospital:* Andrew Hall, Jill Cooke, Caroline Gardiner-Hill, Carolyn Maloney, Nigel Brunskill, Olivia Watchorn, Chloe Hardy; Transfusion: Amardeep Ghattaoraya, Hafiz Qureshi, Yasin Fozdar, Tri Dinh, Marie Browett, all UHL Blood Transfusion Laboratory Staff.

*Leicester Royal Infirmary:* Hafiz R Qureshi, Neil Flint, Sarah Nicholson, Sara Southin, Andrew Nicholson, Amardeep Ghattaoraya; Transfusion: Amardeep Ghattaoraya, Hafiz Qureshi, Yasin Fozdar, Tri Dinh, Marie Browett, all UHL Blood Transfusion Laboratory Staff.

*Lewisham and Greenwich NHS Trust:* Dr Daniel Harding, Sinead O'Halloran, Amy Collins Emma Smith, Estefania Trues;

*Liverpool Foundation Trust Aintree:* Barbara Borgatta, Ian Turner-Bone, Amie Reddy, Laura Wilding; Transfusion: Karen Knowles, Holly Wissett, Rebecca Wright, Cristina Dragomir, Abby Peacock, Joshua Bell, Jan Gorry, Iain Houghton, Paula Burgess and all members of the lab BMS and MLA teams.

*Liverpool Heart and Chest Hospital:* Craig Wilson, Zuhra Surti;

*Luton and Dunstable University Hospital:* Loku Chamara Warnapura, Ronan Agno, Prasannakumari Sathianathan, Deborah Shaw, Nazia Ijaz, Adam Spong, Suganya Sabaretnam, Dean Burns, Eva Lang, Margaret Louise Tate; Transfusion: Dani Fisher, Ellen Strakosch, Charlotte Alford, Mamta Parmar, Noha Gasmalseed, Lena Hunold, Anna Gouveia, Sabiya Khan, Raheem Hussein, Tshinupay Mukwa, Gloria Mmadubuko, Josephine Nnadi, Julie Dijo, Terri-Lisa Jacques- Brown, Jade Tan and Blood Bank team.

*Maidstone and Tunbridge Wells NHS Trust:* David Golden, Miriam Davey, Rebecca Seaman, Alexander Osborne; Transfusion: Robert Reilly, Emma Small, Wendy Bonnert, Francis Ajeneye, Carly Moore, Colin Lawler, Carmel Boyd and the Blood Transfusion laboratory team.

*Manchester Royal Infirmary:* Jonathan Bannard-Smith, Richard Clark, Kathrine Birchall, Joanne Henry, Fiona Pomeroy, Rachael Quayle, Katharine Wylie, Anila Sukuraman, Maya John, Sindhu Sibin; Transfusion: Thomas Trimble, Emma Cooperwaite, Carmel Parker, Sarah Pendlebury, Jayne Peters, S Khan, E Anyanwu, M Evans and the Blood Transfusion laboratory teams.

*Medway Maritime Hospital:* Arystarch Makowski, Beata Misztal, Syeda Haider, Angela Liao, Rebecca Squires; Transfusion: Rachel Nicholas, Sarah Haskins, Jasmine Walker, Alison Davis, Sarah Arnott and the Blood Transfusion laboratory team.

*Milton Keynes University Hospital:* Richard Stewart, Esther Mwaura, Louise Mew, Lynn Wren, Felicity Willams, Sara-Beth Sutherland, Rashmi Rebello; Transfusion: Jasmine Beharry, Terrie Perry, Caroline Lowe, Mohammed Khan, Nikita Jacob and the Blood Transfusion laboratory team.

*Mid & South Essex NHS Foundation Trust:* Aneta Oborska, Abdul Kayani, Selver Kalchko-Veyssal, Rajalakshmi Orath Prabakaran, Bernard Hadebe; Transfusion: Teresa Nicholas, Tina Parker and the Blood Transfusion laboratory team.

*Musgrove Park Hospital:* Richard Innes, Patricia Doble, Libby Graham, Charmaine Shovelton, Tessa Dean; Transfusion: Matthew Barnett, Michelle Davey, Nicolette Heydon, Sandra Harlow and the Blood Transfusion laboratory team

*Nevill Hall Hospital:* Vincent Hamlyn, Nancy Hawkins, Anna Roynon-Reed, Sean Cutler, Sarah Lewis; Transfusion: Cheryl Davies, Jennifer Summers, Lorraine Lewis-Prosser and the Blood Transfusion laboratory team.

*Newham University Hospital:* Juan Martin Lazaro, Tabitha Newman; Transfusion: Helinor Mcaleese, Pascal Winter, Gareth Heywood-Beldon, Julia Lancut, Catherine Booth, Manaf Al-Bayati and the Blood Transfusion laboratory team.

*Ninewells Hospital:* Pauline Austin, Susan Chapman, Louise Cabrelli; Transfusion: Katie Hands, E Knight, L Macdonald.

*Norfolk and Norwich University Hospital:* Simon Fletcher, Jurgens Nortje, Deirdre Fottrell-Gould, Georgina Randell, Katie Stammers, Gail Healey, Marta Goncalves Pinto; Transfusion: Deborah Asher, Janet Pring, Kathy Ford, Alison Rudd and the Blood Transfusion laboratory team.

*Northampton General Hospital:* Mohsin Zaman, Einas Elmahi, Andrea Jones, Kathryn Hall; Transfusion: Karen Spreckley, Mike Comery, Raquel Bisa, Tace Darling and the Blood Transfusion laboratory team.

*Northern General Hospital, Sheffield:* Gary H Mills, Ajay Raithatha, Kris Bauchmuller, Kim Ryalls, Kate Harrington, Helen Bowler, Jas Sall, Richard Bourne; Transfusion: Brian Taylor, Sabira Ali, Helen Wilkinson, Geneen Powell, Zoe Johnson and the Blood Transfusion laboratory teams.

*Northwick Park Hospital NHS Trust:* Jamie Gross, Natalie Massey, Olumide Adebambo, Matilda Long, Kiran Tony; Transfusion: Kathleen Buckmire, Rebecca Kahari, Rebecca Patel, Donna Wiles

*North Manchester General Hospital:* Zoe Borrill, Tracy Duncan, Andrew Ustianowski, Alison Uriel, Ayaa Eltayeb, Jordan Alfonso, Samuel Hey, Joanne Shaw, Claire Fox, Gabriella Lindergard, Bethan Charles, Bethany Blackledge, Karen Connolly, Jade Harris; Transfusion: Thomas Trimble, Emma Cooperwaite, Carmel Parker, Sarah Pendlebury, Jayne Peters, S Khan, E Anyanwu, M Evans and the Blood Transfusion laboratory teams.

*North Middlesex University Hospital:* Jeronimo Moreno Cuesta, Kugan Xavier, Dharam Purohit, Munzir Elhassan, Anne Haldeos, Rachel Vincent, Marwa Abdelrazik, Samuel Jenkins, Arunkumar Ganesan,

Rohit Kumar, David Carter, Dhanalakshmi Bakthavatsalam, Alasdair Frater, Malik Tahir Saleem; Transfusion: Shehan Palihavadana, Karen Madgwick and the Blood Transfusion laboratory team.

*Oxford University Hospitals:* Matthew Rowland, Paula Hutton, Archana Bashyal, Neil Davidson, Clare Hird; Transfusion: Julie Staves, Caryn Vander Riet, W Byrne and the Blood Transfusion laboratory team.

*Pilgrim Hospital Boston:* Manish Chhablani, Gunjan Phalod, Amy Kirkby, Simon Archer, Kimberley Netherton; Transfusion: Ben Holmes, Stuart MacDonald, Anthony Jackson, Carol Richardson and the Blood Transfusion laboratory team.

*Princess Royal Hospital:* Denise Skinner, Jane Gaylard, Julie Newman; Transfusion: Julie Cole, Laura Humber and the Blood Transfusion laboratory team.

*Princess of Wales Hospital:* Sonia Sathe, Lisa Roche, Ellie Davies; Transfusion: Toni Home, Kevin Jones, Rebecca Lawton, Tomos Edwards, Ceri Brookes, Kathryn Hennessy, Rebecca Ward and the Blood Transfusion laboratory team.

*Poole Hospital:* Henrik Reschreiter, Julie Camsooksai, Sarah Patch, Sarah Jenkins, Charlotte Humphrey; Transfusion: Maraneka Greenslade, Lorraine Poole, Nicola Dewland, Max Deighton and the Blood Transfusion laboratory team.

*Queen Alexandra Hospital Portsmouth:* David Pogson, Steve Rose, Zoe Daly, Lutece Brimfield, Angie Nown; Transfusion: Alison Davies, Kay Heron, Gwynn Matthias and the Blood Transfusion laboratory team.

*Queen Elizabeth Hospital, Birmingham:* Dhruv Parekh, Colin Bergin, Michelle Bates, Christopher McGhee, Daniella Lynch, Khushpreet Bhandal, Kyriaki Tsakiridou, Amy Bamford, Lauren Cooper, Tony Whitehouse, Tonny Veenith, Elliot Forster, Martin O'Connell; Transfusion: Suzy Morton, Kathryn Wood, Jessica Jones and the Blood Transfusion laboratory teams.

*Queen Elizabeth University Hospital, Glasgow:* Malcolm AB Sim, Sophie Kennedy Hay, Steven Henderson, Maria Nygren, Eliza Valentine; Transfusion: Claire McKie, Alison Hanlon, April Molloy, Margaret McGarvey and the Blood Transfusion laboratory team.

*Queen's Hospital, Burton:* Amro Katary, Gillian Bell, Louise Wilcox, Michail Mataliotakis, Paul Smith, Murtaza Asif Ali, Agah Isguzar; Transfusion: Julie Buchan, Jeby Jeyachandran, Roger Shiers, Satvinder Kaur, Humayun Ahmad, Heather Clarke, Kararina Kacinova and the Blood Transfusion laboratory teams.

*Queen's Hospital, Romford:* Mandeep-Kaur Phull, Abbas Zaidi, Tatiana Pogreban, Lace Paulyn Rosaroso; Transfusion: Xiaohui Tang, Anne Minogue, Amina Abdulle, Nafissah Munu, Nickolas Page, Sonya Balkee, Priti Patel, Colleen Sanderson, Rose Gad, Abigail Asquith, Arron Minhas, Garikayi Marange, Sandy Lockyer

*Queens Medical Centre and Nottingham City Hospital:* Daniel Harvey, Benjamin Lowe, Megan Meredith, Lucy Ryan, DREEAM Research Team; Transfusion: Michael John Skill, Nicola Horton-Turner, Abbi Tervit, Chloe Wilkes, Rachel Rose, Scott Springworth, Hayley Bond, Stacey Robinson, Louise Allen, Cherry Chang, Maria Isabel Saez-Garcia-Holloway and Blood Transfusion laboratory team.

*The Rotherham NHS Foundation Trust:* Anil Hormis, Rachel Walker, Dawn Collier, Sarah Kimpton, Susan Oakley; Transfusion: Robert Stirk, Rachel Perkins, Stuart Lord, Carron Bilton and the Blood Transfusion laboratory team.

*Royal Alexandra Hospital:* Kevin Rooney, Natalie Rodden, Nicola Thomson, Deborah McGlynn, Lynn Abel, Lisa Gemmell, Radha Sundaram, James Hornsby; Transfusion: Hospital Blood Transfusion Team

*Royal Berkshire Hospital:* Andrew Walden, Liza Keating, Matthew Frise, Sabi Gurung Rai, Shauna Bartley; Transfusion: Kelly Kemsley, Tanya Hawkins and the Blood Transfusion laboratory team.

*Royal Bournemouth and Christchurch Hospitals:* Martin Schuster-Bruce, Sally Pitts, Rebecca Miln, Laura Purandare, Luke Vamplew; Transfusion: Michael Trevett, Annette Jose, Lorraine Mounsey, Charlotte Baylem, Tamsin Haydon, V Chandler-Vizard, B Grice and the Blood Transfusion laboratory team.

*Royal Brompton Hospital:* Brijesh Patel, Debra Dempster, Mahitha Gummadi, Natalie Dormand, Shu Fang Wang

*Royal Cornwall NHS Trust:* Michael Spivey, Sarah Bean, Karen Burt, Lorraine Moore, Fiona Hammonds, Carol Richards

*Royal Devon and Exeter NHS Foundation Trust:* Christopher Day, Letizia Zitter, Sarah Benyon

*Royal Glamorgan Hospital:* Jayaprakash Singh, Ceri Lynch, Lisa Roche, Justyna Mikusek, Bethan Deacon, Keri Turner; Transfusion: Toni Home, Kevin Jones, Rebecca Lawton, Tomos Edwards, Ceri Brookes, Kathryn Hennessy, Rebecca Ward and the Blood Transfusion laboratory team.

*Royal Gwent Hospital*: Evelyn Baker, John Hickey, Shreekant Champanerkar, Lindianne Aitken, Lorraine Lewis-Prosser, Christie James; Transfusion: Cheryl Davies, Jennifer Summers and the Blood Transfusion laboratory team.

*Royal Hallamshire Hospital, Sheffield:* Gary H Mills, Norfaizan Ahmad, Matt Wiles Jayne Willson; Transfusion: Brian Taylor, Sabira Ali, Helen Wilkinson, Geneen Powell, Zoe Johnson and the Blood Transfusion laboratory teams.

*Royal Hampshire Hospitals:* Irina Grecu, Jane Martin, Caroline Wrey Brown, Ana-Marie Arias, Emily Bevan, Samantha Westlake; Transfusion: Marie Cundall, Olivia Martins, Catherine Wilson, Linda Holloway and the Blood Transfusion laboratory team.

*Royal Infirmary of Edinburgh:* Thomas H Craven, David Hope, Jo Singleton, Sarah Clark, Corrienne McCulloch, Simon Biddie; Transfusion: Jonathan Falconer, Jane Oldham and the Blood Transfusion laboratory team.

*Royal Liverpool University Hospital:* Ingeborg D Welters, David Oliver Hamilton, Karen Williams, Victoria Waugh, David Shaw, Suleman Mulla, Alicia Waite, Jaime Fernandez Roman, Maria Lopez Martinez, Brian Johnston; Transfusion: Karen Knowles, Holly Wissett, Rebecca Wright, Cristina Dragomir, Abby Peacock, Joshua Bell, Jan Gorry, Iain Houghton, Paula Burgess and all members of the lab BMS and MLA teams.

*Royal London Hospital:* Zudin Puthucheary, Timothy Martin, Filipa Santos, Ruzena Uddin, Maria Fernandez, Fatima Seidu, Alastair Somerville, Mari-Liis Pakats, Salma Begum, Tasnin Shahid; Transfusion: Helinor Mcaleese, Pascal Winter, Gareth Heywood-Beldon, Julia Lancut, Catherine Booth, Manaf Al-Bayati and the Blood Transfusion laboratory team.

*The Royal Free Hospital:* Sanjay Bhagani, Mark De Neef, Sara Mingo Garcia, Amitaa Maharajh, Aarti Nandani, Jade Dobson, Gloria Fernando, Christine Eastgate, Keith Gomez, Zakee Abdi; Transfusion: Rita Atugonza, Anna Li, Jenny Li, Seble Tekle, Ann-Marie Ellis, Anushka Natarajan, Stephanie Mellin and the Blood Transfusion laboratory team.

*The Royal Marsden NHS Foundation Trust:* Kate Colette Tatham, Shaman Jhanji, Ethel Black, Arnold Dela Rosa, Ryan Howle, Ravishankar Rao Baikady; Transfusion: E Malundas, A Mohamed, L Desai.

*The Royal Oldham Hospital:* Redmond P Tully, Andrew Drummond, Joy Dearden, Jennifer E Philbin, Sheila Munt; Transfusion: Allameddine Allameddine, Jane Uttley, Susan Andrews, Christopher Porada, Kaiser Mushtaq, Debra Curley, Helen Morris, Olajide Akinwumiju, Che Okyne-Turkson, Sara Flynn and the Blood Transfusion laboratory teams.

*The Royal Wolverhampton NHS Trust:* Shameer Gopal, Jagtar-Singh Pooni, Saibal Ganguly, Andrew Smallwood, Stella Metherell Transfusion: Maxine Boyd, Mary Blanton, Mike Herbert and the Blood Transfusion Laboratory team.

*Royal Papworth Hospital:* Alain Vuylsteke, Charles Chan, Saji Victor, COVID Research Team, Papworth Hospital; Transfusion: Martin Muir, Joseph Joseph, Cathy Flatters, Alex Hudson and the Biomedical Scientists and Associate Practitioners of the Transfusion laboratory.

*Royal Stoke Hospital:* Ramprasad Matsa, Minerva Gellamucho, Michelle Davies; Transfusion: Rebecca Sivers, Jane Graham, Pamela Irving, Dionne Bentley, Angela Salmon, Rosemary Rushworth, Charlotte Brackstone, Charles Baker, Susan Mitchell, Elizabeth Brown, Sarah O'Brien, Jenni Jeffrey, F Perkins, Diane Murdoch and the Blood Transfusion laboratory teams.

*Royal Surrey County Hospital:* Ben Creagh-Brown, Cheryl Marriot, Armorel Salberg, Louisa Zouita, Sarah Stone, Natalia Michalak, Sinead Donlon, Shelia Mtuwa, Irving Mayangao, Jerik Verula, Dorota Burda, Celia Harris, Emily Jones, Paul Bradley, Esther Tarr, Lesley Harden, Charlie Piercy; Transfusion: Sudharkar Vimalanathan, Jo Lawrence and the Blood Transfusion laboratory team.

*Royal United Hospital Bath:* Jerry Nolan, Ian Kerslake, Tim Cook, Tom Simpson, James Dalton, Carrie Demetriou, Sarah Mitchard, Lidia Ramos, Katie White, Toby Johnson, William Headdon, Stephen Spencer, Alison White, Lucy Howie; Transfusion: Adele Wardle, Amanda Dornan, Wayne Vietri, Kirsten Pass, Helen Maria-Osborn, Hoodo Ali, Clive Risbridger, Alexis Buchanan, Krystyna Turek, Daniel Loveys, Lorna Swan, Rebecca Coles, Parmi Perera, Zanele Sibanda and the laboratory team.

*Russells Hall Hospital:* Michael Reay, Steve Jenkins, Angela Watts, Eleanor Traverse, Stacey Jennings, Vikram Anumakonda, Caroline Tuckwell, Karen Pearson, Kath Harrow, Julie Matthews, Karen McGarry, Vanessa Moore, Lucie Smith, Anna Summerfield; Transfusion: Anna Smith, Caroline Tuckwell and the Blood Transfusion laboratory team.

*Salisbury NHS Foundation Trust:* Phil Donnison, Ruth Casey, Ben Irving, Wadzanai Matimba-Mupaya, Catherine Reed, Alpha Anthony, Fiona Trim, Lenka Cambalova, Debra Robertson, Anna Wilson; Transfusion: Caroline Matthews, Sheryl Haviland, Chloe Stacey and the Blood Transfusion laboratory team.

*Salford Royal NHS Foundation Trust:* Paul Dark, Alice Harvey, Reece Doonan, Liam McMorrow, Karen Knowles, Jessica Pendlebury, Stephanie Lee, Jane Perez, Bethan Charles, Tracy Marsden, Melanie Taylor, Angiy Michael, Matthew Collis, Andrew Claxton, Wadih Habeichi, Dan Horner, Melanie Slaughter, Vicky Thomas, Nicola Proudfoot, Claire Keatley; Transfusion: Deborah Seddon, James Wesson, Lydia Baxter, Laura Cooper, Eva Loutraris, all Haematology and Blood Transfusion staff.

*Sandwell and West Birmingham NHS Trust:* Jonathan Hulme, Santhana Kannan, Fiona Kinney, Ho Jan Senya, Anne Hayes; Transfusion: Loraine Blackwood, Vipul Chauhan

*Sherwood Forest Hospitals NHS Foundation Trust:* Valli Ratnam, Mandy Gill, Jill Kirk, Sarah Shelton; Transfusion: Joanne Wren, Jane Walden, Leanne Hostler, Senior BMS and the Blood Transfusion laboratory team.

*South Tyneside District Hospital:* Christian Frey, Riccardo Scano, Madeleine McKee, Peter Murphy; Transfusion: Lee Sudlow, Jill Caulfield, Jonathan Trattles, Pauline Patterson, Sue Matthews and all in the Blood Transfusion laboratory teams.

*Southmead Hospital:* Matt Thomas, Ruth Worner, Beverley Faulkner, Emma Gendall, Kati Hayes, Hayley Blakemore, Borislava Borislavova

*St. Bartholomew's Hospital:* Colin Hamilton-Davies, Carmen Chan, Celina Mfuko, Hakam Abbass, Vineela Mandadapu

*St. George's Hospital:* Susannah Leaver, Kamal Patel, Sarah Farnell-Ward, Romina Pepermans Saluzzio, Sam Rawlins, Christine Sicat; Transfusion: Chloe Orchard, Vince Michael, Kelly Feane, James Uprichard and all the transfusion lab team.

*St. Mary's Hospital:* Anthony Gordon; Dorota Banach Ziortza Fernández de Pinedo Artaraz, Leilani Cabreros, Victoria Latham; Transfusion: A Rahman, David Johnson, Linda Chapple, Eugenia Nweje, K Sabljak, S Kassa, U Munu, H Dawson, Fiona Regan, Fateha Chowdhury, Vashira Chiroma and Blood Transfusion laboratory teams.

*St. Peter's Hospital, Chertsey:* Ian White, Maria Croft, Nicky Holland, Rita Pereira; Transfusion: Joanne Finden, Z Takats, Nelsonseelan Johnson and the Blood Transfusion Laboratory team

*Stepping Hill Hospital, Stockport:* Ahmed Zaki, David Johnson, Hywel Garrard, Vera Juhaz, Louise Brown, Abigail Pemberton; Transfusion: Raisa Zaman, Brendan Devine and the Blood Transfusion laboratory team.

*Sunderland Royal Hospital:* Alistair Roy, Anthony Rostron, Lindsey Woods, Sarah Cornell; Transfusion: Lee Sudlow, Jill Caulfield, Jonathan Trattles, Pauline Patterson, Sue Matthews and all in the Blood Transfusion laboratory teams.

*Swansea Bay University Health Board:* Suresh Pillai, Rachel Harford, Helen Ivatt, Debra Evans, Suzanne Richards, Eilir Roberts, James Bowen, James Ainsworth; Transfusion: David Payne, Karen Phillips, Liz Park, Sarah John, Pamela Diamond and the Blood Transfusion laboratory team.

*Torbay and South Devon NHS Foundation Trust:* Angela Foulds, Adam Revill; Transfusion: Alistair Penny, Steve Mills, Julia Pinder, Helen Randall, Meadow Fisher Crisp and the Blood Transfusion laboratory team.

*United Lincolnshire NHS Trust:* Russell Barber, Anette Hilldrith, Gunjan Phalod; Transfusion: Ben Holmes, Stuart MacDonald, Anthony Jackson, Carol Richardson and the Blood Transfusion laboratory team.

*University Hospitals Bristol & Weston NHS Foundation Trust:* Jeremy Bewley, Katie Sweet, Lisa Grimmer, Rebekah Johnson, Rachel Wyatt, Karen Morgan, Siby Varghese, Charlotte Bradbury, Joanna Willis, Emma Stratton, Laura Kyle, Daniel Putensen, Kay Drury, Agnieszka Skorko; Transfusion: Soo Cooke, Ruth Cowburn, Sharif Goolam-Hossen, Laura Johnston, Ana Parejasanchez, Adele Wardle, Stephen White and the Blood Transfusion laboratory team.

*University Hospitals Coventry & Warwickshire NHS Trust:* Pamela Bremmer, Geraldine Ward, Christopher Bassford; Transfusion: Angela Busby, Julie Northcote, and the Blood Transfusion laboratory team.

*University Hospital of North Tees:* Farooq Brohi, Vijay Jagannathan, Michele Clark, Sarah Purvis, Bill Wetherill; Transfusion: Marie Teresa Walker, Debra Anne Cox, and the Blood Transfusion laboratory team.

*University Hospital Southampton NHS Foundation Trust:* Ahilanandan Dushianthan, Rebecca Cusack, Kim de Courcy-Golder, Karen Salmon, Rachel Burnish, Simon Smith, Susan Jackson, Winningtom Ruiz, Zoe Duke, Magaret Johns, Michelle Male, Kirsty Gladas, Satwinder Virdee, Jacqueline Swabe, Helen Tomlinson; Transfusion: Kerry Dowling, Jonathan Ricks, Samantha Carrington, Philippa Downey, Sarah Mumford, Tracey Lofting and the Blood Transfusion laboratory team.

*Warwick Hospital:* Ben Attwood, Penny Parsons, Bridget Campbell, Alex Smith;

*Watford General Hospital:* Valerie J Page, Xiao Bei Zhao, Deepali Oza, Gail Abrahamson, Ben Sheath, Chiara Ellis; Transfusion: D Beckford-Smith, S Bradley

*Western General Hospital, Edinburgh:* Jonathan Rhodes, Thomas Anderson, Sheila Morris; Transfusion: Jonathan Falconer, Jane Oldham and the Blood Transfusion laboratory team.

*Whipps Cross Hospital:* Charlotte Xia Le Tai, Amy Thomas, Alexandra Keen, Carey Tierney, Nimca Omer, Gina Bacon; Transfusion: Helinor Mcaleese, Pascal Winter, Gareth Heywood-Beldon, Julia Lancut, Catherine Booth, Manaf Al-Bayati and the Blood Transfusion laboratory team.

*Whiston Hospital*: Dr Ascanio, Tridente, Karen Shuker, Jeanette Anders, Sandra Greer, Paula Scott, Amy Millington, Philip Buchanan, Jodie Kirk;

*Wirral University Teaching Hospital NHSFT:* Craig Denmade, Girendra Sadera, Reni Jacob, Cathy Jones, Debbie Hughes; Transfusion: Steven Carter, Lisa Delaney, Debra Woods, and the Blood Transfusion laboratory team.

*Worcester Royal Hospital:* Stephen Digby, Nicholas Cowley, Laura Wild, Jessica Thrush, Hannah Wood, Karen Austin; Transfusion: Camran Khan, Gillian Godding, Emily Murphy, Emma Loxley, and the Blood Transfusion laboratory team.

*Wrexham Maelor Betsi Cadwaladr University Hospital*: David Southern, Harsha Reddy, Sarah Hulse, Andrew Campbell, Mark Garton, Claire Watkins, Sara Smuts; Transfusion: Tony Coates, Samantha Thomas-Wright, Christopher Bennett and the Blood Transfusion laboratory team

*Wrightington, Wigan and Leigh Teaching Hospitals NHS Foundation Trust:* Alison Quinn, Benjamin Simpson, Catherine McMillan, Cheryl Finch, Claire Hill, Josh Cooper; Transfusion: Louise McCreery, Joanne Cooper, Jim Wesson and the Blood Transfusion laboratory team.

*Wye Valley NHS Trust:* Joanna Budd, Charlotte Small, Ryan O'Leary, Janine Birch, Emma Collins, Andrew Holland; Transfusion: Ian Hancock, Helen Slade, Margaret Kevern and the Blood Transfusion laboratory team.

*Wythenshawe Hospital*: Peter D G Alexander, Tim Felton, Susan Ferguson, Katharine Sellers, Luke Ward; Transfusion: Thomas Trimble, Emma Cooperwaite, Carmel Parker, Sarah Pendlebury, Jayne Peters, S Khan, E Anyanwu, M Evans and the Blood Transfusion laboratory teams.

*York Teaching Hospital*: David Yates, Isobel Birkinshaw, Kay Kell, Zoe Scott, Harriet Pearson; Transfusion: Claire Purcell, Allison Thorpe, Catrina Ivel, Jenny Fullthorpe and the Blood Transfusion laboratory team.

***United States of America***

*University of Pittsburgh Research Staff:* CRISMA Center—Kelsey Linstrum, Stephanie Montgomery, Kim Basile, Dara Stavor, Dylan Burbee, Amanda McNamara, Renee Wunderley, Nicole Bensen, Aaron Richardson; MACRO Center—Peter Adams, Tina Vita, Megan Buhay, Denise Scholl, Matthew Gilliam, James Winters, Kaleigh Doherty, Emily Berryman

*UPMC Hospital Champions:* UPMC Altoona—Mehrdad Ghaffari, UPMC East—Meghan Fitzpatrick; UPMC Jameson and Horizon —Kavitha Bagavathy; UPMC Mercy—Mahwish Hussain, Chenell Donadee; UPMC Williamsport—Emily Brant; UPMC McKeesport—Kayla Bryan-Morris, John Arnold and Bob Reynolds; UPMC Hamot—Gregory Beard; UPMC Presbyterian—Bryan McVerry, David Huang, Ghady Haidar, Alexandra Weissman, Florian Mayr; UPMC Passavant – Matthew Gingo;

*UPMC COVID Therapeutics Committee:* Erin McCreary, Elise Martin, Ryan Bariola, Alex Viehman, Jessica Daley, Alyssa Lopus, Mark Schmidhofer, UPMC Directors of Pharmacy

*UPMC ICU Service Center:* Rachel Sackrowitz, Chenell Donadee, Aimee Skrtich

*UPMC Wolff Center:* Tami Minnier, Mary Kay Wisniewski, Katelyn Mayak

*UPMC eRecord Team:* Richard Ambrosino, Sherbrina Keen, Sue Della Toffalo, Martha Stambaugh, Ken Trimmer, Reno Perri, Sherry Casali, Rebecca Medva, Brent Massar, Ashley Beyerl, Jason Burkey, Sheryl Keeler, Maryalyce Lowery, Lynne Oncea, Jason Daugherty, Chanthou Sevilla, Amy Woelke, Julie Dice, Lisa Weber, Jason Roth, Cindy Ferringer, Deborah Beer, Jessica Fesz, Lillian Carpio

*Data Collection/Curation Team:* Salim Malakouti (Computer Science, University of Pittsburgh), Edvin Music and Dan Ricketts (CRISMA Center), Andrew King (Biomedical Informatics, University of Pittsburgh), Gilles Clermont (Critical Care Medicine), Robert Bart (UPMC Health Services Division*)*

*UPMC Clinical Analytics:* Oscar Marroquin, Kevin Quinn, William Garrard, Kyle Kalchthaler

*UPMC Office of Healthcare Innovation:* Derek Angus

*Department of Emergency Medicine:* Alexandra Weissman, Donald Yealy, David Barton, Nadine Talia

*Department of Critical Care Medicine:* David Huang, Florian Mayr, Andrew Schoenling, Mark Andreae, Varun Shetty, Emily Brant, Brian Malley, Chenell Donadee, Derek Angus, Christopher Horvat, Christopher Seymour, Timothy Girard, Gilles Clermont, Rachel Sackrowitz, Robert Bart

*Division of Infectious Diseases:* Ghady Haidar

*Division of Pulmonary, Allergy, and Critical Care Medicine:* William Bain, Ian Barbash, Mark Brown, Antu Das, Meghan Fitzpatrick, Christopher Franz, Stefanie Hannan, Georgios Kitsios, Ritchie Koshy, Sophia Lieber, Emily Lyons, John McDyer, Bryan McVerry, Kaveh Moghbeli, Iulia Popescu, Brian Rosborough, Faraaz Shah, Tomeka Suber

*Berry Consultants:* Roger Lewis, Michelle Detry, Anna McGlothlin, Christina Saunders, Mark Fitzgerald, Ashish Sanil, Scott Berry

*Global Coalition for Adaptive Research (GCAR):* Meredith Buxton, Brian Alexander, Tracey Roberts

*UPMC Laboratory Services*: Alan H. Wells

**Clinical Trials Groups**

The REMAP-CAP platform is supported by the Australian and New Zealand Intensive Care Society Clinical Trials Group, the Canadian Critical Care Trials Group, the Irish Critical Care Clinical Trials Network, the UK Critical Care Research Group and the International Forum of Acute Care Trialists.

REMAP-CAP was supported in the UK by the NIHR Clinical Research Network and we acknowledge the contribution of Kate Gilmour, Karen Pearson, Chris Siewerski, Sally-Anne Hurford, Emma Marsh, Debbie Campbell, Penny Williams, Kim Shirley, Meg Logan, Jane Hanson, Anne Oliver, Mihaela Sutu, Sheenagh Murphy, Latha Aravindan, Joanne Collins, Holly Monaghan, Adam Unsworth, Seonaid Beddows, Laura Ann Dawson, Sarah Dyas, Adeeba Asghar, Kate Donaldson, Tabitha Skinner, Nhlanhla Mguni, Natasha Muzengi, Ji Luo, Joanna O'Reilly, Chris Levett, Alison Potter, David Porter, Teresa Lockett, Jazz Bartholomew, Clare Rook, Hannah Williams, Alistair S Hall, Hilary Campbell, Holly Speight, Sandra Halden, Susan Harrison, Mobeena Naz, Charles Rounds, Kaatje Lomme, Johnathan Sheffield, William Van't Hoff, James D Williamson, Catherine Birch, Morwenna Brend, Emma Chambers, , Sarah Crawshaw, Chelsea Drake, Heather Harper, Stephen Lock, Eleanor OKell, Amber Hayes, Susan Walker, Jayne Goodwin, Helen Hodgson, Yvette Ellis, Dawn Williamson, Madeleine Bayne, Shane Jackson, Rahim Byrne, Sonia McKenna, Alison Clinton, NIHR Urgent Public Health Group:
https://www.nihr.ac.uk/documents/urgent-public-health-group-members/24638#Members

REMAP-CAP Immunoglobulin Domain was supported in Canada by the Canadian Institutes of Health Research and we acknowledge the contribution of Alexis Turgeon, Ryan Zarychanski, David Bellemare, Olivier Costerousse, Brian Cuthbertson, Sylvie Debigaré, Dana V Devine, Marc Germain, Donald M Arnold, Philippe Bégin, Jeannie L Callum, Marc Carrier, Michael Chassé, Deborah J Cook, Rana Daher, Nick Daneman, Shane W English, Dean A Fergusson, Robert A Fowler, Ewan C Goligher, Brett L Houston, Anand Kumar, Francois Lamontagne, François Lauzier, Patrick R Lawler, Sylvain Lother, John C Marshall, Lynne Moore, Srinivas Murthy, Bojan Paunovic, Nancy Robitaille, Alan T Tinmouth

REMAP-CAP was supported in France by the CRICS-TRIGGERSEP network

REMAP-CAP was supported in Ireland by the Irish Critical Care Clinical Trials Network and we acknowledge the contribution of Kate Ainscough, Kathy Brickell and Peter Doran.

REMAP-CAP was supported in the Netherlands by the Research Collaboration Critical Care the Netherlands (RCC-Net).

REMAP-CAP was supported in the United States by the Translational Breast Cancer Research Consortium, the UPMC Learning While Doing Program, and the Global Coalition for Adaptive Research

#### Blood Transfusion Services

**Australia**

*Australian Red Cross Lifeblood:* Wayne Bolton, Sue Ismay, June Lee, Dimitra Marinakis, Joanne Pink, Andrew Scott, Andrew Tracy

**Canada**

*Canadian Blood Services:* Shrefee Abouzeenni, Sheila Allan, Kristen Andersen, Corinne Anderson, Joel Armer-Ducker, Michelle Armstrong, Tanya Atallah, Michelle Aube, Harleen Badhesha , Dawn Baglole, Clariza Bagnas, Penny Bakhtiari, Nancy Banning, Mercedes Barillas, Nathaly Barnett , Jan Barwell, Lynette Beaudin, Nathalie Belec , Glenda Bell, Ann Beninato, Carrie Bennett Mills, Andrea Benoit, Lauren Berg, Claudia Berscheid, Jennifer Biemans, Kelly Bignell, Annette Blackwell, Margie Blom, Naomi Borkent, Brandon Bouchard, Gordana Bozovic, Val Brennan, Randy Broom, Emilie Brunet , Pierrette Bugden, Lori Bullock, Vitor Buonavoglia, Annmarie Buoy, Karla Campbell, Sarah Carr, Petrus Chan, Sunita Chand, Terry Channon, Sarah Charnock, Emily Cheung, Christian Choquet, Judy Yung Ching Chou, Jae Chung, Rob Cimaglia, Gwen Clarke, Julie Cleverdon, Pamela Conrod, Mary Corriveau, Mylene Crosby, Oriela Cuevas, Raj Damhar, Carol Daniels, Karen Darling, Janice Dawe, Lisa Dawe, Rosanne Dawson, Dylene Deans, Dianne de Jong, Dana Devine, Trudy Devine, Bodine Dewitt, Heather Donkin, Jan Douziech, Steven Drews, Hannah D'Silva, Deanne Dunphy, Brenda Du Plessis, Simone Ebbinghaus, Cathy Ellsworth, Ian Entino, Pam Fairclough, Kelly Falle, Bill Ferguson, Xiomara Ferndadez, Elaine Fitzimmons, Sara Ford, Kirsten Foster, Terrie Foster, Glenda Fox, Liesa Friedrich, Beth Frise, Ashley Gallant, Koreena Gallant, Cindy Gardner, Jocelynn Gardner, Kim Gaudet, Julie Gazmen, Joel Gilchrist, Balkar Gill, Leah Glasgow, Rayna Glazenburg, Tiphanie Gonzales, Anaita Goodie, Jessica Goulet, Tanya Gray, Maria Cecilia Graza, Amanda Griffith, Elodie Guitteaud, Sara Habel Liboiron, Sandra Halliday, Tracy Hanna, Trish Hansen, Naja Harper, Anita Hebert, Heather Hennebury, Rachel Hera, Tanya Herrell, Michael Hill, Joan Hoar, Valerie Hodder, Karen Hoiland, Michelle Hollands, Lynn Holt, Patti Hooley, Kim Horne, Krissy Hoskin, Pat Hoszko, Kelsey Howard,

David Howe, Colleen Hrabi, Kjierstin Hubka, Chelsey Hugelshofer, Roberta Hurman, Alison Hutchinson, Tammy Ison, Craig Jenkins, Marcella Jimenez, Deanna Jones, Glenda Jorritsma, Jaswinder Kaur, Ellen Kelly, Gord Kerr, Meenu (Promila) Khera, Andrea King, Michelle Knight, Rebecca Kovacevic, Rhonda Krahn, Wanda Krywiak, Kelly Kuntz, Jennifer Lafrenier, Manon Lahaie, Barb Laing, Brenda Lamb, Chantal Larivee, Cheryl Larner, Marilyne Larose, Marylee Lee, Wendy Lee, Wanda Lefresne, Danielle Leguard-White, Brenda Lukasik, Kathleen Lundy, Sherilynn Kavanagh, Kevin MacDonald, Peter MacDonald, Sharon MacLeod, Colleen MacKenzie, Debbie MacKenzie, Nancy MacNeil , Carole Main, Elmer Manuel, Guy Marcotte ,Maria Martins ,John Mazerall ,Shannon Cormick , Charlene McCracken, June McDonald, Mel McGee, Sheila McKilop , Janet McManus ,Joanne McMurdo ,Sue Michalowicz, Margaret Miedema, Adriana Minella, Liliana Miranda, MaryAnne Moreau, Steve Morrisey, Ivan Mounitsyn, Debbie Moyst, Mike Mullowney, Andrea Murphy, Judith Nabess, Gina Neves, Genelle Nicholson, Sukhi Nijjar, Josee Noel, Joelle Nordstrom, Tansey O'Connor, Violet O'Donnell, Kaitlyn O'Halloran, Lisa Orr, Darlene Osborne, Roxanne Osmond, Tanya Paddock, Jennifer Pade, Kimberley Palmer, Chantale Pambrun, Rob Pankhurst, Kerry Parsons, Mithal Patel, Trena Patzer, Debra Pavelich, Shelley Peterson, Tanya Petraszko, Tasha Pettigrew, Sharon Peyton, Janet Piersma, Mary Pitcher, Barb Pollock, Michelle Polson, Carolyn Powel, Ashton Prefontaine, Allison Prihoda, Lisa Privatera, Linda Prokopiuk , Mailene Quinagon, Sandra Randall, Ramina Randhawa, Connie Rasko, Lorna Retzlaff, Kamila Reznicek, Michelle Ritchot, Marlene Roach, Cherie Robson, Cassidy Rodley, Andrea Rogers, Mercedes Rosario, Joanne Ross, Natalie Ross, Patrizia Ruoso, Lisa Rusk, Michael Sanchez, Robert Sawler, Roxanne Schroeder, Marilyn Schuett, Judy Seaman, Faramarz Sedigh-Zadeh, Marlene Shade, Kimberly Shaughnessy , Dylan Shield, Christine Shin, Monyra Siek, Adrienne Silver, Brenda Simpson, Janice Sloan, Patricia Smith, Tracy Smith, Janet Smyrski, Tom Song, Monique Sosa, Jennifer Speidel, Bridgette Spires, Tony Steed Carole St Onge-Legault, Nina Sull, Helen Tackaberry, Donna Tasker, Asley Taylor, Saba Teklehaimanot, Patricia Tendler, Susan Theander, Deepak Thomas , Sylvia Torrance, Anne Trueman, Jonni-Lyn Van Deursen, Austin Veen, Tracy Velcich, Steve Villeneuve, Leanne Voutour, Eleanor Ward, Lea Warren, Kathryn Webert, Melanie Whynot, Katelyn Witherell, Paula Wojcik, Carol Wong, Sheila Wowchuk, Sarah Woycik, Alison Wright, Julie Wu, Patricia Yedynak, Michelle Zeller, Anna Zimperi

*Héma-Québec*: Isabelle Allard, Micheline Antar, Renée Bazin, Julie Beaudoin, Maude Bilodeau, Virginie Boisclair, Daniel Boutin, Lucie Boyer, Stéphanie Brisson, Marie-Eve Brulotte, Éric Chamberland, Marc Cloutier, Mathieu Drouin, Éric Ducas, Nathalie Dussault, Marie-Josée Fournier, Hélène Gagné, Louis-Philippe Gagné, Amaury Gaussen, Marc Germain, Sébastien Girard, Donald Gironne, Annie Jacques, Hélène Lamoureux, Patricia Landy, Maryse Leclerc, Luc Lesage, Antoine Lewin, Chan-Hoa Ly, Nathalie

Marin, Caroline Masse, Laurent-Paul Ménard, Christine Milot, Marie-Ève Nolin, Kheng Ly Oueng, Mélisande Paquet, Émilie Roberge, Caroline Parent, Éric Parent, Vicky Parent, Josée Perreault, Claudia Mireille Pigeon, Isabelle Rabusseau, Pascale Riverin, Pascal Rouleau, Synthia Sauvageau, Carl Simard, Leila Snouber, Stéphane Thellend, Sylvie Tremblay, Tony Tremblay, Catherine Viau; *Plasma distribution team and hubs:* Chantal Armali, Valérie Arsenault, Marie-Christine Auclair, Jeannie Callum, Connie Colavecchia, Line Daigle, Melissa Dougherty, Joanne Duncan, Alioska Escorcia, Stephen Fitzgibbon, Lorrie Giorgi, Janice Hawes, Erin Jamula, Zofia Kelly, Jeff Kinney, Diana Kobes, Robin Lawrence, Lai Ha Lee, Paula Lehto, Kim Licanio, Kayla Lucier, Heather Luyckx, Bruce Lyon, Patty MacNally, Sarah Magwood, Joanna McCarthy, Suzanna Medic, Dimpy Modi, Liz Molson, Darlene Mueller, Nikki Pilutti, Tracey Pronyk-Ward, Margaret Roche, Fernanco Selene, Graham Smith, Melanie St-John, Sharon Sutherland, Alan Tinmouth, Melanie Tokessy, Maxime Veillette, Sherry Walters, Michelle Zeller

**<u>New Zealand</u>**

*New Zealand Blood Service (NZBS):* Richard Charlewood; and all staff at NZBS who have worked on the convalescent plasma project.

**<u>United Kingdom</u>**

*Convalescent Plasma Clinical Trials Unit (on behalf of the UK Blood Services):* Emily Arbon, Francesca Clemons, Nikki Dallas, Alison Deary, Amy Evans, Chloe Fitzpatrick-Creamer, Lise Estcourt (lead), Claire Foley, Renate Hodge, Cara Hudson, Katie Keen, Emma Laing, Maria Lobo-Clarke, Joanne Mullings, Roshni Paul, Gillian Powter, Shaminie Shanmugaranjan, Samaher Sweity, and all staff at NHSBT Clinical Trials Unit that have worked on the Convalescent Plasma Project

*NHS Blood and Transplant (NHSBT):* David Abdo; Lisa Adams; Dominic Affron; Sophia Ahmed; Giulia Airoldi; Naim Akhtar; Beverley Allen; Laura Allen; Jess Allen; Rekha Anand; Kevin Anderson; George Apostolides; Andrea Arroyo; Glen Ashby; Ben Ashford; Suhail Asghar; Richard Athay-Hunt; Stephen Bailey; Gagan Bali; Lauren Banks; Bharat Bansal; Alex Barber; Pavinder Bassi; Betsy Bassis; Ian Bateman; Donna Batty; Pete Baughan; Peter Baughan; Lee Bayliss; Eileen Bays; Kirk Beard; Alexander Bennett; Jane Benstead; Jenny Blaxland; Donna Blofield; Sam Bolton; Cameron Borhani; Darren Bowen; Geoff Bowyer; David Brack; Rob Bradburn; Sarah Bradshaw; Suhail Brailsford; Angela Brazier; Terri Brittin; Andrew Broderick; David Broggio; Chloe Brown; Wayne Brown; Lesley Butler; Sophia

Butt; Ana Canabarro; Nuria Cardenas; Andrew Carlton; Rosanna Chavez; Suzanne Cherif; Karen Chinn; Wendy Clark; Andrew Clarkson; Lee Clifford; Lynne Clifford; Alison Cooper; Alice Cotton; Ryan Creighton; Ian Cron; Helen Crumley; Donna Cullen; Becca Curtis; Abi Curtis; Phil Daggett; Jane Davies; Sophia Davison; Alison Deary; Raji Dehulia; Randle Derbyshire; Jazz Dhaliwal; Jas Dhalliwal; Aman Dhesi; Maria Dineen; Stephen Dixon-Mould; Nikki Drogman; Sam Duff-Miller; Helen Duggan; Mick Duley; Steve Durgacharan; Caroline Eaton; Caroline Eaton-Howell; Dave Edmondson; Toby Elliott; Helen Escreet; Lise Estcourt; Amy Evans; Tony Eve; James Feely-Henderson; Kelly Flynn; Claire Foley; Daniel Fox; Gayle Franklin; Darron Franks; Ian Freestone; Jake Fryer; Robert Garney; Michael Garstka; Andrew Gibb; Sean Gilchrist; Gerry Gogarty; Harriet Gray; Patricia Grealish; James, Gregson; Alexandra Griffiths; Elizabeth Hampson; Helen Hanwell; Fiona Harper; Melanie Harper; Naomi Harris; Heli Harvala; Lee Hennen; Sharnie Beth Hesson; Martin Hill; Elizabeth Hillerby; Elisabeth Hilton; Marcus Hinde; Gillian Hollis; Laura Hontoria del Hoyo; Al Hunter; Isobel Hunter; Claire Ingall; Ali Issa; Douglas Jackson; Sam Jaggard; Rachel Johnson; Hazel Jones; Revati Joshi; Vasita Kara; Samantha Kearley; Jeremy Kellington; Maria Kerr; Tony King; Daniel Kirkbride; Ian Kirker; Emm, Laing; Rebecca Lanaway; Darren Latimer; Mark Lawley; Jemima Lawson; Christina Leaver; Irina Lenchuk; Laura Lordache; Kieron Loregnard; Hayley Lund; Sheila MacLennan; James Marshall; Oliver May; Lee Maynard; Joanne McMahon; Darren McNamara; Jenny Mehew; Jana Meier; Phil Mellor; Greg Methven; Claire Michelson; Gail Miflin; Ian Millar; Melanie Milloy; Reena Mistry; Rachel Moir; Ian Moody; Gillian Moore; Adam Morgan; Jackie Morgan; Rosna Mortuza; Jo Murphy; Fidelma Murphy; Parminder Naga; Shruthi Narayan; Salima Nazarali; Richard Newman; Jamie Norgrave; Tony Noyce; Andrew O'Brien; Dapo Odumeru; Dave Ormrod; Daniel Paul; Belinda Pelle; Craig Peters; Neil Philips; Danielle Phillips; Pat Phillips; Ella Poppitt; Gillian Powter; Maanvi Prasad; Amy Quirk; Wayne Rabin; David Ralphs; Alia Rashid; Michelle Ray; Grace Rees; Debbie Richards; Eric Richardson; Matthew Robb; David J Roberts; Leanne Roberts; Doug Robertson; Katie Robinson; David Rose; Nick Rylance; Umar Sabat; Nichola Salmons; Francisco San Diego; Jo Sell; Peter Senior; Shaminie Shanmugaranjan; Richard Shortland; Andrew Simpson; Alison Sivyer; Katie Smith; Deepti Sood; John Spence; Helen Spicer; Tony Staincliffe; Guy Stevens; Rebecca Stewart; Luke Streeter; Samaher Sweity; Silvia Taibo; Ran Tao; Holly Templar; Kate Tettmar; Wendy Thorne; Jason Towler; Claire Tranter; Sara Tryon; Anna Turco; Louise Turner; Ruth Turner; Laura Unitt; Charlotte Washington; Nicholas Watkins; Adam West; Victoria Whiteside; Kaitlyn Whitley; Ritchie Wildman; Holly Wilkinson; Jayne, Williams; Bruce Willian; Evie Wiltsher; Yun Man Wong; Carrie Anne Wood; Peter Wright; Lee Wright; Andrew

Yeatman; Lynn Zarb; Marian Zelman; Will Zuurbier; and all staff at NHSBT who have worked on the convalescent plasma project.

*Northern Ireland Blood Transfusion Service (NIBTS)*: Barry Bradley; Mark Clarke; Catherine Corry; Alison Geddis; Matt Gillespie; Pauline Gowdy; Ingrid Gray; Leah Hadzik; Karin Jackson, Emma Jefferson, Angela Macauley; Patricia Mackey; Kathryn Maguire; Martina McCalmont; Barbara Mullin; Janice Paul; Sharon Rainey; John Simpson; Fiona Waller; and all staff at NIBTS who have worked on the convalescent plasma project.

*Scottish National Blood Transfusion Service (SNBTS):* Sylvia Armstrong-Fisher, Moira Carter, Ryan Evans, Emily Hargreaves, Lisa Jarvis, Tania MacDonald, Cara MacGuigan, Lynn Mason, Lorna McLintock, Deborah McNaughton, Nicole Priddee, Megan Rowley, Elaine Stevenson, Amanda Stewart, Marc Turner, David Turner, and all staff at SNBTS who have worked on the convalescent plasma project.

*Serious Hazards of Transfusion (SHOT)*: Simon Carter-Graham, Shruthi Narayan (lead), David Poles, Victoria Tuckley; on behalf of the SHOT Office team and all members of the SHOT Steering Group and Working Expert Group

*UK Convalescent Plasma Steering Group*: Millie Banerjee, Betsy Basis, Mark Davies, Kay Ellis, Laura Hontoria del Hoyo, Ann Jarvis, Gail Miflin, Marina Pappa, Ian Rees, David Rose, Jonathan Sheffield, Charlotte Taylor, Chris Townsend

*Welsh Blood Transfusion Service (WBS):* Jacinta Abraham, Janet Birchall, Stuart Blackmore, Mark Briggs, Maria Cheadle, Michelle Evans, Chloe George, Stephen Jolles, Kelly Kent, Huw Lovett, Peter Richardson, Georgia Stevens, Lee Wong; and all staff at WBS who have worked on the convalescent plasma project.

**United States Blood Transfusion Services**

*Vitalant (blood service):* Joseph E. Kiss, Darrell J. Triulzi

#### Sample reception, processing, storage and testing laboratories

**Australia**

*Viral Infectious Diseases, The Doherty Institute for Infection and Immunity, The University of Melbourne:* Julian Druce, Samantha Grimley, Youry Kim, Sara Marrero Hernandez, Julie McAuley

*Australasian COVID-19 Trial (ASCOT) Biobank, The Doherty Institute for Infection and Immunity, The University of Melbourne:* Vi Nguyen and ASCOT Biobank Committee

*Kirby Institute, University of New South Wales:* Anu Aggarwal, Christina Fitcher, Sam McAllery, Alberto Ospina Stella, Stuart Turville

*Prince of Wales, University of New South Wales:* Sonia Issacs, Greg Walker

**Canada**

*CRCHUM, Montréal, Québec:* Sai Priya Anand, Guillaume Beaudoin-Bussières, Mehdi Benlarbi, Catherine Bourassa, Marianne Boutin, Jade Descôteaux-Dinelle, Andrés Finzi, Gabrielle Gendron-Lepage, Guillaume Goyette, Annemarie Laumaea, Halima Medjahed, Jérémie Prévost, Jonathan Richard

*Medical Microbiology & Immunology, University of Alberta, Edmonton, Alberta*: David Evans, James Lin.

*Zoonotic Diseases and Special Pathogens, National Microbiology Laboratory, Public Health Agency of Canada, Winnipeg, Manitoba:* Michael Chan, Kristina Dimitrova, Michael Drebot, Kathy Manguiat, Emelissa Mendoza, Clark Phillipson, Alyssia Robinson, Heidi Wood,

**United Kingdom**

*Experimental Medicine Division, Nuffield Department of Medicine, University of Oxford, Oxford:* Rachael Brown, S Cameron, Derrick Crook (lead), Daniel Ebner, Stephanie B. Hatch, Sarah J. Hoosdally, Alison Howarth, Leonidas Koukouflis, W Lason, Brian D. Marsden, Lucas Martins Ferreira, Hayleah Pickford, P Tamblin-Hopper, M Wolna

*Peter Medawar Building for Pathogen Research, Nuffield Department of Medicine, University of Oxford, Oxford:* Dung Nguyen, Jeremy Ratcliff, Peter Simmonds (lead), Sarah Williams

*Nuffield Department of Surgical Sciences, University of Oxford, Oxford:* Sarah Cross, Hannah McGivern, Marta Oliveira, Rutger Ploeg (lead), Sheba Zyengi

*Radcliffe Department of Medicine, University of Oxford, Oxford:* Miriam Barbosa, Mamu Boshir, Nick Ciccone, Abigail Lamikanra, Ullrich Leuschner, Sindhuja Pratheepkumar, David Roberts (Lead), Atal Roman, Wendy Slack, Alain Townsend, Hoi Pat Tsang, Huiyuan Xiao

*School of Immunology and Microbial Sciences, King's College London and biomedical Research Centre at King's health Partners, London*: Matthew Stewart Fish; Aislinn Rose Jennings; Jakob Jeriha; Carolyn Karman Lam; Jennifer Rynne; Manu Shankar-Hari (PI); Emma M Timms; Isabella Tosi; Silvia Cellone Trevelin; James Dylan Williams

**United States**

Alan Wells, Mark Brown, Antu Das, Stefanie Hannan, Sophia Lieber, Emily Lyons, Ritchie Koshy, John McDyer, Iulia Popescu

#### 2. Supplementary methods

##### Patient eligibility criteria

**Platform eligibility (Pandemic appendix to the core protocol)**

In order to be eligible to participate in the pandemic aspects of REMAP-CAP, a patient must have met the following criteria:

1. Adult patient admitted to hospital with acute illness due to suspected or proven pandemic infection

A potentially eligible patient who met any of the following criteria were excluded from participation in REMAP-CAP trial:

1. Death is deemed to be imminent and inevitable during the next 24 hours AND one or more of the patient, substitute decision maker or attending physician are not committed to full active treatment
2. Patient is expected to be discharged from hospital today or tomorrow
3. More than 14 days have elapsed while admitted to hospital with symptoms of an acute illness due to suspected or proven pandemic infection
4. Previous participation in this REMAP within the last 90 days

**Immunoglobulin Domain eligibility**

Patients who met the platform-level inclusion and none of the platform-level exclusion criteria were eligible for Immunoglobulin Domain if:

1. COVID-19 infection was confirmed by microbiological testing

Patients were excluded from the Immunoglobulin Domain if they had any of the following:

1. More than 48 hours had elapsed since intensive care unit (ICU) admission
2. Patient had already received treatment with any non-trial prescribed antibody therapy (monoclonal antibody, hyperimmune immunoglobulin, or convalescent plasma) intended to be active against COVID-19 during the hospital admission
3. More than 14 days had elapsed since hospital admission
4. The treating clinician believed that participation in the domain would not be in the best interests of the patient
5. Known hypersensitivity/allergy to plasma and plasma products
6. Known previous history of transfusion-related acute lung injury
7. Known objection to receiving plasma products

#### Definition of Moderate and Severe States

Patients were categorised into one of two states, which were mutually exclusive:

Moderate state was defined by:

- Not being admitted to an ICU, or
- Admitted to an ICU but not receiving organ failure support

Severe state was defined by receiving organ failure support in an ICU. An ICU could include an area of the hospital re-purposed to function as an ICU for surge capacity management. Organ failure supports that qualified a patient as severe were respiratory or cardiovascular organ support:

*Respiratory support*

- Provision of invasive mechanical ventilation
- Provision of non-invasive mechanical ventilation (including high flow nasal cannula with a flow rate of at least 30 litres per minutes and a fractional inspired oxygen concentration of 40% or higher)

*Cardiovascular support*

- Receiving infusion of vasopressor or inotropes or both

Patients were assigned to either moderate or severe state at the time of assessment of eligibility. Further details are provided in the Pandemic Appendix to the Core Protocol (REMAP-COVID), provided in the **Protocol and SAP**.

#### Definition of Immunodeficiency

This was the combination of immunosuppressive therapy and disease

*Immunosuppressive Therapy*: The patient has received therapy that has suppressed resistance to infection prior to this hospital admission, includes:

- Immunosuppression, chemotherapy within 4 weeks of admission
- Radiation
- High-dose steroid treatment (e.g. >1.5mg/kg methyl prednisolone or equivalent for ≥5 days)
- Long-term treatment steroid treatment (e.g. >20 mg/day of a steroid)

*Immunosuppressive Disease*: The patient has one or more disease(s) that is sufficiently advanced to suppress resistance to infection (excludes malignancy which has been in remission for 5 years or more):

- Acquired Immunodeficiency Syndrome (AIDS)

- Acute leukaemia
- Lymphoma
- Metastatic Cancer
- Myeloma
- Any other disease that is sufficiently advanced to suppress resistance to infection, for example:
- Primary or inherited immune deficiency syndromes
- Chronic leukaemia
- Aplastic anaemia or other causes of chronic neutropenia

#### Definition of per protocol

In this domain convalescent plasma given per protocol is defined as:

*Convalescent plasma assignment*: patients assigned to receive plasma will receive at least one and not more than two adult units of ABO compatible convalescent plasma (total volume 550ml ± 150ml) within 48 hours of randomization.

*Control assignment*: patients assigned to receive no plasma will not receive convalescent plasma at any time after randomization' for the control intervention.

#### Collection and distribution of convalescent plasma

**Australia**

Supply of CP to local sites was coordinated and managed by the Australian Red Cross Lifeblood.

*Donor selection*
Donors with prior COVID-19 infection, who meet eligibility criteria for acceptance of blood donors, and were at least 28 days from COVID-19 symptom resolution. CP collected from male donors only was used for clinical CP. The median time from test positivity to donation was 108 days (range 54-137 days).

*Plasma manufacture*
Convalescent plasma was collected by apheresis with volume of 250-310 mL, stored at or below 25 degrees Celsius, and met all regulatory requirements for use as clinical plasma.

*CP testing*
All CP underwent routine blood group and infectious disease testing as for any fresh blood component by Lifeblood. Each donation was tested by a reference laboratory using 3 separate tests, the Abbott Architect SARS-CoV-2 IgG CMIA, the Euroimmun anti-SARS-CoV-2 enzyme-linked immunosorbent assay (ELISA) and a virus microneutralization assay using Vero E6 cells. The results for registered tests were reported as per the product inserts. The microneutralization assay is described by Walker et al.[1] The neutralization assay was considered the definitive release test. For a donation to be considered suitable for clinical CP the

donation had to test positive on one of the two screening tests and have a neutralizing antibody titer greater or equal to 1:80.

**Canada**

Supply of convalescent plasma to local sites was coordinated and managed by two blood services (Héma-Québec and Canadian Blood Services).

*Donor selection*

In addition to standard criteria for plasma donation set forth in the Manuals of donor selection in use at Héma-Québec or Canadian Blood Services, donor criteria included: 1) a prior diagnosis of COVID-19 documented by a PCR test at time of infection or by positive anti-SARS-CoV-2 serology following infection, 2) males or females with no pregnancy history or with negative anti-HLA antibodies, 3) At least 6 days since last plasma donation, 4) provided informed consent and 5) a complete resolution of symptoms at least 14 days prior to donation. The median time between symptom onset or test-positivity and donation was 57 days (IQR 46-73) (data from Héma-Québec).

*Plasma manufacture*

Plasma was collected by apheresis from qualified donors who have consented to donate. Donors donated 750 mL of plasma as three 250 mL units.

*CP testing*

All CP donations collected in Canada were tested for SARS-CoV-2 antibodies using ELISA test targeting the receptor binding domain (RBD) of the spike (S) glycoprotein.[2] They were also tested for IgG against the full spike protein with flow cytometry,[3] for antibody-dependent cell cytotoxicity (ADCC) activity against the spike protein[4] and for live virus neutralization using plaque reduction neutralization test (PRNT).[5] At Héma-Québec, only donations with at least three standard deviations above the mean value of the anti-RBD ELISA signal, obtained with SARS-CoV-2 negative plasma (pool of plasma samples collected before 2020) at a 1:100 plasma dilution, were supplied for clinical trial use. At Canadian Blood Services, only donations with a PRNT50 titre of ≥1:160 were supplied for clinical trial use.

**United Kingdom**

Supply of convalescent plasma to local sites was coordinated and managed by the four UK blood services (NHS Blood and Transplant (NHSBT), Northern Ireland Blood Transfusion Service (NIBTS), Scottish National Blood Transfusion Service (SNBTS), and Welsh Blood Service (WBS)). There was a UK-wide collaboration to ensure production of convalescent plasma was consistent across all devolved nations.

*Donor selection*

All donors met the standard donor selection guidelines in the UK (https://www.transfusionguidelines.org/dsg). Only male plasma or plasma from female donors who had been tested and were eligible to donate apheresis platelets were used to reduce the risk of TRALI (HLA & HNA negative).[6] In addition to these standard donor selection criteria, donor criteria included 1) self-reported symptoms consistent with COVID-19 or a laboratory confirmed SARS-CoV-2 infection; and 2) donation at least 28 days following symptom resolution. The median time between symptom onset or test-positivity and donation was 54 days (IQR 41-78) (data from NHSBT).

*Plasma manufacture*

CP was collected via apheresis with a volume of 275mls +/-10%. Additional details about manufacture are available at https://www.transfusionguidelines.org/red-book/annex-3/a3-7-convalescent-plasma-covid-19-ffp-leucocyte-depleted.

*CP testing*

All CP donations collected in the UK were routinely tested for SARS-CoV-2 IgG antibodies using the EUROimmun IgG ELISA targeting the spike (S) glycoprotein (PerkinElmer, London, UK). Only donations with signal to cutoff (S/CO) ratio of 6.0 or above were supplied for clinical trial use, as this cut-off was previously demonstrated to be associated with the presence of neutralising antibody titres of ≥1:100.[7] Among first-time donors with PCR-confirmed infection not leading to hospitalisation, 1,382/5,906 (23%) had Euroimmun S/CO values above this threshold (data from NHSBT).

**United States**

*Donor selection*

CP was collected from individuals in the community with documented COVID-19 infection who were more than 28 days from last symptoms. Prospective donors were screened at UPMC for both IgA and IgG antibodies against the COVID-19 spike protein using an in-house validated version of the Euroimmun assay.[8] This assay is considered positive if reactive at an initial dilution of 1:100, with the semiquantitative readout correlating with neutralizing antibodies titers as demonstrated in vitro.[9] Those individuals with IgG antibodies were referred for plasma donation at the local licensed blood center (Vitalant, Scottsdale AZ).

*Plasma manufacture*

Donors who met all volunteer donor eligibility criteria had 2-4 units of plasma collected by plasmapheresis and labelled according to FDA emergency guidance. Donated CP units approximately 200-250 ml were tagged and sent to the UPMC transfusion service. All blood types were available. Group A units were tested for anti-B and considered "Low Titer- A" units if the titer was <100. These units could be used for patients of any blood type.

*CP testing*
Two CP units were randomly distributed to REMAP-consented patients with proven COVID-19 positivity and admitted for worsening respiratory status throughout the UPMC hospital system, using an additional consent "Expanded Access to Convalescent Plasma for the Treatment of Patients with COVID-19"; IND 19832.

#### Measurement of participant baseline SARS-CoV-2 serostatus

**Australia**

The presence of SARS-CoV-2 neutralising antibodies in serum samples collected at randomization was determined using the surrogate virus neutralisation test (sVNT) and a microneutralisation assay. For the sVNT, patient serum samples were diluted 1:9 and pre-incubated with a horseradish peroxidase-conjugated RBD (HRP-RBD). This mixture was then added to wells of a 96-well plate coated with ACE2. The extent to which the test plasma inhibited RBD binding to ACE2 was measured by optical density in relation to positive and negative internal controls, with inhibition above 20% being considered positive.[10] For the microneutralisation assay, SARS-CoV-2 isolates hCoV/Australia/VIC01/2020[11] and hCoV/Australia/VIC2089/2020 were passaged in Vero cells and aliquots stored at −80 °C. Human plasma was heat inactivated at 56 °C for 30 min prior to use and centrifuged at 10,000 xg for 5 min. Plasma was serially diluted in MEM medium, followed by the addition of 100 TCID50of SARS-CoV-2 in MEM/0.5% BSA and incubation at room temperature for 45 min. Vero cells were washed twice with serum-free MEM before the addition of MEM containing 1 µg/ml of TPCK trypsin. Vero cells were then inoculated in quadruplicate with the plasma:virus mixture and incubated at 37 °C and 5% CO2 for 3 days. Cytopathic effect was scored, and the neutralising antibody titer was calculated using the Reed–Muench method.[12,13]

**United Kingdom**

The presence of SARS-CoV-2 IgG antibodies in serum samples taken at randomization was determined using a validated high throughput ELISA, the Oxford immunoassay. This is an indirect ELISA, measuring serum IgG against trimeric spike protein using a horseradish peroxidase-linked anti-human IgG antibody as the secondary. Readouts are measured as a fluorescent signal, normalised to standard units by calibrating to a dilution series of NHSBT plasma controls and a monoclonal antibody (CR3022) run on each plate. A full description of the assay and its evaluation can be found in the report by the National SARS-CoV-Serology Assay Evaluation Group.[14]

##### United States

Enrolled patients at UPMC had plasma isolated by density gradient centrifugation and frozen until use at -80°C. The EUROIMMUN Anti-SARS-CoV-2 IgG enzyme-linked immunosorbent assay was used for the detection of antibodies to Spike protein SARS-CoV-2 at a dilution of 1/100 according to manufacturer's instructions (PerkinElmer, NJ, USA).

#### Measurement of participant baseline SARS-CoV-2 PCR status

##### United Kingdom

SARS-CoV-2 RNA was detected and quantified by real-time reverse transcriptase polymerase chain reaction (RT-PCR) in baseline nasopharyngeal and/or oropharyngeal samples as previously described.[15] Viral RNA was first extracted using the QIAamp Viral RNA Mini Kit (Qiagen), and then amplified using the Quantitect Probe RT-PCR (Qiagen) with established primers targeting the nucleoprotein gene. A standard curve of NIBSC RNA research reagent 19/304 spanning from 10 copies/reaction to 10,000 copies/reaction was included with each test run to allow intra-assay standardisation and viral load quantification. As cycle threshold (Ct) values above 35 were reported as SARS-CoV-2 RNA not detected corresponding to an RNA level of 10 IU/ml and were hence treated as negative for analysis purposes. All Ct values below 35 were reported as positive.

#### Treatment allocation and interventions

##### Available Interventions

The Immunoglobulin Domain contained three interventions, convalescent plasma at randomization, delayed convalescent plasma (if clinical deterioration) and no immunoglobulin against SARS-CoV-2 (referred to as the control group in the main manuscript). The delayed convalescent plasma intervention was only available in the United States.

##### Convalescent plasma at randomization

Patients assigned to convalescent plasma received two adult units of ABO compatible convalescent plasma (total volume approximately 550ml ± 150ml) within 48 hours of randomization. The first unit was given on the first day of the study. If the patient had no serious adverse reaction to the transfusion the second unit of convalescent plasma was administered with a minimum of 12 hours between transfusions, which was to allow appropriate assessment of adverse reactions to the initial transfusion. Both transfusions were to be given within 48 hours from randomization

**No immunoglobulin to SARS-CoV-2 (control)**

Patients assigned to this intervention did not receive any preparation of immunoglobulin intended to neutralize SARS-CoV-2 (convalescent plasma, hyperimmune globulin or monoclonal antibodies) during the index hospitalization. All other aspects of care that were not specified within the platform (for example if enrolled in another domain) were according to local practice determined by the treating clinician.

**Delayed convalescent plasma**

This intervention was only available at sites participating in the United States. Patients were assigned to no convalescent plasma, however could receive convalescent plasma after 48 hours if they failed to improve or clinically deteriorated. Convalescent plasma was administered as outlined under the convalescent plasma at randomization section above.

#### Secondary outcomes

**REMAP-CAP Core Protocol Secondary Outcomes:**

All-cause mortality censored at 90 days;

ICU outcomes:

- ICU mortality censored at 90 days;
- ICU Length of stay (LOS) censored at 90 days;
- Ventilator free days (VFDs) censored at 28 days;
- Organ failure free days (OFFDs) censored at 28 days;
- Proportion of intubated participants who receive a tracheostomy censored at 28 days;

Hospital outcomes:

- Hospital LOS censored 90 days after enrolment;
- Destination at time of hospital discharge (characterized as home, rehabilitation hospital, nursing home or long-term care facility, or another acute hospital);
- Readmission to the index ICU during the index hospitalization in the 90 days following enrolment

Ventilator- and organ failure-free days are calculated by counting the number of days that the participant is not ventilated or has no organ failure. If a participant dies during the hospitalization during which enrolment occurred, the number of VFDs or OFFDs will be set to zero. If the participant is discharged alive from hospital, the remainder of days censored at 90 days are counted as ventilator- or organ failure-free days.

The index hospital admission is defined as continuing while the participant is admitted to any healthcare facility or level of residence that provides a higher level of care than that corresponding to where the participant was residing prior to the hospital admission. Participants who have been and still are admitted to a healthcare facility 90 days after enrolment are coded as being alive.

**Immunoglobulin Domain Secondary Outcomes**

- All-cause mortality at 28 days
- Serious treatment-related adverse events
- Serious Adverse Events (SAE)
- Venous thromboembolic events at 90 days

**Secondary Outcomes prespecified in Statistical Analysis Plan**

- Progression to intubation and mechanical ventilation, extracorporeal membrane oxygenation (ECMO), or death
- World Health Organization (WHO) 8-point ordinal scale measured at day 14.
    - A modified WHO ordinal scale is used:
    - 0 + 1 + 2 = No longer hospitalized
    - 3 = Hospitalized, no oxygen therapy
    - 4 = Oxygen by mask or nasal prongs
    - 5 = Non-invasive ventilation or high-flow oxygen
    - 6 = Intubation and mechanical ventilation
    - 7 = Ventilation + additional organ support: vasopressors, renal replacement therapy (RRT), ECMO
    - 8 = Death

### 3. Supplementary figures

#### Figure S1

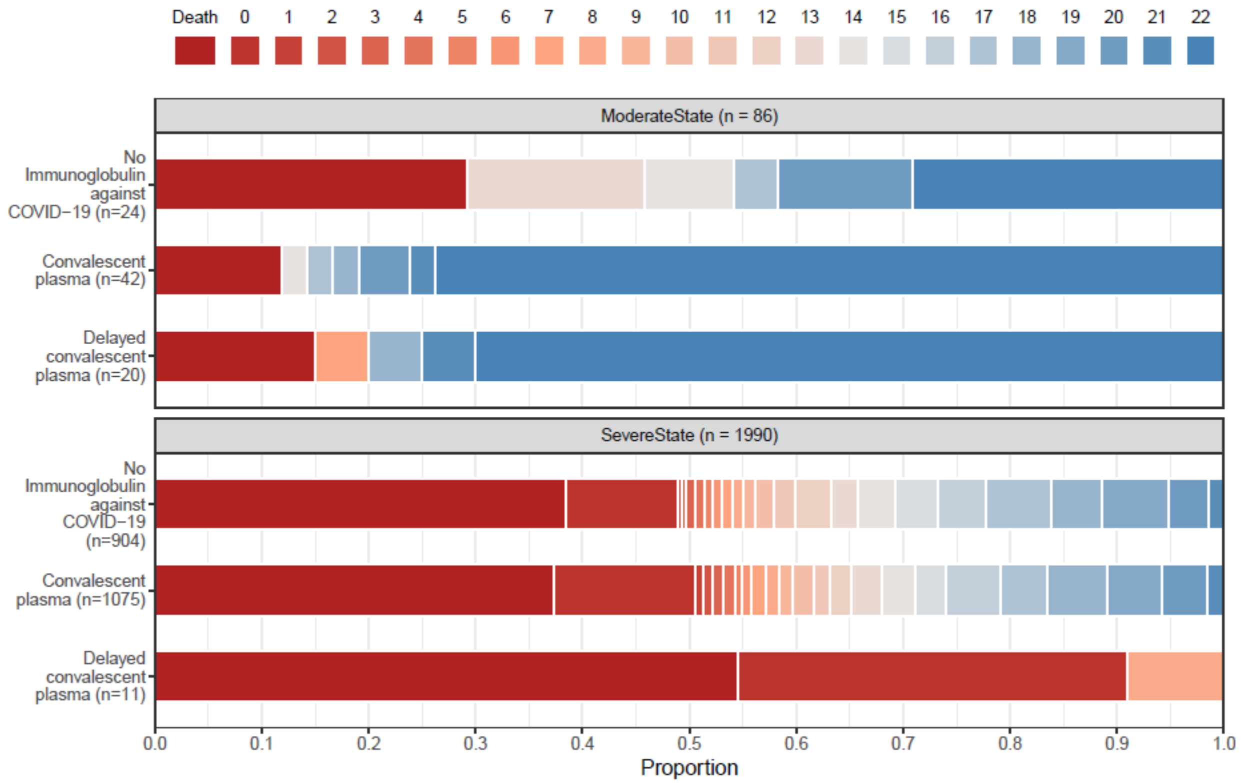

Figure Legend: Organ support–free days as horizontally stacked proportions according to intervention group. Red represents worse outcomes, and blue represents better outcomes. For the median adjusted odds ratios from the primary analysis see Table S3.

#### Figure S2

| Study or Subgroup | Convalescent plasma Events | Convalescent plasma Total | Placebo or standard care Events | Placebo or standard care Total | Weight | Odds Ratio (Non-event) M-H, Random, 95% CI |
|---|---|---|---|---|---|---|
| **1.1.1 Antibody positive** | | | | | | |
| RECOVERY | 566 | 3022 | 495 | 2752 | 34.9% | 0.95 [0.83, 1.09] |
| REMAP-CAP | 190 | 597 | 135 | 409 | 8.6% | 1.06 [0.81, 1.38] |
| **Subtotal (95% CI)** | | **3619** | | **3161** | **43.6%** | **0.97 [0.86, 1.09]** |
| Total events | 756 | | 630 | | | |
| Heterogeneity: Tau$^2$ = 0.00; Chi$^2$ = 0.46, df = 1 (P = 0.50); I$^2$ = 0% | | | | | | |
| Test for overall effect: Z = 0.48 (P = 0.63) | | | | | | |
| **1.1.2 Antibody negative** | | | | | | |
| RECOVERY | 626 | 1982 | 549 | 1626 | 31.9% | 1.10 [0.96, 1.27] |
| REMAP-CAP | 130 | 270 | 78 | 148 | 3.9% | 1.20 [0.80, 1.79] |
| **Subtotal (95% CI)** | | **2252** | | **1774** | **35.7%** | **1.11 [0.98, 1.27]** |
| Total events | 756 | | 627 | | | |
| Heterogeneity: Tau$^2$ = 0.00; Chi$^2$ = 0.15, df = 1 (P = 0.70); I$^2$ = 0% | | | | | | |
| Test for overall effect: Z = 1.61 (P = 0.11) | | | | | | |
| **1.1.3 Antibody unknown** | | | | | | |
| RECOVERY | 206 | 791 | 364 | 1382 | 15.7% | 1.02 [0.83, 1.24] |
| REMAP-CAP | 81 | 205 | 132 | 348 | 5.0% | 0.94 [0.66, 1.33] |
| **Subtotal (95% CI)** | | **996** | | **1730** | **20.7%** | **1.00 [0.84, 1.18]** |
| Total events | 287 | | 496 | | | |
| Heterogeneity: Tau$^2$ = 0.00; Chi$^2$ = 0.16, df = 1 (P = 0.69); I$^2$ = 0% | | | | | | |
| Test for overall effect: Z = 0.05 (P = 0.96) | | | | | | |
| **Total (95% CI)** | | **6867** | | **6665** | **100.0%** | **1.03 [0.95, 1.11]** |
| Total events | 1799 | | 1753 | | | |
| Heterogeneity: Tau$^2$ = 0.00; Chi$^2$ = 3.18, df = 5 (P = 0.67); I$^2$ = 0% | | | | | | |
| Test for overall effect: Z = 0.62 (P = 0.53) | | | | | | |
| Test for subgroup differences: Chi$^2$ = 2.42, df = 2 (P = 0.30), I$^2$ = 17.3% | | | | | | |

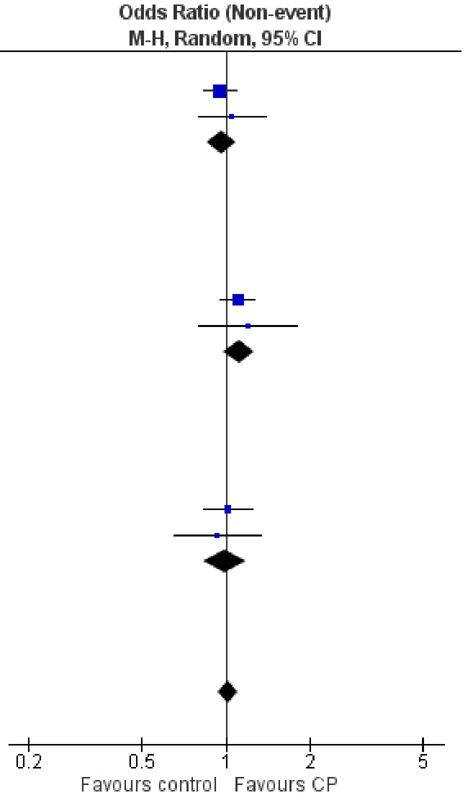

Figure Legend: Meta-analysis of studies reporting in-hospital or 28-day survival in antibody negative and antibody positive patients. Analysis updated with data from REMAP-CAP based on original analysis within the Cochrane review[16]

### 4. Supplementary Tables

#### Table S1. Participant Characteristics at Baseline*

| | **Convalescent Plasma** | | | **Delayed convalescent plasma** | | | **Control [†]** | | | **All patients [‡]** |
|---|---|---|---|---|---|---|---|---|---|---|
| | **All (*n* = 1120)** | **Moderate (n = 42)** | **Severe (n = 1078)** | **All (*n* = 31)** | **Moderate (n = 20)** | **Severe (n = 11)** | **All (*n* = 933)** | **Moderate (n = 24)** | **Severe (n = 909)** | **All (*n* = 2084)** |
| **Age, years** | | | | | | | | | | |
| Mean (SD) | 60.3 (12.8) | 62.1 (15.2) | 60.2 (12.7) | 61.1 (17.5) | 57.2 (17.4) | 68.3 (15.9) | 60.2 (13.1) | 59.7 (13.6) | 60.2 (13.1) | 60.3 (13.0) |
| **Male sex** | | | | | | | | | | |
| N (%) | 753 (67.2) | 26 (61.9) | 727 (67.4) | 18 (58.1) | 10 (50.0) | 8 (72.7) | 633 (67.8) | 15 (62.5) | 618 (68.0) | 1404 (67.4) |
| **Race / Ethnicity § - n / N (%)** | | | | | | | | | | |
| White | 759 / 1011 (75.1) | 28 / 35 (80.0) | 731 / 976 (74.9) | 26 / 30 (86.7) | 17 / 19 (89.5) | 9 / 11 (81.8) | 631 / 846 (74.6) | 12 / 14 (85.7) | 619 / 832 (74.4) | 1416 / 1887 (75.0) |
| Asian | 145 / 1011 (14.3) | 1 / 35 (2.9) | 144 / 976 (14.8) | 0 / 30 (0.0) | 0 / 19 (0.0) | 0 / 11 (0.0) | 135 / 846 (16.0) | 2 / 14 (14.3) | 133 / 832 (16.0) | 280 / 1887 (14.8) |
| Black | 56 / 1011 (5.5) | 5 / 35 (14.3) | 51 / 976 (5.2) | 4 / 30 (13.3) | 2 / 19 (10.5) | 2 / 11 (18.2) | 38 / 846 (4.5) | 0 / 14 (0.0) | 38 / 832 (4.6) | 98 / 1887 (5.2) |
| Mixed | 16 / 1011 (1.6) | 0 / 35 (0.0) | 16 / 976 (1.6) | 0 / 30 (0.0) | 0 / 19 (0.0) | 0 / 11 (0.0) | 8 / 846 (0.9) | 0 / 14 (0.0) | 8 / 832 (1.0) | 24 / 1887 (1.3) |
| Other | 35 / 1011 (3.5) | 1 / 35 (2.9) | 34 / 976 (3.5) | 0 / 30 (0.0) | 0 / 19 (0.0) | 0 / 11 (0.0) | 34 / 846 (4.0) | 0 / 14 (0.0) | 34 / 832 (4.1) | 69 / 1887 (3.7) |
| **Body-mass index, ¶ kg/m²** | | | | | | | | | | |
| Median | 30.8 | 30.9 | 30.8 | 32.4 | 32.3 | 33.3 | 31.1 | 30.3 | 31.1 | 30.9 |
| IQR | (26.9 - 35.6) | (26.0 - 35.1) | (26.9 - 35.6) | (28.7 - 39.7) | (29.2 - 37.4) | (29.1 - 41.8) | (26.9 - 36.5) | (25.0 - 34.6) | (26.9 - 36.6) | (26.9 - 36.1) |
| N | (*n* = 995) | (*n* = 38) | (*n* = 957) | (*n* = 31) | (*n* = 20) | (*n* = 11) | (*n* = 832) | (*n* = 18) | (*n* = 814) | (*n* = 1858) |
| **APACHE II score ‖ - median (IQR)** | | | | | | | | | | |
| Median | 13.0 | 13.0 | 13.0 | --- | --- | --- | 12.0 | 11.0 | 12.0 | 13.0 |
| IQR | (8.0 - 19.0) | (7.0 - 17.5) | (8.0 - 19.0) | | | | (8.0 - 19.0) | (8.0 - 13.0) | (8.0 - 19.0) | (8.0 - 19.0) |
| N | (*n* = 1059) | (*n* = 15) | (*n* = 1044) | (*n* = 0) | (*n* = 0) | (*n* = 0) | (*n* = 911) | (*n* = 23) | (*n* = 888) | (*n* = 1970) |
| **Confirmed SARS-CoV-2 infection [**]** | | | | | | | | | | |
| n / N (%) | 1034 / 1120 (92.3) | 38 / 42 (90.5) | 996 / 1078 (92.4) | 31 / 31 (100.0) | 20 / 20 (100.0) | 11 / 11 (100.0) | 855 / 931 (91.8) | 16 / 24 (66.7) | [illegible]39 / 907 (92.5) | 1920 / 2082 (92.2) |
| **PCR positive at randomization [††]** | | | | | | | | | | |

| | | | | | | | | | | |
|---|---|---|---|---|---|---|---|---|---|---|
| n / N (%) | 683 / 854 (80.0) | 7 / 9 (77.8) | 676 / 845 (80.0) | 6 / 6 (100.0) | 3 / 3 (100.0) | 3 / 3 (100.0) | 500 / 611 (94.5 ) | 11 / 12 (81.6) | 489 / 599 (81.6) | 1189 / 1471 (80.8) |
| **Recipient antibody at randomization- n / N (%)** | | | | | | | | | | |
| Negative | 283 / 901 (31.4) | 12 / 27 (44.4) | 271 / 874 (31.0) | 9 / 15 (60.0) | 4 / 7 (57.1) | 5 / 8 (62.5) | 153 / 568 (26.9) | 4 / 10 (40.0) | 149 / 558 (26.7) | 445 / 1484 (30.0) |
| Positive | 618 / 901 (68.6) | 15 / 27 (55.6) | 603 / 874 (69.0) | 6 / 15 (40.0) | 3 / 7 (42.9) | 3 / 8 (37.5) | 415 / 568 (73.1) | 6 / 10 (60.0) | 409 / 558 (73.3) | 1039 / 1484 (70.0) |
| **Preexisting condition - n / N (%)** | | | | | | | | | | |
| Diabetes | 352 / 1120 (31.4) | 13 / 42 (31.0) | 339 / 1078 (31.4) | 12 / 31 (38.7) | 6 / 20 (30.0) | 6 / 11 (54.5) | 279 / 931 (30.0) | 11 / 24 (45.8) | 268 / 907 (29.5) | 643 / 2082 (30.9) |
| Respiratory disease | 257 / 1120 (22.9) | 12 / 42 (28.6) | 245 / 1078 (22.7) | 7 / 31 (22.6) | 5 / 20 (25.0) | 2 / 11 (18.2) | 220 / 931 (23.6) | 4 / 24 (16.7) | 216 / 907 (23.8) | 484 / 2082 (23.2) |
| Asthma/COPD | 216 / 1120 (19.3) | 10 / 42 (23.8) | 206 / 1078 (19.1) | 7 / 31 (22.6) | 5 / 20 (25.0) | 2 / 11 (18.2) | 180 / 931 (19.3) | 2 / 24 (8.3) | 178 / 907 (19.6) | 403 / 2082 (19.4) |
| Other | 54 / 1120 (4.8) | 3 / 42 (7.1) | 51 / 1078 (4.7) | 0 / 31 (0.0) | 0 / 20 (0.0) | 0 / 11 (0.0) | 50 / 931 (5.4) | 2 / 24 (8.3) | 48 / 907 (5.3) | 104 / 2082 (5.0) |
| Kidney disease | 111 / 1041 (10.7) | 4 / 41 (9.8) | 107 / 1000 (10.7) | 4 / 31 (12.9) | 4 / 20 (20.0) | 0 / 11 (0.0) | 85 / 858 (9.9) | 2 / 21 (9.5) | 83 / 837 (9.9) | 200 / 1930 (10.4) |
| Severe cardiovascular disease | 97 / 1095 (8.9) | 1 / 42 (2.4) | 96 / 1053 (9.1) | 0 / 31 (0.0) | 0 / 20 (0.0) | 0 / 11 (0.0) | 68 / 914 (7.4) | 1 / 24 (4.2) | 67 / 890 (7.5) | 165 / 2040 (8.1) |
| Any immuno-suppressive condition | 68 / 1084 (6.3) | 1 / 18 (5.6) | 67 / 1066 (6.3) | --- | --- | --- | 61 / 931 (6.6) | 1 / 24 (4.2) | 60 / 907 (6.6) | 129 / 2015 (6.4) |
| Cancer | 15 / 1084 (1.4) | 1 / 18 (5.6) | 14 / 1066 (1.3) | --- | --- | --- | 14 / 931 (1.5) | 0 / 24 (0.0) | 14 / 907 (1.5) | 29 / 2015 (1.4) |
| Chronic immuno-suppressive therapy | 47 / 1120 (4.2) | 0 / 42 (0.0) | 47 / 1078 (4.4) | 0 / 31 (0.0) | 0 / 20 (0.0) | 0 / 11 (0.0) | 44 / 931 (4.7) | 1 / 24 (4.2) | 43 / 907 (4.7) | 91 / 2082 (4.4) |
| Other | 39 / 1083 (3.6) | 0 / 17 (0.0) | 39 / 1066 (3.7) | --- | --- | --- | 27 / 931 (2.9) | 1 / 24 (4.2) | 26 / 907 (2.9) | 66 / 2014 (3.3) |
| Liver cirrhosis / failure | 9 / 1095 (0.8) | 1 / 42 (2.4) | 8 / 1053 (0.8) | 0 / 31 (0.0) | 0 / 20 (0.0) | 0 / 11 (0.0) | 5 / 914 (0.5) | 0 / 24 (0.0) | 5 / 890 (0.6) | 14 / 2040 (0.7) |
| **Time to enrollment - median (IQR)** | | | | | | | | | | |
| From hospital admission - days | 1.8 (1.0 - 3.2) | 1.0 (0.8 - 1.9) | 1.8 (1.0 - 3.3) | 1.3 (0.9 - 1.7) | 1.4 (0.9 - 1.7) | 1.3 (0.8 - 1.7) | 1.7 (0.9 - 3.4) | 1.4 (0.7 - 2.1) | 1.7 (0.9 - 3.5) | 1.7 (1.0 - 3.2) |

| | | | | | | | | | | |
|---|---|---|---|---|---|---|---|---|---|---|
| From ICU admission - hours | 17.8 (10.4 - 23.5) | 21.6 (14.1 - 29.4) | 17.7 (10.2 - 23.5) | 22.4 (15.5 - 34.4) | 20.8 (20.6 - 27.4) | 24.1 (13.0 - 35.5) | 17.2 (10.5 - 23.2) | 15.1 (7.6 - 22.6) | 17.2 (10.6 - 23.2) | 17.5 (10.4 - 23.5) |
| **Acute respiratory support - n (%)** | | | | | | | | | | |
| None / supplemental oxygen only | 44 (3.9) | 42 (100.0) | 2 (0.2) | 20 (64.5) | 20 (100.0) | 0 (0.0) | 25 (2.7) | 24 (100.0) | 1 (0.1) | 89 (4.3) |
| High-flow nasal cannula | 225 (20.1) | 0 (0.0) | 225 (20.9) | 6 (19.4) | 0 (0.0) | 6 (54.5) | 211 (22.6) | 0 (0.0) | 211 (23.2) | 442 (21.2) |
| Noninvasive ventilation only | 493 (44.0) | 0 (0.0) | 493 (45.7) | 0 (0.0) | 0 (0.0) | 0 (0.0) | 407 (43.6) | 0 (0.0) | 407 (44.8) | 900 (43.2) |
| Invasive mechanical ventilation | 356 (31.8) | 0 (0.0) | 356 (33.0) | 5 (16.1) | 0 (0.0) | 5 (45.5) | 289 (31.0) | 0 (0.0) | 289 (31.8) | 650 (31.2) |
| ECMO | 2 (0.2) | 0 (0.0) | 2 (0.2) | 0 (0.0) | 0 (0.0) | 0 (0.0) | 1 (0.1) | 0 (0.0) | 1 (0.1) | 3 (0.1) |
| **Vasopressor support** | | | | | | | | | | |
| n (%) | 207 (18.5) | 0 (0.0) | 207 (19.2) | 2 (6.5) | 0 (0.0) | 2 (18.2) | 175 (18.8) | 0 (0.0) | 175 (19.3) | 384 (18.4) |
| **Acute kidney replacement therapy** | | | | | | | | | | |
| n / N (%) | 11 / 1115 (1.0) | 0 (0.0) | 11 / 1073 (1.0) | 0 / 31 (0.0) | 0 (0.0) | 0 / 11 (0.0) | 6 / 928 (0.6) | 0 (0.0) | 6 / 904 (0.7) | 17 / 2074 (0.8) |
| **$PaO_2$ / $FiO_2$** | | | | | | | | | | |
| Median<br>IQR<br>N | 118<br>(91 - 161)<br>(*n* = 979) | 191<br>(178 - 246)<br>(*n* = 10) | 117<br>(91 - 159)<br>(*n* = 969) | 129<br>(82 - 176)<br>(*n* = 11) | 116<br>(109 - 123)<br>(*n* = 2) | 150<br>(81 - 198)<br>(*n* = 9) | 116<br>(91 - 155)<br>(*n* = 829) | 142<br>(106 - 187)<br>(*n* = 17) | 115<br>(90 - 154)<br>(*n* = 812) | 117<br>(91 - 157)<br>(*n* = 1819) |
| **Glasgow coma scale score ‡‡** | | | | | | | | | | |
| Median<br>IQR<br>N | 15.0<br>(15.0 - 15.0)<br>(*n* = 1011) | 15.0<br>(15.0 - 15.0)<br>(*n* = 17) | 15.0<br>(15.0 - 15.0)<br>(*n* = 994) | ---<br><br>(*n* = 0) | ---<br><br>(*n* = 0) | ---<br><br>(*n* = 0) | 15.0<br>(15.0 - 15.0)<br>(*n* = 869) | 15.0<br>(15.0 - 15.0)<br>(*n* = 24) | 15.0<br>(15.0 - 15.0)<br>(*n* = 845) | 15.0<br>(15.0 - 15.0)<br>(*n* = 1880) |
| **Median laboratory values (IQR) §§** | | | | | | | | | | |
| C-reactive protein, μg/mL | | | | | | | | | | |
| Median<br>IQR<br>N | 122<br>(69 - 195)<br>(*n* = 921) | 155<br>(76 - 198)<br>(*n* = 12) | 121<br>(68 - 195)<br>(*n* = 909) | 45<br>(26 - 117)<br>(*n* = 3) | 26<br>(16 - 35)<br>(*n* = 2) | 190<br>(190 - 190)<br>(*n* = 1) | 124<br>(72 - 195)<br>(*n* = 769) | 104<br>(86 - 146)<br>(*n* = 14) | 124<br>(72 - 195)<br>(*n* = 755) | 123<br>(70 - 195)<br>(*n* = 1693) |
| D-dimer, ng/mL | | | | | | | | | | |
| Median<br>IQR<br>N | 954<br>(456 - 2290)<br>(*n* = 622) | 1309<br>(835 - 1870)<br>(*n* = 11) | 941<br>(454 - 2305)<br>(*n* = 611) | 470<br>(445 - 1210)<br>(*n* = 3) | 470<br>(445 - 1210)<br>(*n* = 3) | ---<br><br>(*n* = 0) | 868<br>(417 - 1895)<br>(*n* = 529) | 832<br>(468 - 1519)<br>(*n* = 10) | 868<br>(415 - 1902)<br>(*n* = 519) | 891<br>(440 - 2060)<br>(*n* = 1154) |
| Ferritin, ng/mL | | | | | | | | | | |
| Median<br>IQR<br>N | 1028<br>(545 - 1815)<br>(*n* = 568) | 1218<br>(365 - 3490)<br>(*n* = 5) | 1027<br>(546 - 1813)<br>(*n* = 563) | 334<br>(274 - 394)<br>(*n* = 2) | 334<br>(274 - 394)<br>(*n* = 2) | ---<br><br>(*n* = 0) | 988<br>(525 - 1761)<br>(*n* = 471) | 1555<br>(835 - 1950)<br>(*n* = 6) | 986<br>(525 - 1760)<br>(*n* = 465) | 1011<br>(530 - 1807)<br>(*n* = 1041) |

| | | | | | | | | | | |
|---|---|---|---|---|---|---|---|---|---|---|
| Neutrophils, x10$^9$/L | | | | | | | | | | |
| Median<br>IQR<br>N | 7.9<br>(5.5 - 11.0)<br>(*n* = 1021) | 5.7<br>(4.2 - 8.2)<br>(*n* = 21) | 7.9<br>(5.5 - 11.0)<br>(*n* = 1000) | 2.6<br>(2.0 - 3.2)<br>(*n* = 4) | 2.6<br>(2.0 - 3.2)<br>(*n* = 4) | ---<br><br>(*n* = 0) | 7.9<br>(5.4 - 11.0)<br>(*n* = 858) | 5.2<br>(4.0 - 9.4)<br>(*n* = 23) | 7.9<br>(5.5 - 11.0)<br>(*n* = 835) | 7.8<br>(5.4 - 11.0)<br>(*n* = 1883) |
| Lymphocytes, x10$^9$/L | | | | | | | | | | |
| Median<br>IQR<br>N | 0.7<br>(0.5 - 1.0)<br>(*n* = 1019) | 0.8<br>(0.4 - 1.2)<br>(*n* = 22) | 0.7<br>(0.5 - 1.0)<br>(*n* = 997) | 1.0<br>(0.8 - 1.3)<br>(*n* = 4) | 1.0<br>(0.8 - 1.3)<br>(*n* = 4) | ---<br>(*n* = 0) | 0.7<br>(0.5 - 0.9)<br>(*n* = 856) | 0.6<br>(0.4 - 0.8)<br>(*n* = 23) | 0.7<br>(0.5 - 0.9)<br>(*n* = 833) | 0.7<br>(0.5 - 1.0)<br>(*n* = 1879) |
| Platelet count, x10$^9$/L | | | | | | | | | | |
| Median<br>IQR<br>N | 234<br>(176 - 302)<br>(*n* = 1108) | 223<br>(151 - 266)<br>(*n* = 40) | 235<br>(177 - 303)<br>(*n* = 1068) | 199<br>(170 - 264)<br>(*n* = 26) | 183<br>(0 - 220)<br>(*n* = 17) | 271<br>(181 - 334)<br>(*n* = 9) | 243<br>(182 - 306)<br>(*n* = 923) | 247<br>(6 - 277)<br>(*n* = 24) | 242<br>(182 - 307)<br>(*n* = 899) | 236<br>(178 - 305)<br>(*n* = 2057) |
| Lactate, mmol/L | | | | | | | | | | |
| Median<br>IQR<br>N | 1.3<br>(1.0 - 1.7)<br>(*n* = 980) | 1.4<br>(0.9 - 1.7)<br>(*n* = 18) | 1.3<br>(1.0 - 1.7)<br>(*n* = 962) | 1.3<br>(1.2 - 1.7)<br>(*n* = 17) | 1.2<br>(1.0 - 1.5)<br>(*n* = 8) | 1.6<br>(1.2 - 1.7)<br>(*n* = 9) | 1.3<br>(1.0 - 1.7)<br>(*n* = 831) | 1.0<br>(0.9 - 1.4)<br>(*n* = 21) | 1.3<br>(1.0 - 1.7)<br>(*n* = 810) | 1.3<br>(1.0 - 1.7)<br>(*n* = 1828) |
| Creatinine, mg/dL | | | | | | | | | | |
| Median<br>IQR<br>N | 0.8<br>(0.7 - 1.1)<br>(*n* = 1110) | 0.9<br>(0.7 - 1.0)<br>(*n* = 40) | 0.8<br>(0.7 - 1.1)<br>(*n* = 1070) | 1.0<br>(0.8 - 1.3)<br>(*n* = 28) | 1.0<br>(0.7 - 1.3)<br>(*n* = 17) | 1.1<br>(0.8 - 1.3)<br>(*n* = 11) | 0.9<br>(0.7 - 1.2)<br>(*n* = 920) | 0.9<br>(0.8 - 1.1)<br>(*n* = 23) | 0.8<br>(0.7 - 1.2)<br>(*n* = 897) | 0.8<br>(0.7 - 1.1)<br>(*n* = 2058) |
| Bilirubin, mg/dL | | | | | | | | | | |
| Median<br>IQR<br>N | 0.5<br>(0.4 - 0.8)<br>(*n* = 1053) | 0.4<br>(0.3 - 0.6)<br>(*n* = 25) | 0.5<br>(0.4 - 0.8)<br>(*n* = 1028) | 0.5<br>(0.4 - 0.7)<br>(*n* = 19) | 0.5<br>(0.4 - 0.6)<br>(*n* = 11) | 0.5<br>(0.4 - 0.8)<br>(*n* = 8) | 0.5<br>(0.4 - 0.8)<br>(*n* = 889) | 0.5<br>(0.4 - 0.7)<br>(*n* = 18) | 0.5<br>(0.4 - 0.8)<br>(*n* = 871) | 0.5<br>(0.4 - 0.8)<br>(*n* = 1961) |
| **Received therapies - n (%)** | | | | | | | | | | |
| **Steroids** | | | | | | | | | | |
| At randomization | 976 (87.1) | 33 (78.6) | 943 (87.5) | 27 (87.1) | 16 (80.0) | 11 (100.0) | 810 (86.8) | 22 (91.7) | 788 (86.7) | 1813 (87.0) |
| Within 48 hrs of randomization | 1051 (93.8) | 37 (88.1) | 1014 (94.1) | 29 (93.5) | 18 (90.0) | 11 (100.0) | 867 (92.9) | 22 (91.7) | 845 (93.0) | 1947 (93.4) |
| **Remdesivir** | | | | | | | | | | |
| At randomization | 419 (37.4) | 16 (38.1) | 403 (37.4) | 18 (58.1) | 13 (65.0) | 5 (45.5) | 307 (32.9) | 4 (16.7) | 303 (33.3) | 744 (35.7) |
| Within 48 hrs of randomization | 512 (45.7) | 21 (50.0) | 491 (45.5) | 25 (80.6) | 18 (90.0) | 7 (63.6) | 403 (43.2) | 5 (20.8) | 398 (43.8) | 940 (45.1) |
| **Immunomodulators** | | | | | | | | | | |
| Any | | | | | | | | | | |
| At randomization | 39 (3.5) | 0 (0.0) | 39 (3.6) | 0 (0.0) | 0 (0.0) | 0 (0.0) | 31 (3.3) | 0 (0.0) | 31 (3.4) | 70 (3.4) |

| | | | | | | | | | | |
|---|---|---|---|---|---|---|---|---|---|---|
| Within 48 hrs of randomization | 425 (37.9) | 0 (0.0) | 425 (39.4) | 0 (0.0) | 0 (0.0) | 0 (0.0) | 351 (37.6) | 3 (12.5) | 348 (38.3) | 776 (37.2) |
| Tocilizumab | | | | | | | | | | |
| At randomization | 21 (1.9) | 0 (0.0) | 21 (1.9) | 0 (0.0) | 0 (0.0) | 0 (0.0) | 20 (2.1) | 0 (0.0) | 20 (2.2) | 41 (2.0) |
| Within 48 hrs of randomization | 241 (21.5) | 0 (0.0) | 241 (22.4) | 0 (0.0) | 0 (0.0) | 0 (0.0) | 210 (22.5) | 2 (8.3) | 208 (22.9) | 451 (21.6) |
| Sarilumab | | | | | | | | | | |
| At randomization | 9 (0.8) | 0 (0.0) | 9 (0.8) | 0 (0.0) | 0 (0.0) | 0 (0.0) | 9 (1.0) | 0 (0.0) | 9 (1.0) | 18 (0.9) |
| Within 48 hrs of randomization | 129 (11.5) | 0 (0.0) | 129 (12.0) | 0 (0.0) | 0 (0.0) | 0 (0.0) | 99 (10.6) | 0 (0.0) | 99 (10.9) | 228 (10.9) |

* Percentages may not sum to 100 because of rounding. SD denotes standard deviation; APACHE, Acute Physiology and Chronic Health Evaluation; COPD, chronic obstructive pulmonary disease; IQR, interquartile range; ECMO, extracorporeal membrane oxygenation.

† Control patients include all patient randomized to control who were also eligible to be randomized to convalescent plasma.

‡ All patients includes patient randomized to control (including where convalescent plasma was not a randomization option) and convalescent plasma.

§ Data collection not approved in Canada and continental Europe. “Other” includes “declined” and “multiple”.

¶ Body-mass index is the weight in kilograms divided by the square of the height in meters.

‖ Range 0 to 71, with higher scores indicating greater severity of illness.

** SARS-CoV2 infection was confirmed by respiratory tract polymerase chain reaction test.

†† Positive when the test was taken between 24 hours before and after randomization.

‡‡ Range 3 to 15, with higher scores indicating greater consciousness, using values closest to randomization but prior to the use of sedative agents.

§§ Values were from the sample collected closest to randomization, up to 8 hours prior to randomization. If no samples were collected up to 8 hours prior to time of randomization, the sample collected closest to the time of randomization up to 2 hours after randomization was used (other than $PaO_2$ / $FiO_2$ which was a pre-randomization value only)

#### Table S2. Randomisation into other unblinded domains

| Domain | Domain Arm | Convalescent plasma | Control |
|---|---|---|---|
| **Immune modulation** | Pooled IL-6ra | 395 | 327 |
| | Control | 108 | 95 |
| **Antiviral** | Combination therapy | 0 | 0 |
| | HCQ | 0 | 2 |
| | Lopinavir-ritonavir | 50 | 48 |
| | No antiviral | 78 | 68 |
| **Anticoagulation** | Therapeutic | 166 | 143 |
| | Thromboprophylaxis | 169 | 135 |
| **Steroid** | Shock-dependent corticosteroids | 3 | 2 |
| | Fixed-dose corticosteroids* | 1057 | 887 |
| | No steroids | 4 | 6 |

* This includes patients randomized after the closure of the steroid domain in the fixed-dose corticosteroids intervention

#### Table S3. Primary and Secondary Analyses of Primary Outcomes

| Analysis | Severe State<br>Convalescent plasma (N= 1078) | Delayed convalescent plasma (N = 11) | Control (N = 909) | Moderate State<br>Convalescent plasma (N= 42) | Delayed convalescent plasma (N = 20) | Control (N = 24) |
|---|---|---|---|---|---|---|
| **Primary Outcome** | | | | | | |
| **Organ support-free days (day 21)** | | | | | | |
| Median (IQR) | 0 (-1 to 16) | -1 (-1 to 0) | 3 (-1 to 16) | 22 (21.25 to 22) | 22 (20.25 to 22) | 14 (-1 to 22) |
| Adjusted OR - mean (SD) | 0.97 (0.08) | -* | 1 | 2.68 (5.19) | -* | 1 |
| - median (95% Credible Interval) | 0.97 (0.83 to 1.15) | -* | 1 | 1.58 (0.82 to 5.95) | -* | 1 |
| Probability of superiority to control, % | 37.8 | | | 86.3 | | |
| Probability of futility compared to control, % | 99.4 | | | 30.8 | | |
| **Subcomponents of organ support-free days** | | | | | | |
| In-hospital deaths, no./total number (%) | 401/1075 (37.3) | 6/11 (54.5) | 347/904 (38.4) | 5/42 (11.9) | 3/20 (15.0) | 7/24 (24.2) |
| Median no. of organ support-free days in survivors (IQR) | 14 (3 to 18) | 0 (0 to 0) | 14 (7 to 18) | 22 (22 to 22) | 22 (22 to 22) | 20 (14 to 22) |
| **Hospital Survival** | | | | | | |
| Adjusted OR - mean (SD) | 1.04 (0.11) | -* | 1 | 1.55 (1.15) | -* | 1 |
| - median (95% Credible Interval) | 1.04 (0.85 to 1.27) | -* | 1 | 1.20 (0.70 to 4.54) | -* | 1 |
| Probability of superiority to control, % | 64.5 | | | 75.6 | | |
| Probability of futility compared to control, % | 91.8 | | | 50.0 | | |
| **Secondary Analysis of OSFD, model restricted to Immunoglobulin Domain participants and any previously reported interventions and domains (n=3446)** | N = 1072† | N = 11 | N = 900 | | | |
| Adjusted OR - mean (SD) | 0.95 (0.08) | 0.52 (0.3) | 1 | -* | -* | -* |
| - median (95% Credible Interval) | 0.94 (0.81 to 1.11) | 0.46 (0.16 to 1.16) | 1 | -* | -* | -* |
| Probability of superiority to control, % | 24.3 | -* | | -* | -* | -* |
| **Secondary Analysis of OSFD, model restricted to Immunoglobulin Domain participants‡** | | | | | | |
| Adjusted OR - mean (SD) | 0.96 (0.08) | -* | 1 | -* | -* | -* |

| | | | | | | |
|---|---|---|---|---|---|---|
| - median (95% Credible Interval) | 0.95 (0.81 to 1.13) | -* | 1 | -* | -* | -* |
| Probability of superiority to control, % | 28.5 | -* | | -* | -* | -* |
| **Secondary Analysis of in-hospital survival, model restricted to Immunoglobulin Domain participants and any previously reported interventions and domains (n=3446)** | N = 1072† | N = 11 | N = 900 | | | |
| Adjusted OR - mean (SD) | 1.02 (0.10) | 0.81 (0.56) | 1 | -* | -* | -* |
| - median (95% Credible Interval) | 1.02 (0.83 to 1.24) | 0.67 (0.20 to 2.22) | 1 | -* | -* | -* |
| Probability of superiority to control, % | 55.7 | -* | | -* | -* | -* |
| **Secondary Analysis of OSFD, model restricted to Immunoglobulin Domain participants‡** | | | | | | |
| Adjusted OR - mean (SD) | 1.04 (0.11) | -* | 1 | -* | -* | -* |
| - median (95% Credible Interval) | 1.03 (0.84 to 1.27) | -* | 1 | -* | -* | -* |
| Probability of superiority to control, % | 61.9 | -* | | -* | -* | -* |

* Analysis not performed due to limited sample size. SD - standard deviation; CrI - credible interval; OR - odds ratio.

†All interventions and specified interactions, age, sex, site, time, Immunoglobulin Domain interventions: control, convalescent plasma, delayed convalescent plasma, Covid-19 Antiviral Therapy Domain interventions: control, lopinavir–ritonavir, hydroxychloroquine, combination therapy, Corticosteroid Domain interventions: no steroids, fixed-duration corticosteroids, and shock-dependent steroids, reported interventions of the Immune Modulation Therapy Domain: tocilizumab, sarilumab (combined to a single IL-6 arm) and no immune modulation, and interventions of the Anticoagulation Domain: prophylaxis dose anticoagulation, therapeutic anticoagulation.

‡ Interventions and specified interactions, age, sex, site, time, Immunoglobulin Domain interventions: control, convalescent plasma. All Models are structured such that a higher OR is favorable.

#### Table S4. Secondary outcomes

| Outcome or Analysis | Convalescent plasma (N = 1078) | Control (N = 909) |
| --- | --- | --- |
| **Other Secondary Outcomes, restricted to Immunoglobulin Domain participants and any previously reported interventions and domains (n=3446)** | | |
| **28-day survival (time to event)** | N = 1072* | N = 900 |
| Adjusted HR - mean (SD) | 1.05 (0.08) | 1 |
| - median (95% Credible Interval) | 1.05 (0.91, 1.20) | 1 |
| Probability of superiority to control, % | 74.0 | - |
| Probability of odds ratio < 1.2 compared to control, %† | 98.1 | - |
| **90-day survival (time to event)** | | |
| Adjusted HR - mean (SD) | 1.05 (0.07) | 1 |
| - median (95% CrI) | 1.05 (0.92, 1.19) | 1 |
| Probability of superiority to control, % | 75.3 | - |
| Probability of odds ratio < 1.2 compared to control, %† | 98.1 | - |
| **Respiratory support-free days** | N=1074* | N=902 |
| Median (IQR) | 0 (-1, 15) | 2 (-1, 16) |
| Adjusted OR - mean (SD) | 0.95 (0.08) | 1 |
| - median (95% CrI) | 0.95 (0.81, 1.11) | 1 |
| Probability of superiority to control, % | 25.7 | - |
| Probability of odds ratio < 1.2 compared to control, %† | 99.8 | |
| **Cardiovascular support-free days** | N=1074* | N=902 |
| Median (IQR) | 14(-1, 21) | 14.5(-1, 21) |

| | | |
|---|---|---|
| Adjusted OR - mean (SD) | 0.96 (0.08) | 1 |
| - median (95% Credible Interval) | 0.95 (0.80,1.13) | 1 |
| Probability of superiority to control, % | 29.4 | - |
| Probability of odds ratio < 1.2 compared to control, %† | 99.6 | |
| **Length of ICU stay** | N=1075* | N=905 |
| Adjusted HR - mean (SD) | 0.94 (0.05) | 1 |
| - median (95% Credible Interval) | 0.94 (0.85, 1.04) | 1 |
| Probability of superiority to control, % | 10.1 | - |
| Probability of odds ratio < 1.2 compared to control, %† | >99.9 | |
| **Length of hospital stay** | N=1075* | N=905 |
| Adjusted HR - mean (SD) | 0.96 (0.05) | 1 |
| - median (95% Credible Interval) | 0.95 (0.86, 1.06) | 1 |
| Probability of superiority to control, % | 18.5 | - |
| Probability of odds ratio < 1.2 compared to control, %† | >99.9 | |
| **WHO scale at day 14** | N=1075* | N=905 |
| Adjusted OR - mean (SD) | 0.92(0.08) | 1 |
| - median (95% Credible Interval) | 0.92 (0.79, 1.08) | 1 |
| Probability of superiority to control, % | 15.5 | - |
| Probability of odds ratio <1.2 compared to control, %† | >99.9 | |
| **Progression to invasive mechanical ventilation, ECMO or death, restricted to those not intubated at baseline (n=1307)*** | | |
| Free of invasive mechanical ventilation at baseline, n (%) | N=701 | N=606 |

| | | |
|---|---|---|
| Progression to intubation, ECMO or death, n (%) | 347(49.5) | 275 (45.4) |
| Adjusted OR - mean (SD) | 0.83 (0.10) | 1 |
| - median (95% Credible Interval) | 0.82 (0.65,1.03) | 1 |
| Probability of superiority to control, % | 4.7 | - |
| Probability of odds ratio < 1.2 compared to control, %[†] | 99.9 | |
| **Safety Outcomes, restricted to Immunoglobulin Domain participants (N=1980)** | | |
| **Serious Adverse Events (n=1980)[*, ‡]** | | |
| Patients with ≥1 serious adverse event, n/N (%) | 32/1075 (3.0) | 12/905 (1.3) |
| **Venous thromboembolic events at 90-days (n=1980)[a]** | | |
| Patients with ≥ 1 venous thromboembolic event at 90-days, n/N (%) | 74/1075 (6.9) | 61/905 (6.7) |

[*] Data for secondary analyses excluded participants who had been randomized within another domain within the moderate stratum and then randomized to the immunoglobulin domain in the severe stratum (excluded 7 participants), maximum of 1980 participants included within secondary analyses and sensitivity analyses.

[†] An odds ratio < 1.2 equates to the threshold for futility for the primary outcome

[‡] Details of the adverse events reported are available in Table S7. Only 1 adverse event was definitely or probably related to convalescent plasma, a transfusion reaction.

#### Table S5: Sensitivity Analyses to Primary Outcomes

| Outcome or Analysis | Convalescent plasma (N = 1075) | Delayed convalescent plasma (N = 11) | Control (N = 904) |
|---|---|---|---|
| **Sensitivity analysis of OSFD for Unblinded ITT population with site and time factors removed[†]** | | | |
| Adjusted OR - mean (SD) | 0.96 (0.08) | 0.60 (0.30) | 1 |
| - median (95% Credible Interval) | 0.96 (0.82 to 1.13) | 0.54 (0.20 to 1.34) | 1 |
| Probability of superiority to control, % | 30.9 | * | - |
| **Sensitivity analysis of OSFD for the Convalescent Plasma Per Protocol population** | | | |
| Median (IQR) | 0 (-1 to 15) | * | 3 (-1 to 16) |
| Adjusted OR - mean (SD) | 0.93 (0.08) | * | 1 |
| - median (95% Credible Interval) | 0.92 (0.78 to 1.10) | * | 1 |
| Probability of superiority to control, % | 17.9 | * | - |
| **Sensitivity analysis of hospital survival for Unblinded ITT population with site and time factors removed[†]** | | | |
| Adjusted OR - mean (SD) | 1.04 (0.10) | 1.04 (0.64) | 1 |
| - median (95% Credible Interval) | 1.03 (0.85 to 1.25) | 0.89 (0.29 to 2.66) | 1 |
| Probability of superiority to control, % | 63.0 | * | - |
| **Sensitivity analysis of hospital survival for the convalescent plasma per protocol population** | | | |
| In-hospital mortality n/N (%) | 335/918 (36.5) | | 342/895 (38.2) |
| Adjusted OR - mean (SD) | 1.05 (0.12) | | |
| - median (95% Credible Interval) | 1.05 (0.85 to 1.30) | | 1 |
| Probability of superiority to control, % | 66.7 | | - |

| |
|---|
| * Analysis not performed<br>† age, sex, Immunoglobulin Domain interventions: control, convalescent plasma, delayed convalescent plasma, Covid-19 Antiviral Therapy Domain interventions: control, lopinavir–ritonavir, hydroxychloroquine, combination therapy, Corticosteroid Domain interventions: no steroids, fixed-duration corticosteroids, and shock-dependent steroids, reported interventions of the Immune Modulation Therapy Domain: tocilizumab, sarilumab (combined to a single IL-6 arm) and no immune modulation, and interventions of the Anticoagulation Domain: prophylaxis dose anticoagulation, therapeutic anticoagulation. |

Table S6. Interactions with other domains

| | Adjusted odds ratio (Mean) | Adjusted odds ratio (SD) | Adjusted odds ratio (Median) | Adjusted odds ratio (Credible Interval) | Posterior probability superior to control, % |
|---|---|---|---|---|---|
| **Organ support-free days*** | | | | | |
| **Convalescent plasma** | 0.90 | 0.09 | 0.90 | 0.73 to 1.10 | 15.3 |
| **Pooled IL-6ra** | 1.48 | 0.20 | 1.46 | 1.13 to 1.91 | - |
| **Lopinavir-ritonavir** | 0.75 | 0.12 | 0.74 | 0.55 to 1.02 | - |
| **Therapeutic anticoagulation** | 0.93 | 0.12 | 0.92 | 0.71 to 1.20 | - |
| **Convalescent plasma*Lopinavir-ritonavir combination** | 1.23 | 0.36 | 1.18 | 0.67 to 2.07 | 71.9 |
| **Convalescent plasma*Pooled IL6ra combination** | 1.53 | 0.25 | 1.51 | 1.10 to 2.07 | 99.5 |
| **Convalescent plasma*Therapeutic anticoagulation combination** | 0.69 | 0.13 | 0.68 | 0.47 to 0.98 | 1.9 |
| **Convalescent plasma*Lopinavir-ritonavir interaction** | 1.84 | 0.55 | 1.76 | 1.00 to 3.13 | - |
| **Convalescent plasma*Pooled IL6ra interaction** | 1.16 | 0.17 | 1.15 | 0.87 to 1.53 | - |
| **Convalescent plasma*Therapeutic anticoagulation interaction** | 0.83 | 0.16 | 0.82 | 0.56 to 1.20 | - |
| **In-hospital mortality[a]** | | | | | |
| **Convalescent plasma** | 1.01 | 0.13 | 1.00 | 0.78 to 1.30 | 50.5 |
| **Pooled IL-6ra** | 1.51 | 0.25 | 1.49 | 1.06 to 2.07 | - |
| **Lopinavir-ritonavir** | 0.71 | 0.14 | 0.70 | 0.47 to 1.04 | - |
| **Therapeutic anticoagulation** | 0.96 | 01.6 | 0.94 | 0.69 to 1.31 | - |
| **Convalescent plasma*Lopinavir-ritonavir combination** | 1.34 | 0.51 | 1.26 | 0.62 to 2.56 | 73.8 |
| **Convalescent plasma*Pooled IL6ra combination** | 1.74 | 0.36 | 1.71 | 1.15 to 2.54 | 99.7 |

| | | | | | |
|---|---|---|---|---|---|
| **Convalescent plasma*Therapeutic anticoagulation combination** | 0.64 | 0.15 | 0.62 | 0.40 to 0.97 | 1.9 |
| **Convalescent plasma*Lopinavir-ritonavir interaction** | 1.92 | 0.72 | 1.80 | 0.90 to 3.71 | - |
| **Convalescent plasma*Pooled IL6ra interaction** | 1.17 | 0.22 | 1.15 | 0.80 to 1.64 | - |
| **Convalescent plasma*Therapeutic anticoagulation interaction** | 0.68 | 0.16 | 0.66 | 0.41 to 1.04 | - |

[*] All interventions and specified interactions, age, sex, site, time, Immunoglobulin Domain interventions: control, convalescent plasma, delayed convalescent plasma, Covid-19 Antiviral Therapy Domain interventions: control, lopinavir−ritonavir, hydroxychloroquine, combination therapy, Corticosteroid Domain interventions: no steroids, fixed-duration corticosteroids, and shock-dependent steroids, reported interventions of the Immune Modulation Therapy Domain: tocilizumab, sarilumab (combined to a single IL-6 arm) and no immune modulation, and interventions of the Anticoagulation Domain: prophylaxis dose anticoagulation, therapeutic anticoagulation. Additional interactions between convalescent plasma and lopinavir−ritonavir, therapeutic anticoagulation, and pooled IL-6ra interventions are reported. All interaction effects are estimated using standard normal priors. The SAP specified that all interactions between convalescent plasma and unblinded interventions would be explored. Supplementary table 2 shows a breakdown of patients randomized to the Immunoglobulin and other unblinded domains. Due to small sample sizes, interactions between convalescent plasma and corticosteroids and HCQ/combination therapy were not specified. The model therefore estimates interactions between convalescent plasma and the pooled IL-6ra interventions, lopinavir-ritonavir, and therapeutic-dose anticoagulation.

### Table S7 – Serious Adverse Events

| Serious Adverse Event (SAE) | Convalescent Plasma | Control |
|---|---|---|
| **Total SAEs** | 32 | 12 |
| **Total Excluding thrombotic events*** | 18 | 7 |
| **Bleeding (All)†** | 7 | 4 |
| Cerebral | 1 | 1 |
| Gastrointestinal | 2 | 1 |
| Pulmonary | 1 | 0 |
| Other | 3 | 2 |
| **Thrombotic events (All)*** | 14 | 6 |
| Cerebrovascular accident | 6 | 1 |
| Myocardial infarction | 2 | 0 |
| Pulmonary embolism | 4 | 4 |
| Deep venous thrombosis | 1 | 0 |
| Multiple sites | 2 | 0 |
| **Elevated enzymes (Total)‡** | 4 | 1 |
| Creatinine kinase | 1 | 1 |
| ALT | 4 | 0 |
| **Transfusion reaction** | 1§ | 0 |
| **Multi-organ failure** | 1 | 0 |
| **Cardiac arrythmia** | 0 | 1 |
| **Cardiac arrest** | 2 | 0 |
| **Infection (All)** | 2 | 1 |
| Fever | 1 | 1 |
| Cytomegalovirus colitis | 1 | 0 |

* Thrombotic events also reported as a secondary outcome and therefore not all thrombotic outcomes reported as an SAE. Please see secondary outcomes for a more complete report of these outcomes. One of the multiple site thrombotic events was reported as HITT (heparin-induced thrombotic thrombocytopenia)

† Some of the bleeding adverse events were related to other randomised interventions such as therapeutic anticoagulation and antiplatelet agents.

**‡ Some of the elevated blood results were related to other randomised drug interventions or concomitant drug therapy which were stopped.**

**§ Classified as a febrile, allergic or hypotensive reaction after review by Serious Hazards of Transfusion (SHOT) panel (Hemovigilance reporting system in UK). No other cases reported to SHOT. This was the only serious adverse event classified as definitely or probably related to the intervention.**

#### 5. References

1. Walker G, Naing Z, Ospina Stella A, et al. SARS Coronavirus-2 Microneutralisation and Commercial Serological Assays Correlated Closely for Some but Not All Enzyme Immunoassays. Viruses 2021;13:247.
2. Beaudoin-Bussieres G, Laumaea A, Anand SP, et al. Decline of Humoral Responses against SARS-CoV-2 Spike in Convalescent Individuals. mBio 2020;11.
3. Anand SP, Prevost J, Richard J, et al. High-throughput detection of antibodies targeting the SARS-CoV-2 Spike in longitudinal convalescent plasma samples. Transfusion 2021;61:1377-82.
4. Tauzin A, Nayrac M, Benlarbi M, et al. A single BNT162b2 mRNA dose elicits antibodies with Fc-mediated effector functions and boost pre-existing humoral and T cell responses. bioRxiv 2021.
5. Abe KT, Li Z, Samson R, et al. A simple protein-based surrogate neutralization assay for SARS-CoV-2. JCI Insight 2020;5.
6. Epstein J, Burnouf T. Points to consider in the preparation and transfusion of COVID-19 convalescent plasma. Vox sanguinis 2020.
7. Harvala H, Mehew J, Robb ML, et al. Convalescent plasma treatment for SARS-CoV-2 infection: analysis of the first 436 donors in England, 22 April to 12 May 2020. Eurosurveillance 2020;25.
8. Zilla M, Wheeler B, Keetch C, et al. Performance of commercially available EUA SARS-CoV-2 antibody assays in critically ill and mildly symptomatic patients assessed at a large academic medical center. American Journal of Clinical Pathology 2020;155:343-53.
9. Klimstra W, Tilston-Lunel N, Nambulli S, et al. SARS-CoV-2 growth, furin-cleavage-site adaptation and neutralization using serum from acutely infected, hospitalized COVID-19 patients. Journal of General Virology 2020;101:1156-69.
10. Tan CW, Chia WN, Qin X, et al. A SARS-CoV-2 surrogate virus neutralization test based on antibody-mediated blockage of ACE2–spike protein–protein interaction. Nature Biotechnology 2020;38:1073-8.
11. Caly L, Druce J, Roberts J, et al. Isolation and rapid sharing of the 2019 novel coronavirus ( SARS - CoV-2) from the first patient diagnosed with COVID -19 in Australia. Medical Journal of Australia 2020;212:459-62.
12. Subbarao K, McAuliffe J, Vogel L, et al. Prior Infection and Passive Transfer of Neutralizing Antibody Prevent Replication of Severe Acute Respiratory Syndrome Coronavirus in the Respiratory Tract of Mice. Journal of Virology 2004;78:3572-7.
13. Reed LJ, Muench H. A simple method of estimating fifty per cent endpoints. American Journal of Epidemiology 1938;27:493-7.
14. Ainsworth M, Andersson M, Auckland K, et al. Performance characteristics of five immunoassays for SARS-CoV-2: a head-to-head benchmark comparison. The Lancet Infectious Diseases 2020;20:1390-400.
15. Ratcliff J, Nguyen D, Fish M, et al. Virological and serological characterization of critically ill patients with COVID-19 in the UK: a special focus on variant detection. Cold Spring Harbor Laboratory; 2021.
16. Piechotta V, Iannizzi C, Chai KL, et al. Convalescent plasma or hyperimmune immunoglobulin for people with COVID-19: a living systematic review. Cochrane Database of Systematic Reviews 2021.

#### 6. Appendices

**Appendix A – Statistical Analysis Committee Primary Analysis Report**
**Appendix B - ITSC Secondary Analysis Report**
